# A Manual of Procedures for the Generation of the AI-Ready and Exploratory Atlas for Diabetes Insights (AI-READI) Database

**DOI:** 10.64898/2026.03.30.26349552

**Authors:** Dawn S. Matthies, Jeffrey C. Edberg, Sally L. Baxter, Aaron Y. Lee, Cecilia S. Lee, Gerald McGwin, Julia P. Owen, Linda M. Zangwill, Cynthia Owsley, the AI-READI Consortium

**Affiliations:** Department of Ophthalmology & Visual Sciences, Heersink School of Medicine, University of Alabama at Birmingham, Birmingham, AL; Department of Medicine, Heersink School of Medicine, University of Alabama at Birmingham, Birmingham AL; Division of Ophthalmology Informatics and Data Science, Hamilton Glaucoma Center, Viterbi Family Department of Ophthalmology and Shiley Eye Institute, University of California, San Diego, La Jolla, CA, USA; Division of Biomedical Informatics, Department of Medicine, University of California, San Diego, La Jolla, CA, USA; John F. Hardesty MD Department of Ophthalmology and Visual Sciences, Washington University, St. Louis, MO; University of Washington Department of Ophthalmology, Seattle, WA, The Roger and Angie Karalis Johnson Retina Center, Seattle, WA; Department of Epidemiology, University of Alabama at Birmingham, Birmingham AL

## Abstract

The ability to understand and affect the course of complex, multi-system diseases like diabetes has been limited by a lack of well-designed, high-quality and large multimodal datasets. The NIH Bridge2AI AI-READI project (aireadi.org) aims to address this shortfall by generating an AI-ready dataset to support AI discoveries in type 2 diabetes mellitus (T2DM). This manual of procedures provides a detailed description of the AI-READI protocol.

## 1. PROJECT SUMMARY AND STUDY OBJECTIVES

### 1.1 Introduction

The Artificial Intelligence Ready and Exploratory Atlas for Diabetes Insights (AI-READI) project is designed to develop a flagship, ethically sourced dataset that will support future artificial intelligence and machine learning (AI/ML) research. Our goal is to enable the generation of critical insights into type 2 diabetes mellitus (T2DM), including the identification of salutogenic pathways that promote a return to health. The ability to understand and affect the course of complex, multi-organ diseases such as T2DM has been limited by a lack of well-designed, high quality, large, and inclusive multimodal datasets. The AI-READI team of investigators aim to collect a cross-sectional dataset of 4,000 people that is approximately balanced for diabetes disease stage. Data collection is specifically designed to permit downstream pseudotime manifold analysis, an approach used to predict disease trajectories by collecting and learning from complex, multimodal data from participants with differing disease severity (normal to insulin-dependent T2DM). The long-term objective for this project is to develop a foundational dataset in T2DM, agnostic to existing classification criteria or biases, which can be used to reconstruct a temporal atlas of T2DM development and reversal towards health. Data will be optimized for downstream AI/ML research and made available to researchers through a registered, public access or a controlled access database, depending on the variables requested. This project will also create a roadmap for best practices in standardizing, archiving, sharing, and protecting datasets for AI/ML research. An overview of the AI-READI study protocol is published separately.^1^

Specific project goals include:

1. Collect and share the AI-READI dataset for AI/ML research according to the Findable, Accessible, Interoperable, Reusable (FAIR) data principles.
2. Create a model for developing diverse and representative datasets.
3. Increase access to and quality of AI/ML research by recruiting and training personnel with diverse backgrounds.

## 2. Study Design

AI-READI is a multicenter, cross-sectional study.

### 2.1 Data Acquisition - Domains Addressed in the AI-READI Dataset

### 2.2 Study Population

This cross-sectional study will enroll 4,000 people across three data collection sites, stratified by approximately equal numbers of four levels of T2DM severity: no diabetes, prediabetes/lifestyle-controlled, diabetes controlled with oral medications or non-insulin injectable medications, and insulin-controlled diabetes. The dataset will include approximately equal numbers of participants from each data site. The study population will encourage enrollment of several race/ethnicities and approximately equal numbers of males and females. Data collection will occur at three geographic areas of the US: Birmingham AL (University of Alabama at Birmingham, UAB), Seattle WA (University of Washington, UW), and San Diego County CA (University of California at San Diego, UCSD). All participants will be aged 40 or above.

### 2.3 Study Timeline

The proposed timeline for the AI-READI project takes place over four years (Figure 3). Year one focuses on the development of the protocol, training and certification of clinical research coordinators (CRCs), and piloting the protocol with 10 volunteer participants per site. The first 10 participants will allow for the identification of areas within the protocol that need refinement. After protocol adjustment (if necessary), an additional 20 participants per site will be recruited in year 1. In years 2-4, ∼1300 participants per site will be recruited, with a goal of 4000 participants recruited overall.

**Figure 1.**
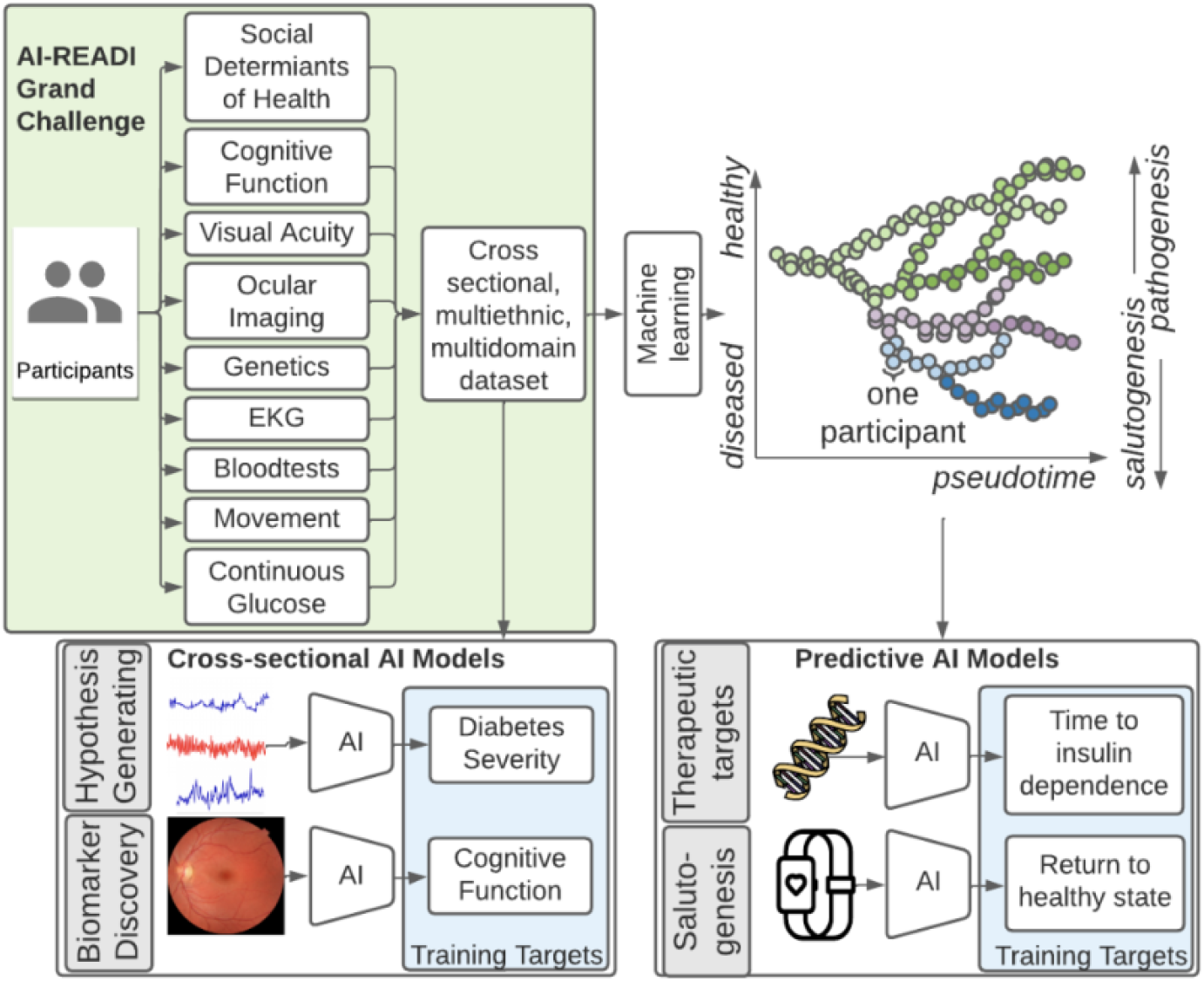
A large, multi-domain dataset will be collected to enable downstream pseudotime manifolds and numerous AI applications.

**Figure 2.**
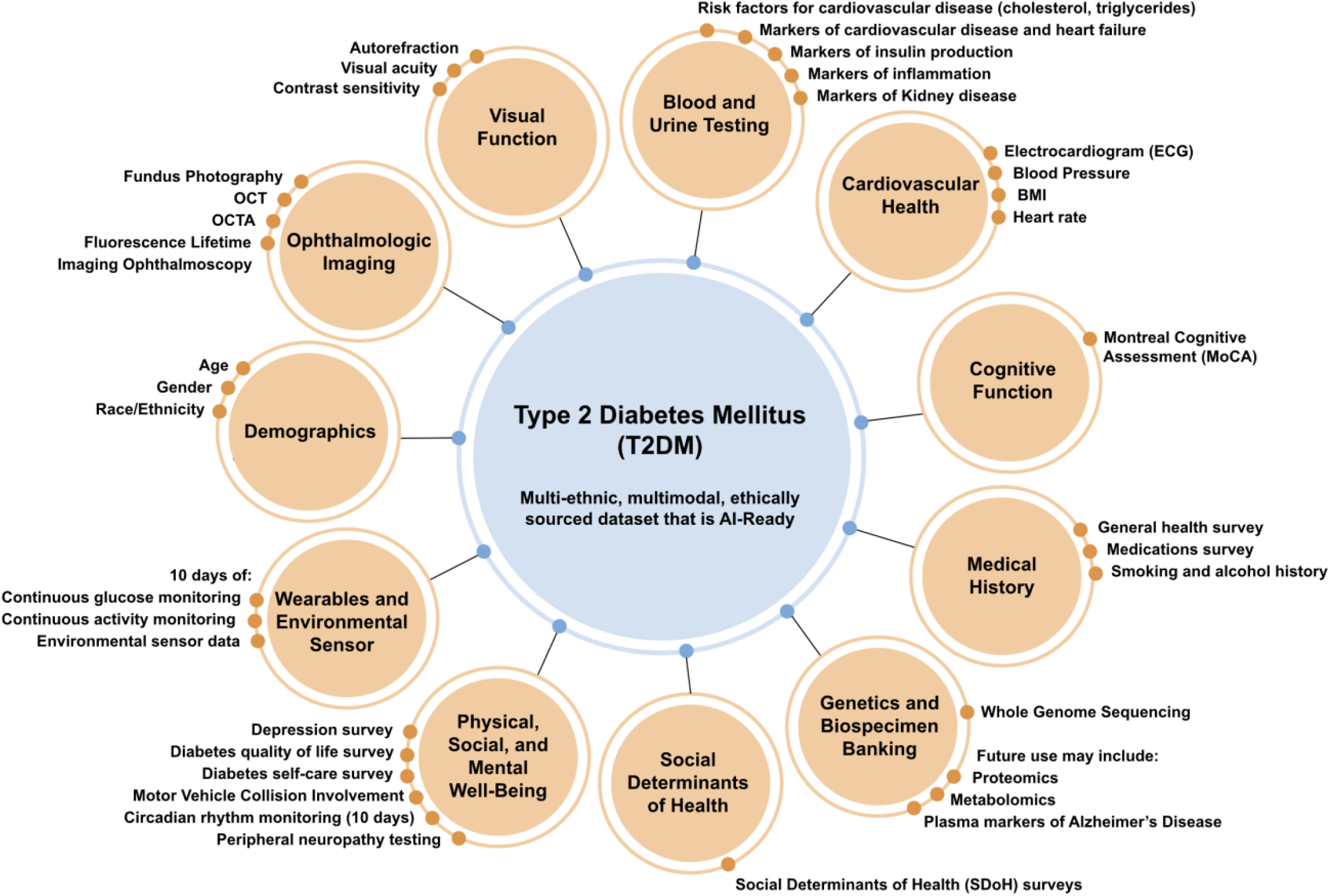
This project will generate an accessible, shared dataset that includes a diverse set of health and behavioral domains and is harmonized across all variables to be AI/ML ready.

**Figure 3.**
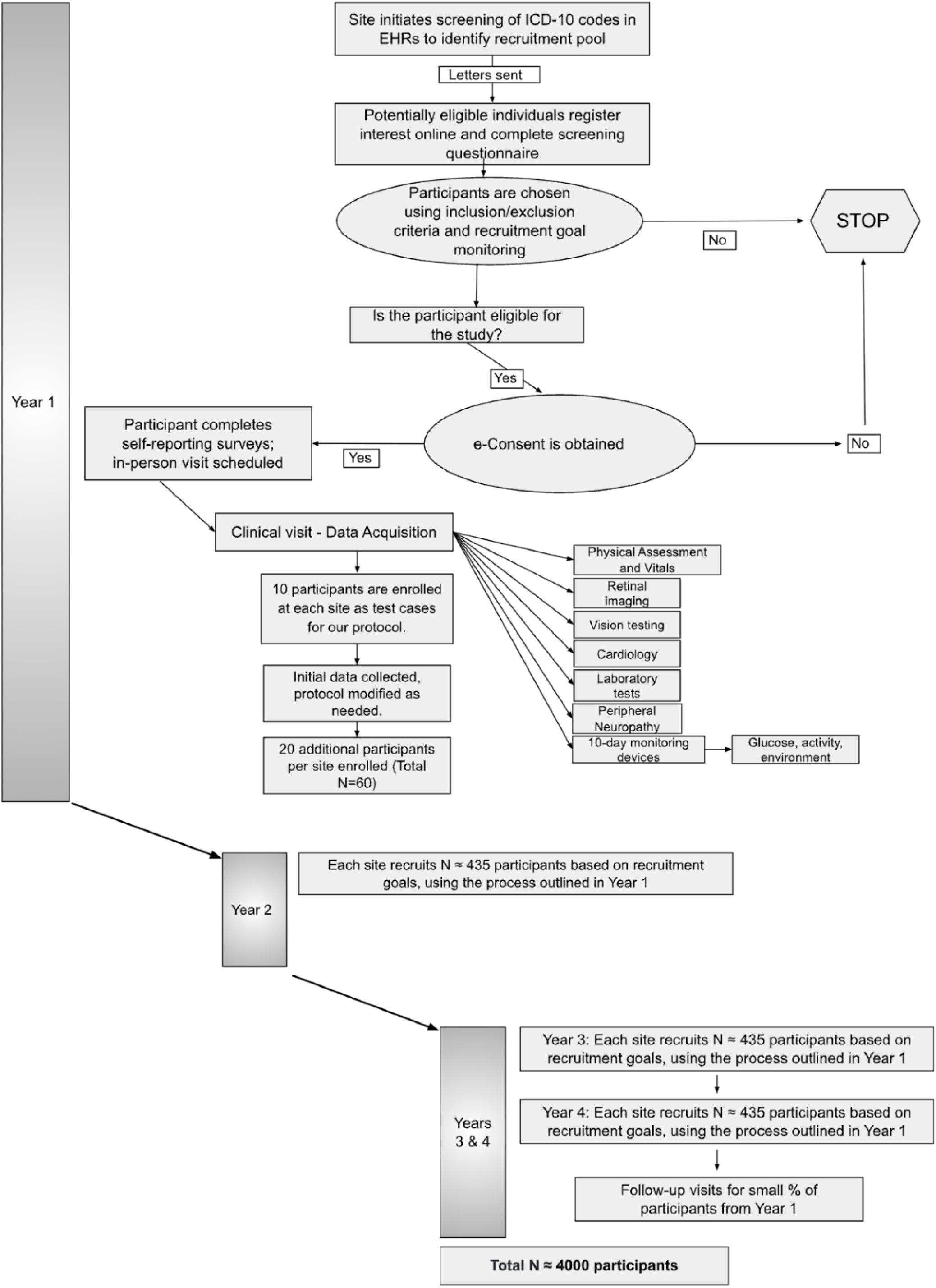
The AI-READI study timeline will include a pilot phase in year one to refine the protocol, followed by 3 years of ∼1300 participants per year at each of the 3 sites.

### 2.4 Summary of Full Participation in AI-READI

Voluntary participation in the AI-READI research program involves three key stages: the completion of questionnaires on health, lifestyle and functioning (electronically at home or in person), a single 3-4 hour in-person clinical visit at the data collection site local to the participant, and 10 days of at home monitoring with 2 wearable devices and a plug-in environmental sensor (Figure 4). At the end of the monitoring period, participants will be compensated for their participation upon return of the monitoring devices.

**Figure 4.**
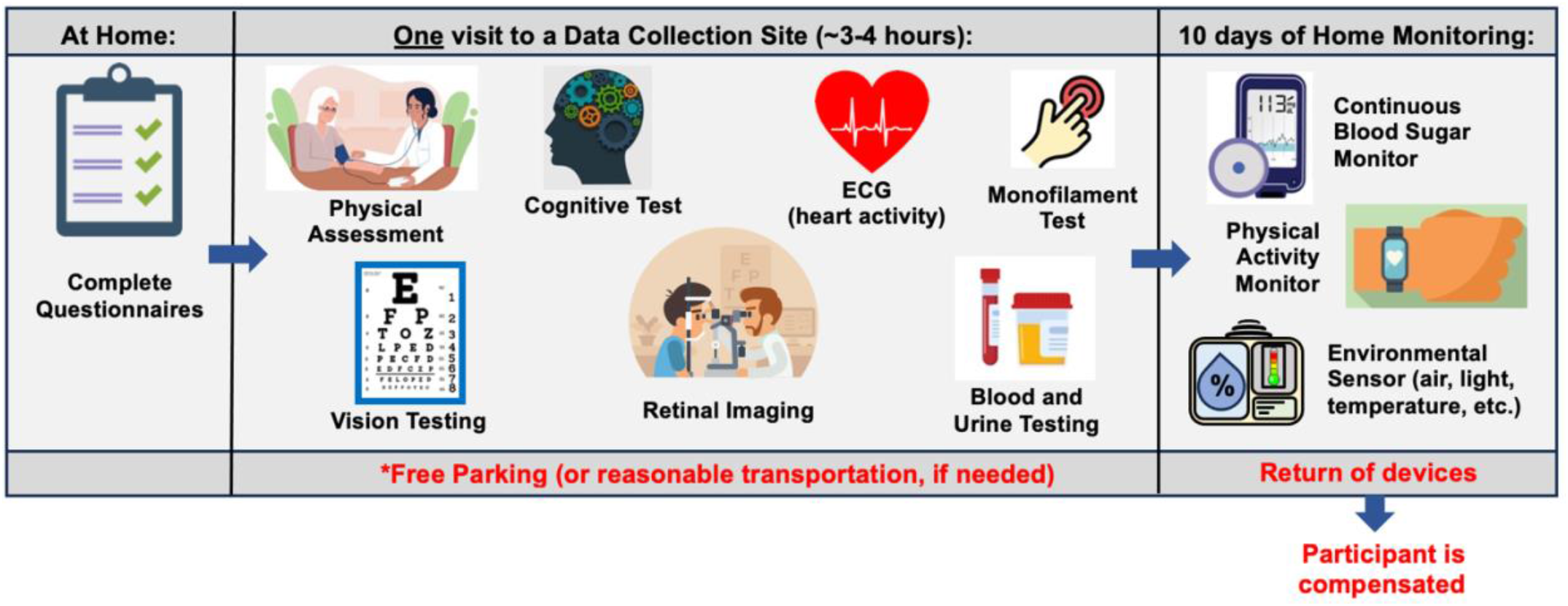
Basic overview of full participation in the Al-READI research program.

### 2.5 Specific Data Domains and all Variables to be collected in the AI-READI project

Table 1 lists the specific domains and variables that will be collected in the AI-READI study and included in the FAIRhub.io database. Also included are the Biorepository samples. See Sections 5.13 and 5.16.1 for more details on blood and urine testing and Sections 5.11 and 5.12.3 for specific devices used and data formats.

**Table 1.** Domains and Variables to be collected and included in the AI-READI dataset.

| Domain | Variable | Rationale for Inclusion | Source |
| --- | --- | --- | --- |
| Demographic | Age | Eligibility criterion | person |
|  | Gender | Cohort stratification | person |
|  | Race/ethnicity |  | person |
| Physical assessment | Height | Known risk factor for Type 2 diabetes | device |
|  | Weight | " | device |
|  | Waist to Hip Ratio (WHR) | " | calculation |
|  | Body Mass Index (BMI) | " | calculation |
| Questionnaires | General health history | Indicate known risk factors and complications | person |
|  | Depression (CES-D-10) | Depression is comorbid with T2DM | person |
|  | Social Determinants of Health (PhenX Toolkit) | Investigates health inequities among people with T2DM | person |
|  | Dietary habits | Diet impacts development and severity of T2DM | person |
|  | Smoking and alcohol history, marijuana use and vaping | Smoking and alcohol use can impact T2DM |  |
|  | Diabetes Quality of Life (PAID-5) | Investigates the impact of T2DM on well-being | person |
|  | Diabetes Self-Care (Diabetes Score Survey) | Investigates dietary habits, fitness, and self-care | person |
|  | Medications (self report) | Current medications may influence T2DM | person |
|  | Diabetes severity (self report) | Initial screening process for participation; self-report of treatment approach | person |
|  | Visual Impairment and Access to Eye Care | Self-assessment of visual function information about eye care | person |
| Vision | Autorefraction | Vision is affected by hyperglycemia | person |
|  | Best-Corrected Visual Acuity (BCVA) - photopic | Vision is affected by hyperglycemia | person |
|  | BCVA mesopic (low luminance) | " | person |
|  | Contrast sensitivity - photopic | Indicator of retinal health | person |
|  | Low luminance contrast sensitivity | " | person |
| Cognition | Montreal Cognitive Assessment (MoCA) | Cognitive Impairment is comorbid with T2DM | person |
| Retinal Imaging | Fundus photo (both undilated and dilated) | Investigates retinal and general health | device |
|  | OCT | " | device |
|  | OCTA | " | device |
|  | FLIO (Heidelberg FLIO) | " | device |
| Cardiology | ECG | Assesses risk for coronary artery disease | device |
|  | Blood pressure | Indicator of hypertension | device |
|  | Resting Heart Rate (HR) | High HR is associated with heart disease and T2DM |  |
| Peripheral Neuropathy | Monofilament test | Peripheral neuropathy is comorbid with T2DM | person |
| Laboratory tests | Plasma: |  |  |
|  | N-terminal pro-B-type natriuretic peptide (NT-proBNP) | Severity and outcome predictor of heart failure | Lab test |
|  | Troponin-T | Marker of myocardial injury | Lab test |
|  | C-Peptide | Indicator of insulin production | Lab test |
|  | Insulin | Marker for diabetes | Lab test |
|  | Serum: |  |  |
|  | C-Reactive Protein, high sensitivity (CRP-HS) | Inflammation marker; risk factor for T2DM | Lab test |
|  | Lipid panel | Cardiovascular disease risk factor | Lab test |
|  | Glucose | Marker for diabetes | Lab test |
|  | Blood Urea Nitrogen (BUN) | Indicator of kidney function | Lab test |
|  | Creatinine | Indicator of kidney function | Lab test |
|  | Electrolytes | Markers of metabolic health | Lab test |
|  | Protein, total | Useful tool for overall health status | Lab test |
|  | Albumin (A) | Useful tool for overall health status | Lab test |
|  | Globulin (G), total | Useful tool for overall health status | calculated |
|  | Bilirubin | Marker of liver health | Lab test |
|  | Alkaline phosphatase | Marker of liver health | Lab test |
|  | Aspartate aminotransferase (AST) | Marker of liver health | Lab test |
|  | Alanine aminotransferase (ALT) | Marker of liver health | Lab test |
|  | Whole blood: |  |  |
|  | HbA1c (%) | Marker for diabetes | Lab test |
|  | Complete Blood Count (CBC) | Useful tool for overall health status | Lab test |
|  | Urine: |  |  |
|  | Urine albumin | Urinary biomarker for early diabetic nephropathy | Lab test |
|  | Urine creatinine | Urinary biomarker for early diabetic nephropathy | Lab test |
| Physical Activity monitor | Activity Monitoring: #steps, oxygen saturation, heart rate | Potential biomarker and confounder of diabetes related outcomes | device |
|  | Sleep duration (circadian and diurnal rhythm) | Potential biomarker of diabetes related complications | device |
| Continuous Glucose Monitoring | CGM | Indicator of diabetes and marker for diabetes management | device |
| Environmental sensor | Air quality measurements | Investigates quality of air in home environment (e.g. PM10, PM 2.5) | device |
| Other | Motor vehicle accident reports | Used to measure driver safety | Outside source |
| Biospecimens | Buffy coat samples | DNA isolations for future studies such as whole genome sequencing | Lab storage |
|  | Genomic DNA | Genomic applications | Lab storage |
|  | Plasma samples | Future proteomics and metabolomics studies | Lab storage |
|  | Serum samples | Future proteomics and metabolomics studies | Lab storage |
|  | PAXgene RNA samples | Future RNA isolations | Lab storage |
|  | Peripheral Blood Mononuclear Cells (PBMCs) | PBMCs for future immunologic studies; generation of iPSCs | Lab storage |

## 3. Onboarding, Training and Certification of Project Members

### 3.1 Bridge2AI, the NIH Common Fund and the Grand Challenge

An overall introduction to the Bridge2AI program may be found at http://commonfund.nih.gov/bridge2ai. All members of the AI-READI project will use the Bridge2AI site to familiarize themselves with the goals of this ambitious program. Through the Bridge2AI website, members will learn about the Grand Challenge and the AI-READI response to a call for projects targeting salutogenesis.

### 3.2 Regulatory Training Components

Each Data site will be responsible for their Clinical Research Coordinators (CRCs) and other relevant staff meeting regulatory training components. These are described below.

#### HIPAA

Each institution has its own specific HIPAA course. CRCs will complete the course associated with their specific institution.

#### IRB/GCP/RCR

IRB, Good Clinical Practice (GCP), and Responsible Conduct of Research (RCR) training certifications are required for AI-READI since human subjects are involved. All three courses are available on the CITI Program website (https://about.citiprogram.org/). Trainees may access the courses through their institution.

#### Bloodborne Pathogens

All project members who will be in contact with blood samples (including preparing blood samples for transport and simply transporting tubes) must complete training and certification in the OSHA Bloodborne pathogens training course, as required under CFR 29 - 1910.1030. The Bloodborne Pathogens training course may be accessed online at https://www.nationaloshafoundation.com/blood-borne-pathogen/ or through each institution’s training division.

#### Shipping and Transporting Regulated Biological Samples

Project members who will be shipping blood or other biohazardous samples are required to complete a Shipping and Transport of Regulated Biological Samples training and certification course, as required by the International Air Transport Association (IATA) and the U.S. Department of Transportation (DOT). Trainees may access this course through the CITI program website (https://about.citiprogram.org) or through their institution.

#### Training in informed consent

The CRC must be trained in the process of obtaining informed consent in the event that a participant does not complete informed consent electronically from home, or if the participant has questions about the informed consent process. CRCs should contact the appropriate person at their site for informed consent training or any other question about how to meet regulatory requirements on the items above.

### 3.3 Certification for Collecting Data on Variables

This section focuses on the certification process for variables collected in the protocol. The instructions on how to collect the data for each variable are detailed in Section 5. The data/project manager at each data site will oversee and manage the process of certifying CRCs at their site.

#### 3.3.1 Certification of Non-Retinal Imaging Data Elements

##### MoCA Training and Certification

A one-hour training module is completed online via the MoCA website (https://www.mocatest.org/training-certification/). To access the MoCA training module, CRCs will need to create an online account and provide the IRB approval number (STUDY00016228). After successfully completing the training and properly administering the test to 3 volunteers, CRCs will be certified to administer the MoCA test. Certification must involve the administration of both electronic and paper forms of the MoCA test.

##### ECG

Philips (manufacturer of Pagewriter TC30 Cardiograph) will provide initial on-site training. Additional training materials, including videos, are available on the shared AI-READI Google drive. CRCs at each site will submit data from three volunteers to their site manager for approval.

##### Peripheral Neuropathy / Monofilament Test

The Monofilament Test will be performed on three volunteers (both feet) using the standard operating procedure listed in section 5.8 and observed by the site manager. Data will be collected on the data collection form and submitted to their site manager for review and certification.

##### Autorefraction and Visual acuity

Visual acuity testing using the M & S Technologies EVA-*e*-ETDRS device will be performed on three volunteers OD and OS, under photopic conditions and mesopic conditions, using the appropriate data collection forms (described in sections 5.10.2 and 5.10.3, respectively). Forms will be submitted to the site manager for review and certification.

##### Contrast Sensitivity

Contrast sensitivity testing using the Mars charts will be performed on three volunteers OD and OS, under photopic conditions and mesopic conditions, using the appropriate data collection forms (described in section 5.10.4). Data is then submitted to the site manager for review and certification.

##### Continuous Glucose Monitor

The Dexcom G6 process and instructions will be described to two volunteers. Successful explanations will be verified by the site manager. G6 units must be inserted on those two volunteers and initialized successfully. The G6 should be worn by the volunteers for a minimum of 24 hours and the data exported into a file to be submitted to the site manager for review (discussed in section 5.11.1). If the site manager deems the results acceptable, the CRC will be certified for CGM administration.

##### Physical Activity Monitor

Physical activity will be monitored via a Garmin watch during a 10-day period. Instructions on the following will be provided to two mock participants: wearing the device, battery check, charging instructions, and returning the watch in the prepaid shipping box. The Garmin Watch must be initiated and worn by the two volunteer participants for a minimum of 24 hours, the data downloaded after completion, and the watch properly prepared for the next participant as described in section 5.11.2. Exported data must be submitted to the site manager for review and if acceptable, the CRC will be certified for physical activity monitor administration.

##### Environmental Sensor

Various air pollutants will be tested via a device to be placed at home during a 10-day period. CRCs will explain to two volunteers how the device works, where to place it, and how to return the sensor in the shipping box (including the completion of the location card) following the instructions in section 5.11.3 for the Environmental Sensor. The site manager will export test data after 24 hours of collection and if the results are acceptable, the CRC will be certified for environmental sensor administration.

##### Dilation Procedures

A certified ophthalmic technician at each site will train CRCs on administering eye drops for dilation.

#### 3.3.2 Certification of Retinal Imaging Devices and Retinal Imaging Technicians

Each site is responsible for ensuring that retinal imaging technicians have been fully trained in the AI-READI protocol. Retinal imaging technicians must also be certified by IRB, GCP, and HIPAA. See section 3.2 for a description on how to complete these.

#### 3.3.3 Certification of Biospecimen Processing and Storage

Research staff from the UAB CCTS biorepository will travel to UCSD and UW to train technicians on the processing of blood and urine samples for storage and testing. Training will include the centrifugation of samples, the proper labeling of tubes, the scanning of tube bar codes, and the proper entry of all samples into appropriate manifests. The UAB CCTS team will decide if the technicians are proficient in AI-READI sample preparation and are ready for handling participant samples.

### 3.4 REDCap Access, Roles, and User Rights

REDCap is the data management system to be used for recruitment and screening, the administration of questionnaires, operational functions of the protocol, and clinical data entry from elements of the in-person visit. The AI-READI REDCap database is hosted by the University of Washington and maintained by the Oregon Health and Science University REDCap team.

#### 3.4.1 REDCap Roles and User Rights

The following roles and user rights have been established for team members using the AI-READI database, based on applying the principle of least privilege:

1. **Research Coordinators with Retinal Imaging**
  a. Assigned to a Data Access Group (DAG)
  b. View access to forms and reports; data entry; survey management; no export access
2. **Research Coordinator - Retinal Imaging only**
  a. Assigned to a DAG
  b. View access to retinal imaging forms and reports; data entry retinal imaging form
3. **Site Data Manager**
  a. Assigned to a DAG
  b. View and export reports for their site, data querying, limited export access for their site
  c. Edit access to Data Management form; read access to almost all forms
4. **Research Coordinator and Site Data Manager**
  a. Combined role (#1 and #3 above)
5. **Study Data Manager (no import)**
  a. Site data manager access, plus:
  b. Not assigned to a DAG (can view all site data)
6. **Site PIs**
  a. View access only for forms (no edit access)
  b. View access only for reports and dashboards (no export capability)
  c. Read access to data queries
7. **Recruitment Manager**
  a. Unrestricted export access
  b. No import access
8. **Export for Azure Storage Explorer**
  a. Export access for dashboards and transfer of data to Azure Storage Explorer
9. **REDCap Builders**
  a. Build/design access for development and maintenance of the database
10. **Lab Specimen Management**
  a. Edit access to lab specimen management forms only

#### 3.4.2 CRC Interviewer Codes

Coordinators at each site will be de-identified in REDCap and on devices. We have established designated Interviewer Codes that are unique for each CRC, and Interviewer Codes should be used in all places where the CRC’s name would normally be entered. The actual names of CRC should never be entered in REDCap forms or on data collection devices. Data Managers will provide each CRC at their site with their designated Interviewer Code.

## 4. Recruitment, Enrollment, Follow-up and Policies

This chapter will explain the recruitment strategy and enrollment procedures.

### 4.1. Recruitment Strategy Overview

The three study sites (UAB, UCSD, UW) will follow the same recruitment strategy for the purpose of harmonization and to minimize sampling bias. The following schematic outlines the approach we will use for recruitment (Figures 5a and 5b):

**Figure 5a and 5b.**
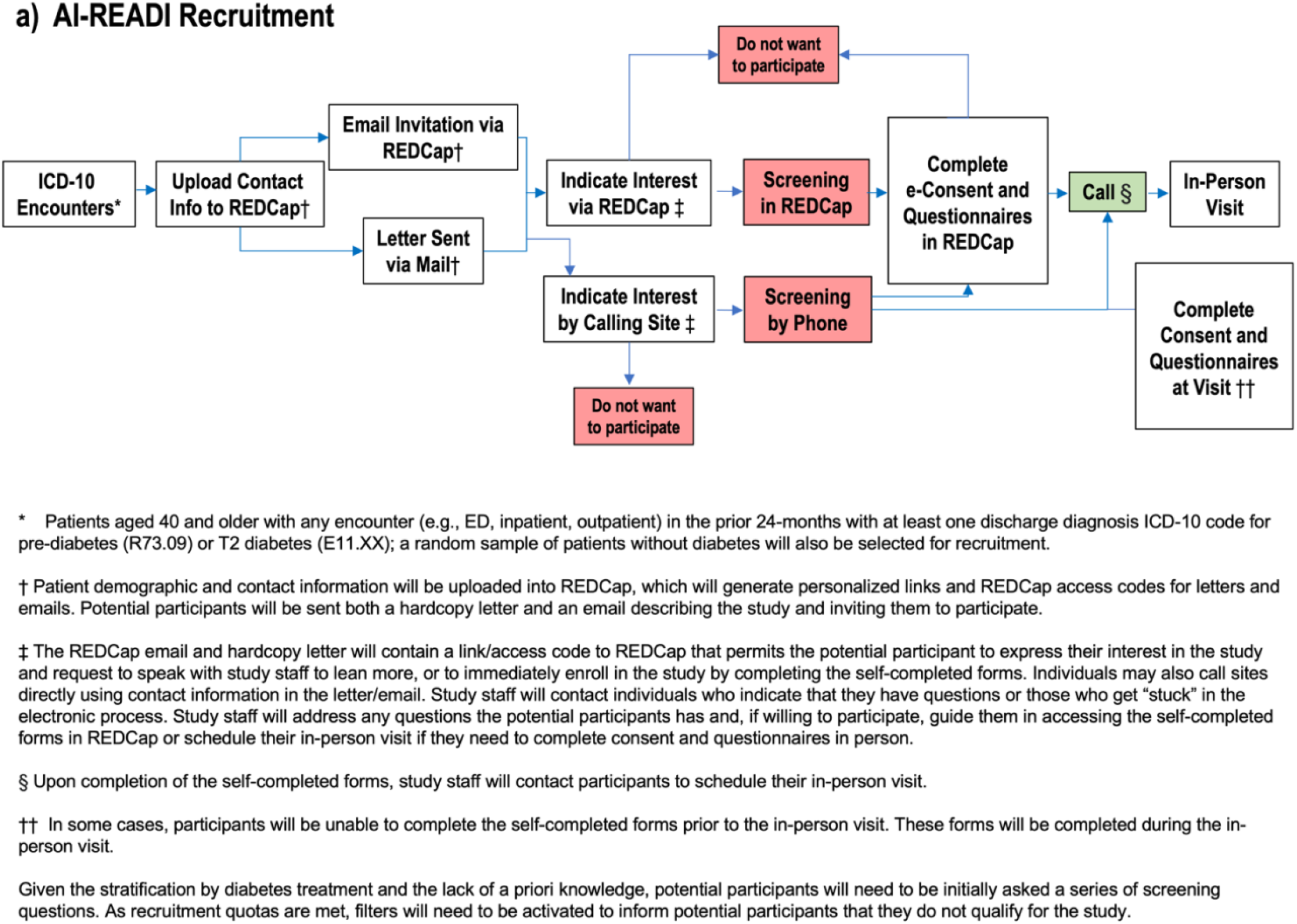
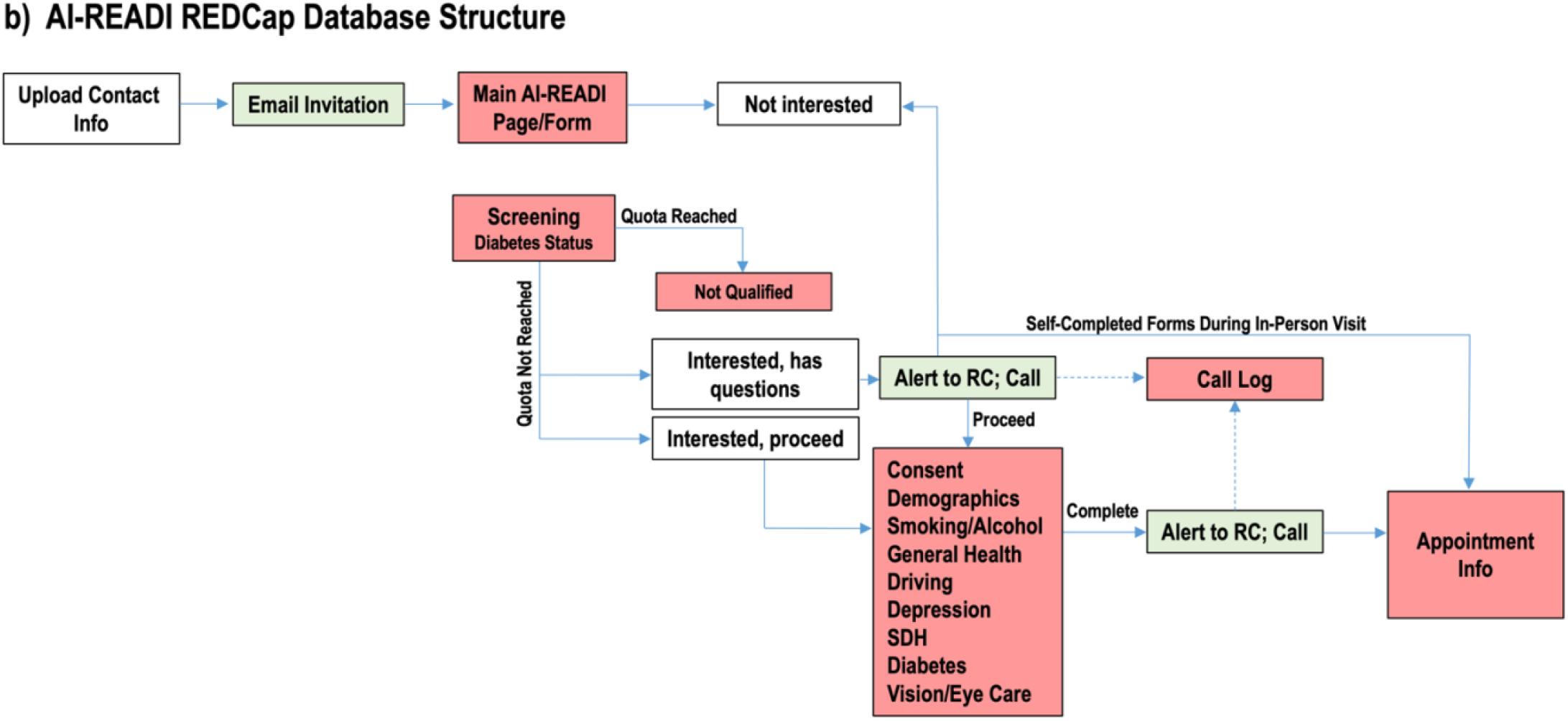
5a) Recruitment pools will be identified for each data site by screening Electronic Health Records (EHR) for diabetes and prediabetes ICD-10 codes for all patients who have had an encounter with their university health system within the past 2 years. Additional pools will be collected throughout the project to select for patients new to each health system since the prior extraction. 5b) Our recruitment strategy utilizes a REDCap database interface that allows for the self-completion of electronic informed consent and questionnaires prior to the in-person visit. Pathways also exist for the completion of informed consent and questionnaires in person, if a participant chooses not to complete these electronically.

### 4.2 Study Base and Sampling

The study base is all patients aged ≥ 40 years of age who had a medical encounter within each health system site (UAB, UCSD, UW) between 2020 and 2025. Patients with diabetes and pre-diabetes will be identified using ICD-10 diagnosis codes R73.09 and E11.X, respectively. The recruitment process (meaning the sending of letters and emails) will occur in waves in order to facilitate the efficient sampling of the study base. The composition and size of each wave will be influenced by the observed participation characteristics of severity of T2DM and site. The desire is for the study sample to reflect the composition explained in section 2.2. As recruitment progresses, we will actively monitor the composition of participants and adjust as needed. Recruiting in waves will also allow sufficient time for CRCs to respond to those who are interested and follow up on mailings with expediency.

### 4.3 Recruitment and Enrollment Procedures

For each wave of recruitment, a file containing participant contact and demographic information will be uploaded into the REDCap study database, where unique URLs and access codes will be auto-assigned to each individual. A file containing the name, address, data site ID, personalized URLs and access codes will be exported from REDCap and shared with our mailing service, Adapt Data, Inc., through a HIPAA-compliant server. Patients selected for recruitment will be initially contacted via two mechanisms: (1) a hard copy letter will be mailed to each selected participant describing the goals of the study and inviting them to participate (see Appendix B); and (2) REDCap will send an email to each participant that contains the same content as the hardcopy letter. Emails will be sent 7 days after hardcopy letters are mailed. If a participant does not have an email address, we will only use option #1. The purpose of this two-pronged approach is to ensure no potential participant is “left behind” (for example if they do not regularly read email, or if the email gets stuck in a spam filter, they still will receive the letter; or if they do not have an email in the health system). For letter mailing, AI-READI will use a mailing service for cost efficiency. The initial letter/email will give a brief overview of the project, explain the importance of the work, and convey a sense of excitement about the research. The letter/email will contain a personalized tinyURL (and a general link plus personalized access code) that will take them into the AI-READI REDCap database. Once they enter the database, they will be presented with an overview of the project and opportunities to view frequently asked questions about participation. They may also request a study team member to contact them to answer additional questions.

Upon receipt of the email and/or letter, the patient will have several pathways for recruitment into the study (Figures 5a and 5b):

- The potential participant can immediately enroll in the study by indicating in REDCap that they are interested and would like to determine if they are eligible. Indicating interest will take them into a screening questionnaire that will automatically determine if they screen in based on answers to diabetes questions, or screen out if they are ineligible (see inclusion/exclusion criteria in section 4.5). If they are screened in, they will be given the opportunity to complete informed consent and a HIPAA waiver electronically (“e-consent”) followed by completing the REDCap questionnaire forms on health and functioning electronically. These are questionnaires that address multiple aspects of current health, health history, and functioning (see section 5.5 for more details). After the REDCap questionnaires are received at the site, a CRC will phone the participant to schedule an in-person visit (section 4.7).
- For those who express general interest but request a call from the CRC and who do not complete e-consent, they will either indicate they want to participate during the call or decline participation. If they want to participate, they will be requested to complete e-consent and the REDCap questionnaire forms on health and functioning electronically. Once completed, the CRC will phone them to schedule a visit (section 4.7).
- For those who do not want to complete e-consent or the REDCap questionnaires on health and functioning electronically, but do want to enroll and be scheduled for a visit: they will be scheduled for a visit. The CRC will explain that the duration of their visit will be longer than the visits discussed above because they will have to go through the consent process and complete the questionnaires at the beginning of their CRU visit, before any other clinical elements are initiated. In this scenario, the CRC should ask the participant if they would like the consent documentation to be emailed to them for their review prior to coming to the visit.
- For those who do not wish to participate in the study, they will be asked if they are willing to share why they are not interested in participating and their answers recorded either directly REDCap (if participant replied electronically) or by CRCs in the REDCap call log (if verbally provided to the CRC during a phone call).
- Following the completion of e-consent by any individual, REDCap will send an email to the site-specific ai-readi@ email address indicating that the consent is ready for attestation. CRCs and Data Managers will monitor their site ai-readi email address and perform the attestation in REDCap prior to scheduling the in-person visit. **Note: in REDCap, the attestation form must be completed as a survey**.

### 4.4 Further Comments on Informed Consent

Informed consent to participate will be required before participation in any part of the protocol (including questionnaires). As described above, potential participants will be given the option to read all consent documentation electronically (e-consent) before their visit and give their consent with an electronic signature without verbal communication with a CRC. Potential participants will also be given the option to receive a phone call from a CRC for more information about the study prior to e-consent. Participants may access e-consent documentation in REDCap and decide at that point they do not want to participate or would like additional information. There will be pathways within REDCap for these possibilities. If they decide that they are no longer interested after reading the e-consent, then they will be asked to complete an optional, one question short survey indicating why they decided not to participate. They will also be given the option of exiting out of e-consent and requesting a phone call from a CRC for additional information.

If e-consent is not obtained prior to the scheduled in-person visit, the informed consent discussion will take place upon arrival at the visit and will occur in an area where privacy and confidentiality can be respected. E-consent documentation and the in-person consent process used by CRCs will describe the background/purpose of the project and the voluntary nature, commitment, risks, and benefits of participating (plus other elements of informed consent). Eligible individuals will always have the opportunity to ask questions prior to signing the consent form. Once all questions have been answered, if the individual is interested in participation and has signed the consent form and HIPAA authorization, they will be given a copy of the consent documents. Completed e-consent documents will be automatically emailed to individuals by REDCap. Consent documents will include the contact phone numbers of the PIs for that site, in the event the participant has subsequent questions or concerns.

### 4.5 Eligibility: Inclusion / Exclusion Criteria

Eligibility is determined by the following inclusion/exclusion criteria:

#### Inclusion Criteria

- Able to provide consent
- ≥ 40 years old
- Persons with or without type 2 diabetes
- Must speak and read English

#### Exclusion Criteria

- Must not be pregnant
- Must not have gestational diabetes
- Must not have Type 1 diabetes

### 4.6 Screening and Enrollment Monitoring

As discussed above, all individuals from the potential recruitment pool will be preloaded into REDCap prior to sending out recruitment letters/emails. Screening questionnaires, dates and modes of contact, e-consent, questionnaire completion, enrollment, in-person visit scheduling, and protocol checklists will be recorded and maintained in REDCap. The screening and enrollment logs will be used to monitor interest and participation rates, and for CRCs to prepare for in-person visits.

### 4.7 Scheduling Participant Visits

Once potential participants have been determined to meet the eligibility criteria, have consented to participate in AI-READI, and have submitted all questionnaires in REDCap, an in-person visit to each site’s clinical research unit will be scheduled at the participant’s convenience. If a person wants to participate but did not complete e-consent, consent will be emailed to the participant beforehand then the consenting process will be conducted at the beginning of the visit and questionnaires completed on site. Appointments will be scheduled within normal working hours (7:30am/8:00am - 5:00pm, M-F); however, some sites may have extended hours into the early evening. Each site will determine when appointments may be scheduled based on their limitations for overnight shipping of biorepository samples or any other scheduling limitations. **Important: Appointments may only be scheduled on Fridays and Saturdays if there is a procedure in place for delivering overnight biorepository samples to FedEx for Monday morning delivery to UAB**. Please see sections 5.13 and 5.14 for more information.

During the initial scheduling phone call and in reminder notifications, the participant will be asked to bring all current prescribed and over-the-counter medications with them to their appointment. If participants choose to not bring in medications, they will be given the option of taking photographs of all medications and/or preparing a written list that they bring with them to their visit. The list must include all medication-related information on the product or prescription label. Participants will also be reminded to drink plenty of fluids (at least 2 cups, 16 oz) before their visit in order to arrive well hydrated for blood and urine collection. CRCs may also suggest that participants wear loose fitting clothing and/or button-down shirts to allow for electrocardiogram (ECG) measurements without changing into a hospital gown.

Leading up to the in-person visit, emails should be sent and/or texts or phone calls made a few days in advance to remind participants of their upcoming appointment. This reminder will also include a request to bring all current prescribed and over-the-counter medications and to arrive well-hydrated in button-down shirts.

If a participant cannot complete the RedCap questionnaires electronically before their visit (or prefer not to), then they will be asked to complete the forms during their visit. As mentioned above, this will lengthen the visit time, so CRCs will encourage the participant to complete the forms at home before their appointment. CRCs will view reports of all surveys for each participant prior to their visit. They will identify any missing information and address those questions with the participant at the start of the visit.

Free parking is provided at each site within a short walking distance of each CRU. Each site will implement their usual procedures for arranging for free parking for clinical research participants.

### 4.8 Participant Withdrawal

Participants may withdraw consent at any time and cease study participation. However, any data that has been shared or used up to that point will stay in the study. This is clearly communicated in the consent document. CRCs will immediately complete the study disposition form in REDCap to indicate the date of withdrawal and provide any additional information, if available.

### 4.9 Participant Compensation for Completion

Participants will be compensated $200 for completing the study and for returning the three monitoring devices (Garmin watch, environmental sensor, continuous glucose monitor transmitter). Compensation will be made using an institutionally-approved gift card. If a participant fails to return any of the monitoring devices (monitored by FedEx tracking number), the gift card will not be provided. If a participant initiates the shipment, but the devices are lost in transit, they will still be compensated.

Participants are consenting to participate in all elements of the protocol. The expectation is that they must attempt to participate in all elements in order to receive the stipend. Participating in the 10-day at-home monitoring period for continuous glucose, fitness/activity (Garmin watch), and environmental sensing, and the subsequent return of the three devices is mandatory for receiving the stipend. If a participant attempts to participate in all elements, but is unable to complete an element because of discomfort, physical inability, or other, but returns the devices, they will still receive the stipend. It will be rare, not common, for the following items to be incomplete during the protocol: vision testing, retinal imaging, ECG, blood specimen collection, cognitive testing, and questionnaires. If they are incomplete, the participant will still receive the stipend.

### 4.10 Missed Visits

In the event that a participant misses a scheduled visit, the CRC will contact the participant by phone or email and attempt to reschedule the visit. If a participant misses a scheduled visit two times, the CRC may consider not to attempt to re-schedule the visit again. Each site can implement this specific procedure as appropriate. If the participant has completed the consent and some questionnaires, but does not complete an in-person visit, the questionnaire data may still be released in the AI-READI final dataset. Please see Section 4.11 below for assigning Participant IDs under these circumstances.

### 4.11 Assigning Randomized Participant IDs

Unique Participant IDs must be created in REDCap once an individual has passed the screening process, has signed the consent document, and has completed questionnaires. **Participant IDs will be assigned when the participant arrives for their in-person visit and not before**. This will produce sequential Participant IDs, chronologically, and will not allow for gaps in ID#s if participants fail to show up for their visits. To assign a participant ID, CRCs will select the “study enrollment” form under the participant’s record number and add a new number to the “Participant Study ID” field. Please be sure to add the next number in the series for your site. **Important: Each Data site has been assigned a range of numbers they will use. CRCs will assign the next available number in the range for their site. REDCap will not allow duplications in Participant Study ID**.

Once a Participant ID has been generated for an individual, that ID must be used on all data collection elements including e-forms, hard-copy forms, retinal imaging files, ECG files, CGM files, physical activity tracker files, environmental sensor files, visit checklists, clinical lab samples/reports, and biorepository samples. **Personal identifiers must be absent from all collected data and the AI-READI dataset**. Instructions on how to enter other participant information into devices may be found under the procedures for each device.

**For participants who sign the consent document and complete questionnaires, but never complete an in-person visit**: survey data will be preserved and eventually transferred to fairhub.io. Only data associated with a Participant ID is transferred to the dataset. At the end of the data collection period of the project, Data Managers will assign Participant IDs to individuals who have completed some questionnaires, but never completed their in-person visit.

### 4.12 Monitoring Recruitment Numbers

All recruitment data on study progress and completeness of data will be monitored. This will inform the Data team of any needed adjustments to planned processes for recruitment and retention should they not be as successful as planned. During the recruitment process, REDCap will be used to monitor recruitment in general and by site. The UAB MPIs will monitor all recruitment and provide alerts to any site where recruitment is lagging (defined as <8 participants per week on average) with recommendations to ameliorate this problem. UAB Data MPIs will also monitor recruitment and adjust sampling to balance enrollment for the four categories of diabetes health.

## 5. Clinical Visit Schedule and Procedures

The standard operating procedures (SOPs) for collecting data in the AI-READI study protocol are described in this chapter and listed in Table 2 below. The protocol has been standardized and harmonized between the 3 data collection sites. The protocol will only move forward after informed consent is completed.

**Table 2.** Clinical Visit Schedule.

|  | **Element (Tasks are completed by the CRC unless otherwise noted)** | **Time (min)** | **Procedure** | **Data Form** |
| --- | --- | --- | --- | --- |
|  | CRC prepares CRU rooms for visit (turn on and calibrate the EVA device, sanitize, etc.) and reviews REDCap reports for missing content. |  | 5.2, 5.10.3 |  |
|  | *Do informed consent if not obtained electronically<br>*Participant completes Questionnaires if they did not give e-consent and/or were not able to complete questionnaires electronically. Missing items addressed. | 20-30 | 4.3, 4.4, 5.5 | All questionnaires in Section 5.5 (except Medications List) |
| **1** | Driving Record form completed | 2 | 5.4 |  |
| **3** | Blood and urine collection; if the participant arrives at the visit and has not eaten since the night before, the blood and urine collection should be done first and then the participant can receive a snack. | 15 | 5.13.1, 5.13.2, 5.13.3, 5.13.4 | Lab requisition form (site-specific) |
| **3** | Obtain medications list | 10 | 5.5 | Medications forms |
| **4** | Obtain height, weight, waist circumference, hip circumference, blood pressure, heart rate. | 10 | 5.6 | Physical Assessment form |
| **5** | Autorefracton and BCVA (photopic and mesopic) | 10 | 5.10.1<br>5.10.2<br>5.10.3<br>5.10.4 | BCVA data sheet |
| **6** | Contrast sensitivity testing (photopic and mesopic) | 10 | 5.10.5 | Photopic Mars data sheet<br>Mesopic Mars data sheet |
| **7** | 12-lead ECG (monofilament test during rest period, see below) | 20 | 5.7 |  |
| **8** | Monofilament test for peripheral neuropathy, performed during the ECG rest period | - | 5.8 | Monofilament test data sheet |
| **9** | Cognitive assessment | 15 | 5.9 | MoCA Duo App |
| 10 | Optomed Fundus prior to dilation, then Pupillary dilation: This should be done before other retinal imaging. | 10 | 5.12.1 | Retinal Imaging Checklist |
|  | CRC demonstrates use of physical activity wearable monitor, environmental sensor, and CGM. CRC sets up Dexcom G6 (CGM) (inserts the sensor) and Garmin watch. *This step of the protocol may occur during the eye dilation period | 20 | 5.11.1<br>5.11.2<br>5.11.3 |  |
| 11 | Dilation and Retinal Imaging | 90 | 5.12.2<br>5.12.3 | Retinal Imaging Checklist |
| 13 | Review protocol checklist; notify participant of date to return fitness tracker, environmental sensor, and CGM and emphasize that these items must be returned; provide return box and instructions; log FedEx tracking number in REDCap; validate parking; present the Participant Data Card; participant is finished | 10 | 5.11.1<br>5.11.4 | Checklist on REDCap<br>Enter FedEx return label (tracking number) into REDCap |
| 14 | CRCs process blood samples for clinical testing. CBC test to be processed at local lab for each site. Other samples are frozen and batch delivered (quarterly) to UW testing lab. |  | 5.14,<br>5.14.1,<br>5.14.2,<br>5.14.3 | Laboratory services requisition |
| 15 | CRCs centrifuge blood in CPT tubes to prepare for daily FedEx shipment to the UAB Biorepository laboratory. PaxGene tubes are included in shipment |  | 5.14.3,<br>5.14.4<br>5.14.7 |  |
| 16 | CRCs prepare buffy coat, plasma, and serum samples and place in -80°C freezer for storage until batch shipped to the UAB Biorepository lab |  | 5.14.3,<br>5.14.5,<br>5.14.6 |  |
| 17 | CRCs monitor device return in REDCap; provide compensation to participant once devices are received |  | 5.11.5 |  |

\* We anticipate that this situation will be infrequent. Participants will be encouraged to complete questionnaires electronically. In-person consent and questionnaire completion will substantially increase the duration of the visit.

### 5.1 Overview of Schedule and Description of Participant Visits

Table 2 outlines the elements of a participant’s in-person visit. Please note: if e-consent was not obtained prior to arrival, the informed consent process must occur first, before proceeding with Step 1. Because each Data site has different resources available to them, the actual order of elements within the visit may vary; however, some parts of the protocol are sensitive to order. **Important:** prior to a scheduled in-person visit, CRCs should view REDCap reports of completed surveys, making note of any missing data. Attempts to address missing survey information should be performed at the start of the in-person visit.

### 5.2 Sanitation Procedures

Each Data site will have their own institutional requirements for maintaining good sanitation procedures during the in-person evaluation.

### 5.3 Participant Contact Information

While this information is available from the EHR, it is important to confirm information again, for example because of moving, phone number changes, and name changes. Participant name, address, phone number, email address, and preferred mode of contact will be provided electronically by the participant prior to their in-person visit. If the participant does not complete REDCap forms electronically, the CRC may collect their information at the beginning of the in-person visit. Contact information will **not** be included in the AI-READI dataset and will only be used by the site to communicate with the participant about the visits.

### 5.4 Motor Vehicle Collision Reports (“Accident Reports”)

The CRC will ask for the participant’s driver’s license using the following language: “We are interested in driving safety for participants in our study. We would like to obtain information on your driving safety, namely obtaining any accident reports from the state for any collisions you have been involved in. This information will not impact your driving license by the State in any way, and it will not have any implications for your driving status. We are only using the information to improve our understanding of what circumstances facilitate collisions.”

The CRC will complete all information on the “Driving Record” form in REDCap, including their driving habits, driver’s license number, and license issuing state. **The information on the license will not be placed in the AI-READI dataset**. The name of the driver and the DL number will be used to request “accident reports” from each site’s state for each driver. Requests for retrospective accident reports will be done at the end of Year 4, after enrollment is complete.

### 5.5 Questionnaires

These are designed to be self-administered questionnaires. As mentioned previously, many participants will complete them electronically at home. Others will complete them during their visit, although we encourage the electronic submission of questionnaires prior to the visit in order to prevent extending the CRU visit time. CRCs will view REDCap reports of missing survey items prior to a participant’s in-person visit and make a plan for addressing missing answers at the start of the visit.

The instructions at the top of each questionnaire on how to complete each form should be self-explanatory. However, participants will be instructed to contact the CRC if they have questions. For most questionnaires, each question will include an option of “prefer not to say” and/or “don’t know”, which allows the participant to not answer questions they choose not to answer, but still qualify as “complete” in REDCap. Opt-out answers will not be provided for scored, validated surveys that do not already incorporate “prefer not to say” as an option.

#### Initial Screening Questionnaire

This will be used as an initial screening tool for enrollment. The questionnaire asks if an individual is pregnant or has Type 1 diabetes, which are both exclusion criteria for the AI-READI study. Those who are 40 or older, are not pregnant, and do not have Type 1 diabetes will be presented with a series of additional questions on previous diabetes diagnoses, diabetes-related medications/treatment, biological sex assigned at birth, and race/ethnicity. This questionnaire will enable us to properly stratify our enrollment based on recruitment needs and the ongoing monitoring of participation rates.

All questionnaires used in the AI-READI protocol are summarized in Table 3 below:

**Table 3.** Questionnaires used in the AI-READI Protocol.

| Questionnaire | Domain Addressed |
| --- | --- |
| Screening Questionnaire | Screens for eligibility (no type I diabetes or pregnancy). Also asks about diabetes health status, diabetes treatments (lifestyle, medications), biological sex, race and ethnicity |
| Demographic information | Date of birth, marital status |
| Center for Epidemiological Studies Depression Scale (CES-D) – 10 | Validated, 10-item instrument that screens for depressive symptoms |
| Problem Areas in Diabetes (PAID-5) | Validated, 5-item instrument assessing diabetes-associated injuries, lifestyle changes, and medical care |
| Diabetes Score (Self-Care) | A 10-item instrument focusing on diabetes self-management by querying dietary, exercise, healthcare, and foot care habits |
| Dietary Assessment | A 9-item diet questionnaire; asks basic questions about food and drink habits over the past few months |
| Visual Impairment and Eye Care Access Questionnaire | A 6-item instrument that asks about difficulties with vision and recent eye care |
| Smoking, alcohol use, vaping, and marijuana use* | Asks about the history for each of these behaviors (past and present use) |
| General health | Asks about health history using this question: "Has a doctor or other healthcare professional ever told you that you have/had" followed by a list of chronic health conditions. Responses are yes/no; if yes, may be asked to specify a condition. |
| Social Determinants of Health (SDoH) Surveys from the PhenX Toolkit<br>(https://www.phenxtoolkit.org/). | A chosen set of surveys that ask about social determinants that may impact health and access to healthcare. Included are surveys on food and job insecurity, educational attainment, health insurance coverage, access to healthcare, housing and neighborhood environment, perceptions of discrimination in medical settings, and racial/ethnic discrimination (current and lifetime). |
| Current Medications with RxNorm codes | Asks about all prescription and over-the-counter medications currently used. Includes pills, injections, creams, salves, sprays, eye drops, and dermal patches. For each medication taken, the medication name, type of medication, associated RXNorm code, dosage (mg or other units), quantity taken, and frequency taken will be obtained. |

\*Marijuana use not assessed for participants living in Alabama because it is an illegal substance

### 5.6 Physical Assessment

Height, weight, waist circumference (WC), hip circumference (HC), blood pressure (BP), and heart rate (HR) measurements will be taken using the methods described below. Because obtaining waist circumference (WC) and hip circumference (HC) involve lifting up the shirt to expose the abdomen, this should be done in a location where privacy is ensured.

Directly enter the participant’s data into the REDCap form titled “Physical Assessment” using an iPad or laptop, or record the data on a hardcopy printout of the form for later entry into REDCap.

#### Dietary Questions

Ask the participant the question “How many hours since you last ate?”. Enter the whole number (in hours) into the appropriate REDCap field. **Round down if the answer is not a whole number**.

If the number entered is 6 hours or greater, you will be prompted by REDCap to ask a clarifying question: “Have you had anything to eat or drink in the last six hours, including coffee or tea with milk or sugar or any other drink other than water?”. Select Yes or No option based on the participant’s response.

##### Important note

This question is essential for identifying participants in the dataset who had blood drawn under fasting conditions. If the participant indicates they have not consumed anything other than plain water in the previous 6 hours, arrange to have their blood drawn as soon as possible and do not offer them a snack before their blood is drawn. If they require a snack for physiological reasons, then update the dietary question in REDCap accordingly.

#### Measuring Height

1. Take the patient into the room with the height rod and/or weight scale.
2. Have the participant remove shoes.
3. Raise the headpiece above the patient’s head and have the patient step under the headpiece.
4. Position the patient against the height rod.
5. Ensure that the patient is standing upright with their head straight and level.
6. Push the headpiece down until it rests on the patient’s head.
7. Determine the patient’s height by reading the value on the measuring strip nearest to the line on the indicator.
8. Record the height to the tenth of a centimeter (cm)

#### Weight Scale

(weight measurements may be made on any calibrated scale.)

1. Have the patient step onto the scale. Follow the instructions that are specific for the type of scale being used. This will vary by site. Make sure that the scale is set to kilograms (kg).
2. Record the weight to the tenth of a kg.

#### Waist hip ratio

##### Waist Circumference (WC)

1. This requires a tape measure that provides measurements in cm
2. Have the participant stand with their shoes off with their arms crossed in front of their chest
3. Have the participant pull up their shirt to expose the abdomen
4. Feel for the hip bone (iliac crest). Place a tape measure around their abdominal cavity, just above the hip bones
5. Make sure the tape is horizontal around the waist
6. Keep the tape snug around the waist, but not compressing the skin
7. Take the measurement at the end of a deep breath out.
8. Record the waist circumference to the tenth of a cm

##### Hip Circumference (HC)

1. Have the participant stand with their shoes off with their arms crossed in front of their chest
2. Have the participant pull up their shirt to expose the hip area.
3. Place a tape measure around the widest part of the hips
4. Make sure the tape is horizontal around the hips
5. Keep the tape snug around the hips, but not compressing the skin
6. Take the measurement at the end of a deep breath out.
7. Record the hip circumference to the tenth of a cm

##### Waist Hip Ratio (WHR)

REDCap will perform this calculation automatically, using the following formula:

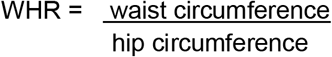

#### Blood Pressure (BP) Measurement Using an Automatic (Oscillometric) Device

1. Clean hands prior to starting the procedure by either washing hands or using hand sanitizer gel. Don on gloves.
2. The participant should be seated in a quiet room, with the back supported and feet flat on the floor. Legs should not be crossed. Have the participant rest in this position for at least 5 minutes before measuring.
3. BP should be measured in the RIGHT arm for consistency. If RIGHT arm is unavailable, then the LEFT arm may be used. Note: if the blood draw was taken on the right arm and the participant expresses discomfort at having their blood pressure taken on the same arm, you may use the left arm for BP and indicate in a REDCap comment.
4. The participants’ midpoint of arm should be at heart level, supported, and uncovered above the cuff. The patient and observer should not speak while the measurement is being taken.
5. Ensure the correct cuff size is being used. The bladder length should be 80%-100% of the circumference of the arm, and the width should be at least 40%
6. Place the deflated cuff with the bottom border about 1 inch above the antecubital fossa (crease of arm). Newer cuffs have an indication line where it should line up with the brachial artery. The artery line indicator should align with the brachial artery. If no line exists, fold the bladder (not the cuff) in half – the line created should be placed over the artery. You should be able to place two fingers snuggly underneath the cuff once it’s placed. If it’s too loose or so tight that two fingers cannot be placed under the cuff, tighten or loosen the cuff appropriately.
7. Press the ON/OFF button on the equipment. Display symbols will appear. Let the subject know that the measurement will begin and ask for them to relax, remain still and not speak.
8. Press the START button. The cuff starts to inflate automatically. Measurement will be displayed when completed and the arm cuff completely deflates.
9. Record the measurement in REDCap or on a data entry form for later entry into REDCap
10. Wait 2 minutes before completing measurement again in the same arm. Make sure item 6 is still correct. Let the subject know the measurement will begin and ask for them to relax, remain still and not speak. Repeat item 8.
11. Record the measurement in REDCap or on a data entry form for later entry into REDCap

#### Heart Rate (HR) Measurement

1. Together with the information on blood pressure, heart rate information will be provided by the equipment.

#### Abnormal Readings

When collecting blood pressure and heart rate, any urgent or emergent findings will specifically be called out that is not known to be usual for the participant. Actionable Health-Related Findings at the time of baseline physical measurements are as follows below. In the event of needing emergent care, patient refusal will be documented according to site-specific policies and procedures. Before proceeding with any of the actions below, ensure to ask the question “Is this normal for you”? If not, proceed with directions below.

#### Systolic Blood Pressure

- Readings of >180 mmHg or <100mmHg (with any symptoms of hemodynamic instability): patient is immediately advised of the need for emergent care, stop visit and recommend patient go to the emergency department.
- Readings of 130-180 mmHg: recommend that patient seek medical care and notify their provider.

##### Diastolic Blood Pressure

- Readings of >120 mmHg or <60 mmHg (with any symptoms of hemodynamic instability): patient is immediately advised of the need for emergent care, stop visit and recommend patient go to the emergency department.
- Readings of 110-120 mmHg: recommend that patient seek medical care and notify their provider.

##### Heart Rate

- <60 or >100 bpm and not known to be usual for the participant (with any symptoms of hemodynamic instability): patient is immediately advised of the need for emergent care, stop visit and recommend patient go to the emergency department.
- <60 or >100 bpm and hypotension (systolic blood pressure of <90) without symptoms of hemodynamic instability: patient is immediately advised of the need for emergent care, stop visit and recommend patient go to the emergency department.
- Asymptomatic heart rate <50 bpm or >120 bpm, not known to be usual: recommend that patient seek medical care and notify their provider.
- Heart rate >100 bmp and irregular pulse unable to be determined via digital cuff: recommend that patient seek medical care and notify their provider (unless confirmed by participant that an irregular heart rhythm is already a known condition)

##### Return of Data Card

Fill out the blood pressure and heart rate sections of the “Participant Data Return Card” (Appendix C). If the participant’s data is abnormal for them, take appropriate action as described above.

### 5.7 Cardiology

A 12-lead electrocardiogram (ECG) is a non-invasive procedure used to ascertain information about the electrophysiology of the heart. All data collection sites will utilize the Philips Pagewriter TC30 Cardiograph (Philips Healthcare, Atlanta, GA) to obtain ECG data from participants. Please note: the monofilament test for peripheral neuropathy will be performed during the rest period of the ECG procedure. Study personnel are responsible for ensuring that the 12-lead ECG required by study protocol is being utilized. Each data site must have available a private room with a reclining chair or bed designated for ECG purposes.

1. Walk the participant to the ECG room and ask them to sit in the reclining chair or lie supine on the bed.
  - Cell phones should not be nearby when performing the ECG
  - Wash hands and gather the following:
  - 12-lead ECG device
  - ECG electrodes
  - Adhesive remover swabs (optional)
  - 2 x 2 gauze pads (optional)
  - Razor (optional)
2. Have the individual lie supine or recline (with feet elevated) for a 10-minute rest period. Important: the participant’s legs must be straight and uncrossed. **During the 10-minute rest period, attach the electrodes (but not the leads) and perform the Monofilament test (section 5.8, below)**.
3. Turn on the ECG device and stand to the side of the subject. Assess whether electrodes may be accurately placed by raising and/or unbuttoning the participant’s shirt. This is the preferred method for privacy and expediency. If this is not possible, ask the subject to remove clothing from the waist up and don a hospital gown with the opening to the front (bra is not to be removed). Close the curtain and door for privacy before changing. *If the participant has worn loose fitting clothing that is easily moved for electrode placement, then they do not need to change into a hospital gown.
4. Press the “ID” button located just above the keyboard. Select “New Patient” and enter the following required information into the fields as indicated:
  a. **First Name**: enter Participant ID#
  b. **Last Name**: enter AIREADI
  c. **ID**: enter AIREADI-Participant ID#
  d. **DOB**: enter 01/01/actual birth year
  e. **Gender:** actual biological sex (this is the only device where default=male doesn’t apply)
  f. **Position**: choose one of 4 options from the pull-down menu
    i. 0 degrees (supine) - if lying flat on a bed
    ii. 30 degrees (reclined) - if propped up in a gurney or the most reclined setting of a reclining chair
    iii. 60 degrees (slight recline) - if slightly reclined in a chair
    iv. 90 degrees (sitting) - if sitting upright
5. Explain the ECG procedures to the subject and ask the subject if they have any questions. If you are unsure of the answer, refer the subject to the Data Manager or PI.
6. If needed, shave the skin where electrodes will be placed to ensure proper adhesion. If you need to shave, use soap and water only, following the manufacturer’s training instructions. **Do NOT clean the skin using alcohol wipes or swabs. This will prevent the electrodes from sticking properly**. It is recommended that you rub the skin gently with gauze to abrade the area prior to electrode placement, but do **not** use alcohol or other substances to clean the skin. Electrodes should be placed and then left alone during the 10-minute rest period before applying the leads. This gives the electrodes time to properly adhere to the skin before applying the weight of the lead wires.
7. Apply the disposable electrodes according to the lead map provided in the Pagewriter TC30 User Manual, starting with the lower legs, lower forearms, then chest area. (https://www.documents.philips.com/assets/Instruction%20for%20Use/20220324/7042d3deea004adb9229ae6200e32ae6.pdf?feed=ifu_docs_feed). The electrode and lead placements are summarized below: **RL** On the right leg, lateral calf muscle OR upper thigh or lower abdomen **LL** In the same location that RL was placed, but on left leg, thigh, or lower abdomen **RA** On the right arm, avoiding thick muscle **LA** Same location that RA was placed, but on left arm **V**_**1**_ In the *fourth* intercostal space (between ribs 4 & 5) just to the *right* of the subject’s sternum (breastbone) **V**_**2**_ In the *fourth* intercostal space (between ribs 4 & 5) just to the *left* of the subject’s sternum **V**_**3**_ Between leads V_2_ and V_4_. **V**_**4**_ In the *fifth* intercostal space (between ribs 5 & 6) in the mid-clavicular line (the imaginary line that extends down from the midpoint of the clavicle). In female subjects, place at the base of the breast. **V**_**5**_ Horizontally even with V_4_, but in the anterior axillary line. (The anterior axillary line is the imaginary line that runs down from the point midway between the middle of the clavicle and the lateral end of the clavicle; the lateral end of the collarbone is the end closer to the arm) In female subjects, place at the base of the breast. **V**_**6**_ Horizontally even with V_4_ and V_5_ in the midaxillary line. (The midaxillary line is the imaginary line that extends down from the middle of the subject’s armpit)
8. **Once the electrodes are placed, perform the Monofilament Test following the instructions in Section 5.8 - Peripheral Neuropathy (below)**.
9. After the 10-minute rest period, connect the electrical leads to the electrodes following the labels on the leads and according to instructions provided in the manual referenced above. Once the leads are connected, electrical activity will be visible on the screen. When you look at the waveform it will be color coded:
  a. Green = good connection; take the ECG
  b. Yellow = moderate noise level (participant legs moving?); poor electrode connection. Electrical interference from another device (make sure phones are put away). Verify that all electrodes are firmly adhered to skin.
  c. Orange = severe artefact or poor electrode connection. One or more limb leads are disconnected (chest leads are probably fine). Patient moving around or possibly dry electrodes.
  d. Red = ECG waveform data cannot be analyzed. Loose electrode connection or bad lead. Electrode has fallen off of the participant; lead displaced.
  e. See the diagram below. You may need to re-position electrodes if you cannot establish strong connections for each of the leads.
10. Touch the “**Map**” button to check for any loose or inoperative electrodes or lead wires. A red “X” mark identifies the location of a bad electrode or lead connection.
11. Verify that the participant’s legs are uncrossed and their arms are lying flat to their side. **Important: make sure the participant understands that they should not move their arms or legs during the next steps. They need to lie very still**. Ask the subject to breathe normally, relax and remain still.
12. After all the leads are connected and showing green in the lead map (the “ECG” button on the lead gang-input will also glow green when ready), acquire the ECG reading by pressing the far left ECG button on the device. Refer to the manufacturer’s operating instructions for more details. If the reading comes back abnormal, readjust the placements of the electrodes to rule out improper electrode placement or poor skin contact. **Two important indicators to look for at the top of the ECG display is: “Baseline Wander” and “Low Voltage Precordial Leads”**. Possible causes of these errors are patient movement and poor electrode contact. Address with the participant and repeat the ECG until these indicators are absent.
13. When a satisfactory ECG is acquired, save the ECG into archives.
14. Remove lead wires and electrodes from the subject. Adhesive swabs or a warm, moist washcloth may be used to remove adhesive gel from skin.

### 5.8 Peripheral Neuropathy

Peripheral neuropathy is characterized as the loss of nerve function in the extremities due to nerve damage. We will use the monofilament test to assess a loss of touch sensation on the foot. We implement this test during the rest period of the ECG procedure. **Data collected during this test will be entered directly into the REDCap form titled “Monofilament” using an iPad or laptop**, or completed on a hardcopy form for later entry into REDCap.

1. Monofilament testing must be carried out in a quiet/relaxed setting to allow the participant to fully participate in the test. Have the participant remove shoes/socks.
2. Clean hands prior to starting the procedure by either washing hands or using hand sanitizer gel. Put on gloves.
3. Explain the procedure to the participant and demonstrate the monofilament test on a gloved hand to show how the test is done. Optional: perform the test on the participant in an area where normal sensation is expected. This allows the participant to experience the feeling before the test is done on their feet.
4. It is important that the participant’s foot be supported (i.e., allow the heel of the foot to rest on a flat, warm surface). The filament should initially be pre-stressed (4-6 perpendicular applications to the dorsum of the first finger). A standard 10g monofilament must be used.
5. Perform the test on each foot separately. Beginning with the RIGHT foot, have the participant close their eyes and hold the monofilament perpendicular to the foot. With a smooth steady motion, apply the filament to the dorsum of the great toe midway between the nail fold and the DIP joint (plantar aspect of first toe). Do not hold the toe directly. The filament is applied perpendicularly and briefly (<1 second) with an even pressure. When the filament bends, the force of 10 grams has been applied.
6. The participant, whose eyes are closed, is asked to verbally respond yes if they feel the filament.
7. Repeat item 5 until 10 touches have been completed. *Use the foot diagram provided to your site to test area A four times, area B three times, and area C three times. With each touch, the filament is moved very slightly but staying in the same area (top of big toe, between the nail and joint). Record the number of touches the participant verbally responds yes to feeling.
8. Eight correct responses out of ten applications is considered normal; one to seven correct responses indicate reduced sensation and no correct answers translates into absent sensation.
9. Repeat item 5-7 on the LEFT foot.
10. **When the test is complete, resume the ECG procedure (section 5.8, above) starting with step #8**.
11. Thoroughly sanitize the monofilament device after each use or discard (if disposable).

### 5.9 Cognition

The Montreal Cognitive Assessment (MoCA) will be administered electronically using an iPad and the MoCA Duo Application (MoCA Cognition, Greenfield Park, Québec, Canada). The total possible score is 30 points.

The MoCA is available in English and Spanish. By interacting with the participant, the CRC will determine which version to use.

#### MoCA Duo Application Instructions

All iPads used for AI-READI data collection must be preloaded with the MoCA Duo app, which is available in the Apple App store. AI-READI has been granted copyright permissions for use of the MoCA Duo app at the three data sites and the subscription is free for all members of the project. Once the MoCA Duo app is downloaded to the iPad, testing may be administered by following the instructions listed below.

##### Procedure

1. **Important!** Be sure that you are logged out of REDCap prior to initiating the MoCA Duo App, because the participant will be performing some tasks directly on the iPad.
2. Log into the MoCA Duo App using your email address and password
3. Click “Add New Subject” or search for an existing subject
4. Enter the following into the appropriate required fields:
  a. **Age Range**: ask the participant their age and select the correct option
  b. **Gender**: enter default of male, regardless of actual biological sex
  c. **Education**: ask the participant their highest level of education and select the appropriate option
  d. **Subject File Number**: enter the Participant ID#
5. Click “Start New Test.” You can also look at previous tests done by that subject.
6. Indicate preferred language and subject’s spoken dialect.
7. CRC will be on one side of the tablet, participant on the other. Instruct the participant on how to draw on screen.
8. Tap the pencil icon to write on the screen, and tap the eraser icon to erase.
9. Administer the pre-test “TEST” to ensure the participant is comfortable taking the test on the tablet.
10. Icons at the bottom of the screen suggest how many times the instructions can be repeated (for the “Test” writing, instructions can be repeated once).
11. Click the red button in the bottom left corner to terminate the test.
12. Slide the blue arrow in the bottom right corner to the right to move on to the next section.
13. If the subject is having difficulty writing on the screen, they can write on paper. You can take a picture of it and upload it on the app.
14. Proceed with the test as you would with the paper version except you will use the verbal instructions provided at the bottom of the screen for each test. The instructions will also indicate how many times you are permitted to repeat verbal instructions. **After Naming (task #4), DO NOT show the screen to the subject going forward**.
15. When finished with the test, select **“Save and Score Later”**
16. **After the participant visit, re-open their test and check the following:**
  a. Quickly look at the drawings. Override the score if the app has incorrectly marked something wrong or didn’t detect a correct drawing.
  b. Quickly verify that there are no typos or misspellings of the animal names, because this will result in an incorrect score (even if participant answers correctly)
  c. For the Fluency tast, eliminate any extra recorded words that don’t apply to the task (unless the participant gets over 11 correct words)
  d. If all looks fine, select **“Save Final Score”**

##### If the MoCA duo app is unavailable, the identical paper version of the test will be used

The instructions below are provided for when use of the paper version is necessary. The paper version of the MoCA Procedure (MoCA Version 8.1 English 2020) may be found at https://mocacognition.com/. The following are instructions for administering the paper version of the MoCA. All instructions may be repeated once.

##### 1. Alternating Trail Making

###### Administration

The examiner instructs the subject: *“Please draw a line going from a number to a letter in ascending order. Begin here* [point to (1)] *and draw a line from 1 then to A then to 2 and so on. End here* [point to (E)].”

###### Scoring

One point is allocated if the subject successfully draws the following pattern:

1- A- 2- B- 3- C- 4- D- 5- E, without drawing any lines that cross. Any error that is not immediately self-corrected (meaning corrected before moving on to the Cube task) earns a score of 0. A point is not allocated if the subject draws a line to connect the end (E) to the beginning (1).

##### 2. Visuoconstructional Skills (Cube)

###### Administration

The examiner gives the following instructions, pointing to the cube: “*Copy this drawing as accurately as you can*.*”*

###### Scoring

One point is allocated for a correctly executed drawing.

- Drawing must be three-dimensional.
- All lines are drawn.
- All lines meet with little or no space.
- No line is added.
- Lines are relatively parallel and their length is similar (rectangular prisms are accepted).
- The cube’s orientation in space must be preserved.

A point is not assigned if any of the above criteria is not met.

##### 3. Visuoconstructional Skills (Clock)

###### Administration

The examiner must ensure that the subject does not look at his/her watch while performing the task and that no clocks are in sight. The examiner indicates the appropriate space and gives the following instructions: *“Draw a clock. Put in all the numbers and set the time to 10 past 11*.*”*

###### Scoring

One point is allocated for each of the following three criteria:

- Contour (1 pt.): the clock contour must be drawn (either a circle or a square). Only minor distortions are acceptable (e.g., slight imperfection on closing the circle). If the numbers are arranged in a circular manner but the contour is not drawn the contour is scored as incorrect.
- Numbers (1 pt.): all clock numbers must be present with no additional numbers. Numbers must be in the correct order, upright and placed in the approximate quadrants on the clock face. Roman numerals are acceptable. The numbers must be arranged in a circular manner (even if the contour is a square). All numbers must either be placed inside or outside the clock contour. If the subject places some numbers inside the clock contour and some outside the clock contour, (s)he does not receive a point for Numbers.
- Hands (1 pt.): there must be two hands jointly indicating the correct time. The hour hand must be clearly shorter than the minute hand. Hands must be centered within the clock face with their junction close to the clock center.

##### 4. Naming

###### Administration

Beginning on the left, the examiner points to each figure and says: *“Tell me the name of this animal*.*”*

###### Scoring

One point is given for each of the following responses: (1) lion (2) rhinoceros or rhino (3) camel or dromedary.

##### 5. Memory

###### Administration

The examiner reads a list of five words at a rate of one per second, giving the following instructions: *“This is a memory test. I am going to read a list of words that you will have to remember now and later on. Listen carefully. When I am through, tell me as many words as you can remember. It doesn’t matter in what order you say them*.*”* The examiner marks a check in the allocated space for each word the subject produces on this first trial. The examiner may not correct the subject if (s)he recalls a deformed word or a word that sounds like the target word. When the subject indicates that (s)he has finished (has recalled all words), or can recall no more words, the examiner reads the list a second time with the following instructions: *“I am going to read the same list for a second time. Try to remember and tell me as many words as you can, including words you said the first time*.*”* The examiner puts a check in the allocated space for each word the subject recalls on the second trial. At the end of the second trial, the examiner informs the subject that (s)he will be asked to recall these words again by saying: *“I will ask you to recall those words again at the end of the test*.*”*

###### Scoring

No points are given for Trials One and Two.

##### 6. Attention

###### Forward Digit Span: Administration

The examiner gives the following instructions: “*I am going to say some numbers and when I am through, repeat them to me exactly as I said them*.” The examiner reads the five number sequence at a rate of one digit per second.

###### Backward Digit Span: Administration

The examiner gives the following instructions: “*Now I am going to say some more numbers, but when I am through you must repeat them to me in the backward order*.” The examiner reads the three number sequence at a rate of one digit per second. If the subject repeats the sequence in the forward order, the examiner may not ask the subject to repeat the sequence in backward order at this point.

###### Scoring

One point is allocated for each sequence correctly repeated (N.B.: the correct response for the backward trial is 2-4-7).

###### Vigilance: Administration

The examiner reads the list of letters at a rate of one per second, after giving the following instructions: “*I am going to read a sequence of letters. Every time I say the letter A, tap your hand once. If I say a different letter, do not tap your hand*.”

###### Scoring

One point is allocated if there is zero to one error (an error is a tap on a wrong letter or a failure to tap on letter A).

###### Serial 7s: Administration

The examiner gives the following instructions: *“Now, I will ask you to count by subtracting 7 from 100, and then, keep subtracting 7 from your answer until I tell you to stop*.*”* The subject must perform a mental calculation, therefore, (s)he may not use his/her fingers nor a pencil and paper to execute the task. The examiner may not repeat the subject’s answers. If the subject asks what her/his last given answer was or what number (s)he must subtract from his/her answer, the examiner responds by repeating the instructions if not already done so.

###### Scoring

This item is scored out of 3 points. Give no (0) points for no correct subtractions, 1 point for one correct subtraction, 2 points for two or three correct subtractions, and 3 points if the subject successfully makes four or five correct subtractions. Each subtraction is evaluated independently; that is, if the subject responds with an incorrect number but continues to correctly subtract 7 from it, each correct subtraction is counted. For example, a subject may respond “92 – 85 – 78 – 71 – 64” where the “92” is incorrect, but all subsequent numbers are subtracted correctly. This is one error and the task would be given a score of 3.

##### 7. Sentence repetition

###### Administration

The examiner gives the following instructions: *“I am going to read you a sentence. Repeat it after me, exactly as I say it* [pause]: ***I only know that John is the one to help today***.*”* Following the response, say: *“Now I am going to read you another sentence. Repeat it after me, exactly as I say it* [pause]: ***The cat always hid under the couch when dogs were in the room***.*”*

###### Scoring

One point is allocated for each sentence correctly repeated. Repetitions must be exact. Be alert for omissions (e.g., omitting “only”), substitutions/additions (e.g., substituting “only” for “always”), grammar errors/altering plurals (e.g. “hides” for “hid”), etc.

##### 8. Verbal fluency

###### Administration

The examiner gives the following instructions: *“Now, I want you to tell me as many words as you can think of that begin with the letter F. I will tell you to stop after one minute. Proper nouns, numbers, and different forms of a verb are not permitted. Are you ready?* [Pause] [Time for 60 sec.] *Stop*.*”* If the subject names two consecutive words that begin with another letter of the alphabet, the examiner repeats the target letter if the instructions have not yet been repeated.

###### Scoring

One point is allocated if the subject generates 11 words or more in 60 seconds. The examiner records the subject’s responses in the margins or on the back of the test sheet.

##### 9. Abstraction

###### Administration

The examiner asks the subject to explain what each pair of words has in common, starting with the example: *“I will give you two words and I would like you to tell me to what category they belong to* [pause]: *an orange and a banana*.” If the subject responds correctly the examiner replies: *‘‘Yes, both items are part of the category Fruits*.*’’* If the subject answers in a concrete manner, the examiner gives one additional prompt: *“Tell me another category* to *which these items belong to*.*”* If the subject does not give the appropriate response *(fruits)*, the examiner says: *“Yes, and they also both belong to the category Fruits*.*”* No additional instructions or clarifications are given. After the practice trial, the examiner says: *“Now, a train and a bicycle*.*”* Following the response, the examiner administers the second trial by saying: *“Now, a ruler and a watch*.*”* A prompt (one for the entire abstraction section) may be given if none was used during the example.

###### Scoring

Only the last two pairs are scored. One point is given for each pair correctly answered. The following responses are acceptable:

- train-bicycle = means of transportation, means of travelling, you take trips in both
- ruler-watch = measuring instruments, used to measure

The following responses are **not** acceptable:

- train-bicycle = they have wheels
- ruler-watch = they have numbers

##### 10. Delayed recall

###### Administration

The examiner gives the following instructions: *“I read some words to you earlier, which I asked you to remember. Tell me as many of those words as you can remember*.*”* The examiner makes a check mark (√) for each of the words correctly recalled spontaneously without any cues, in the allocated space.

###### Scoring

One point is allocated for each word recalled freely without any cues.

#### Memory index score (MIS)

##### Administration

Following the delayed free recall trial, the examiner provides a category (semantic) cue for each word the subject was unable to recall. Example: *‘‘I will give you some hints to see if it helps you remember the words, the first word was a body part*.*’’* If the subject is unable to recall the word with the category cue, the examiner provides him/her with a multiple choice cue. Example: *“Which of the following words do you think it was, NOSE, FACE, or HAND?”* All non-recalled words are prompted in this manner. The examiner identifies the words the subject was able to recall with the help of a cue (category or multiple-choice) by placing a check mark (√) in the appropriate space. The cues for each word are presented below:

| Target Word | Category Cue | Multiple Choice |
| --- | --- | --- |
| FACE | body part | nose, face, hand (shoulder, leg) |
| VELVET | type of fabric | denim, velvet, cotton (nylon, silk) |
| CHURCH | type of building | church, school, hospital (library, store) |
| DAISY | type of flower | rose, daisy, tulip (lily, daffodil) |
| RED | color | red, blue, green (yellow, purple) |

\* The words in parentheses are to be used if the subject mentions one or two of the multiple choice responses during the category cuing.

| MIS scoring |  |  |  | Total |
| --- | --- | --- | --- | --- |
| Number of words recalled spontaneously | ... | multiplied by | 3 | ... |
| Number of words recalled with a category cue | ... | multiplied by | 2 | ... |
| Number of words recalled with a multiple choice cue | ... | multiplied by | 1 | ... |
| Total MIS (add all points) |  |  |  | ---/15 |

##### 11. Orientation

###### Administration

The examiner gives the following instructions: “*Tell me today’s date*.” If the subject does not give a complete answer, the examiner prompts accordingly by saying: *“Tell me the [year, month, exact date, and day of the week]*.*”* Then the examiner says: *“Now, tell me the name of this place, and which city it is in*.*”*

###### Scoring

One point is allocated for each item correctly answered. The date and place (name of hospital, clinic, office) must be exact. No points are allocated if the subject makes an error of one day for the day and date.

###### TOTAL SCORE

Sum all subscores listed on the right-hand side. Add one point for subject who has 12 years or fewer of formal education, for a possible maximum of 30 points. A final total score of 26 and above is considered normal.

###### Data Manager

Scan the graded test and share a copy with the Data manager at UAB via Sharefile.

### 5.10 Vision Testing

The following sections will explain how to perform the vision testing procedures proposed in AI-READI.

#### 5.10.1 Lensometer

Participants will be asked to wear their prescribed glasses to the visit. If their reading glasses are separate from their distance glasses, they should be asked to bring both types of glasses. Eyeglass prescription strength will be determined using a lensometer. Participants wearing contact lenses may be asked to remove them to perform the visual testing.

##### Operating the Lensometer

This device is used to determine the prescription of the participant’s eyeglasses.

Instructions below are for the NIDEK LM-600PUV lensometer (NIDEK Co., LTD, Gamagori, Aichi, Japan):

1. Turn the lensometer on by using the switch that is on the left side of the device in the back.
2. Press the L-selection button or R-selection button to specify which lens is being measured.
3. Place the chosen lens on the reader. Measure the top portion of the lens first (usually used for distance).
4. Move the lens around on the reader until the circle on the screen is centered. Once it’s aligned, a cross will appear on the screen. While keeping the glasses in place, press the blue button beneath the lens reader to record the lens measurement.
5. If the participants’ glasses have a visible bifocal, slide the glasses down so that the reader is focused on the bifocal at the bottom of the lens.
6. Move the bifocal lens around on the reader to center it. Once the cross appears, hold the glasses in place and press the blue read button.
7. Press the L-selection or R-selection button to repeat on the other lens. When you finish recording the last measurement, press the print button.
8. Write the Participant ID on the printed results.

Measuring Glasses that are called progressive lens which have a hidden bifocal:

1. Follow the same steps as above in order to get a reading of both the right and left lens, leaving out the steps that are used to capture a reading of the bifocal.
2. Print out the reading that is captured for the top lens and write notate the Participant ID#
3. In order to find out the measure of a bifocal on a progressive lens you must locate a small number that is located on the front of the lens at the bottom toward the side as if you were going towards their ear.
4. This particular number could be on either the right or left lens and may be more easily seen by holding the glasses up to a bright light, front side facing you. The number is very small and sometimes looks as if it is engraved into the lens. The number may appear as +25, which means that it is +2.50 or +27, which means +2.75.
5. Make sure to write this down on your print out for both the right and the left lens; the readings for them will be the same for ex. Add +2.50 for both right and left lens.

Measuring glasses that are considered readers and are not a progressive lens nor do they have a visible bifocal:

1. For glasses that do not have a hidden or visible bifocal, follow steps 4-8 (in first section) and print out the results. Mark the Participant ID# on the printout.

Measuring glasses that are considered to be trifocals (this type of lens has a top lens and two visible lenses at the bottom that are separated by a line:

1. For trifocals, follow steps 4-8 (from first section). You do not have to measure the middle section of the lens; just slide the lens down so that you can measure the bottom portion
2. Measure trifocals the same way you would a regular pair of bifocal glasses
3. Print out the reading and write down the Participant ID#

The lensometer is NOT USED if participant wears contact lenses

Clean the glasses for the participant with the lens paper and lens cleaner in the black tray that is in front of you

#### 5.10.2 Autorefraction

Autorefraction provides the spherical and cylindrical corrections with axis for the right eye and the left eye. The power of lenses in refraction are expressed in diopters. Participants may wear distance spectacles. Some spectacle prescriptions may be current, others may not be current. Some may not have spectacles but will need distance spectacles. **We are interested in measuring best-corrected visual acuity**. Therefore, we will do autorefraction first before we measure visual acuity. Autorefraction procedures are located after the trial frame assembly instructions below. **Autorefraction data will be entered into the REDCap form titled “BCVA”**.

All three data collection sites will use the Topcon KR-800 Auto Kerato-Refractometer (Topcon Healthcare, Inc., La Jolla, CA).

1. Powering on the device
  - Standing in the operator position, there is a toggle switch on the right side of the device. Turn it to the “I” position.
2. Calibration
  - Calibration of this device requires manufacturer maintenance. No operator calibration is possible or necessary.
3. Testing
  - The device defaults to measurement mode when turned on. To enter a patient ID use the touch screen and tap “ID” here and tap “OK” when complete to return to the measurement screen.
  - Begin testing with the right eye. Be sure that “RK” is chosen at the lower left of the capture screen to ensure you are measuring both refraction and K’s.
  - Align the subject’s eye within this target using the joystick for up and down alignment and in and out for focus.
  - Once the eye is aligned and within focus ask the subject to blink several times to ensure an even tear layer spread.
  - Upon occasion it is difficult to get the eye into clear focus, especially in the case of dry eyes. It may be helpful at this point to add artificial tears and avoiding gel tears.
  - Next, press the button on top of the joystick to capture your measurements.
4. Printing results
  - From the touch screen tap the printer icon on the touch screen and this will print your results. These results can then be transferred to the subject record.
5. Entering autorefraction results in REDCap: **if a participant has a spherical reading for an eye, but the autorefractor printout is blank for cylinder and axis (for that eye) - enter 0 for cyl and 0 axis in REDCap**. A blank cyl and axis indicate no astigmatism and a spherical eye, but for the sake of data completeness, this must be indicated by using zeros in REDCap.

#### 5.10.3. Using Autorefraction Results to Obtain Best Corrected Visual Acuity (BCVA)

##### For those who have distance glasses

CRCs will assemble trial frames based on differences in measurement values by the autorefractor and lensometer. This is based only on the sphere measurement. If the difference in spherical dioptric power (from the autorefractor readings) is equal to or greater than 0.50 D (in either direction) compared to their own spectacles (from the lensometer readings), please assemble the prescription in a trial frame based on the autorefractor readings. Autorefractors are not the same as subjective refractions. **At times participants insist that their own spectacles give a clearer image than an autorefraction. In those situations, have the participant use their own spectacles**.

Assembling the lenses into a trial frame: The spherical lens can be mounted at any axis since it is spherical. For the cylindrical lens, mount the cylinder lens according to the axis of the cylinder. Trial frames have axis demarcations on the frame so it is easy to position the lens. Once the lenses have been mounted in the trial frame, adjust the trial frame so it is comfortable for the participant to wear. For optimal optical clarity, insert the least number of lenses possible to create the desired power. The highest powered lens (SPH-sphere) should be placed closest to the cornea (in the back lens clamp). You will refer to the Sphere measurement from the printed receipt from the lensometer and/or autorefractor. Sphere measurements can be positive or negative, so make sure you choose the correct lens.

The CYL (cylinder) lens should be placed in the front clamp. It will always have a positive value. The cylinder lenses will have tick lines on them. Adjust the tick lines using the twist mechanism on the side to the correct axis degree.

When using a mesopic filter or blackout lens on the trial frames, put it on the front lens clamp in front of the cylinder lens.

##### For those who do not have distance glasses

Do the autorefraction measurements. If the dioptric power is ≤ 0.50D, do not use a trial frame refraction. If the dioptric power is > 0.50D, then assemble the prescription in a trial frame based on the autorefractor readings. Ask the participant to wear the trial frames and compare it to their wearing no glasses. If the participant says the trial frames give clearer vision than with no glasses, then use the trial frames. **If clearer vision is obtained without glasses (vs. with the trial frames), then use no glasses**.

#### 5.10.4 Best Corrected Visual Acuity (BCVA) Under Photopic and Mesopic Conditions

We will use an M&S Technologies Clinical Trial Suite EVA-*e*-ETDRS device (M&S Technologies, Niles, IL) to test visual acuity. The test is administered at a distance of 3 meters from a touch-screen monitor that is 12×20 inches. Participants will read letters from the screen. **All data will be entered into the REDCap form titled “BCVA”**.

**Photopic** Conditions: No neutral density filters are used. A general occluder will be used for photopic testing. The participant wears their own **prescription** spectacles or trial frames (see above under autorefraction).

For **Mesopic** conditions, a neutral density (ND) filter will be used. The ND filter will either be a lens added to trial frames to reduce incoming light on the tested eye, OR a handheld occluder with a neutral density filter (which we will designate as “ND-occluder) over the glasses. The ND-occluder is different from a standard occluder and is used only for vision testing under mesopic conditions. Please use the following instructions for care and use of the occluders and neutral density filters.

**Low Luminance Visual Acuity using the ND-Occluder, Setup and Use:** When using the ND-occluder for low luminance visual acuity (LLVA), remember these tips for gaining the best results across all sites.

1. Make sure there is no source of ambient or extraneous light coming from ***behind*** the subject when measuring LLVA. This is important because the 2.0 ND filter within the occluder is a glass substrate and will reflect any light coming from behind the subject and could potentially impact the results of the measurement.
2. When using your ND-occluder for measurements of LLVA, be sure to inform your subject to mindfully be aware of the surface of the ND filter when using glasses during the measurement. It is important that the subject’s glasses or frames do not inadvertently rub against or negatively impact the surface. When administering the LLVA test to the subject, sit off to the side where you can “see” if the subject is aligned properly using the ND-occluder and to ensure that any glasses they may be wearing are not impinging upon the ND filter.
3. Be aware that the most vulnerable side of the ND lens occurs when measuring the LLVA of the OS. This is because the lens is positioned on the ND-occluder slightly more posteriorly than when measuring the LLVA of the OD. This places the ND lens slightly closer to the subject’s glasses and/or eyelashes, etc. and could be more subject to damage or debris.
4. When cleaning the surface of the 2.0 ND filter, be sure to only use a lens cloth (like the one that you received with the occluder) that will not scratch or cause damage to the surface. The microfiber lens cloth is made from a polyester base fabric that will not affect the surface of the ND lens. Never use a Kleenex type tissue to clean the surface of the ND filter because it is made from a wood cellulose pulp that can damage the surface of the ND filter at a microscopic level and over time will erode the coating on the lens.
5. In the event that your ND-occluder becomes damaged or ineffective to the extent that it needs replacing, UAB can provide a replacement ND-occluder made to the same standards.

#### EVA device instructions

Please DO NOT turn on the tablet until the EVA device has been running **10 MINUTES**

1. Powering ON the System
  - Press the power button located on the lower left side of the monitor.
  - When the Open Study Form box appears, press **DRCR**
  - Select **Start Selected Study**
  - Wait at least 10 minutes before you turn on the tablet. After waiting ten minutes, turn on tablet by pressing the BIG middle button for 5 seconds
  - When the blue screen appears on the tablet use your finger to Swipe up on the screen to unlock the tablet
  - On the tablet launch the CT Controller 2.6 app
  - Select Start on EVA Monitor
  - If Date and Time are correct Select Continue on the Monitor
2. Calibration Process (must be performed each morning, prior to the first scheduled visit)
  - Press the Power Button on the LIGHT METER (located in the black back under the monitor)
  - Take the orange cap off and the screen should read x1 Lux
  - Press the Gray button by the word “Range” on the light meter
  - The screen on the light meter should display a 3-digit number
  - Turn off all lights in the room
  - Hold sensor on light meter with white globe facing the display’s white sequence box so that the sensor is nearly touching the display about 1 to 2 cm away
  - The light meter should measure between 138-140 Lux before continuing
  - If you are measuring too high or low, use the buttons on the display screen to make it dimmer or brighter until it measures 138-140 Lux.
  - Once you measure in the right range, press the NEXT button on the EVA Monitor
  - Select the POWER button on LIGHT METER to turn off, place the orange cap back on the sensor and place it back in the black box
  - Next measure the “E” and type 10 in front of the zero in the height and width box so that 100mm is displayed and Click Next
3. Photopic EVA Testing: Set-up Using the EVA Monitor **Testing must be performed undilated**. The participant will view the screen using a **general occluder**.
  - Select “Non-Study” on the monitor
  - Enter AIREADI-Participant ID#
  - Tech ID will be your specific Interviewer Code.
  - Test Selection will be eETDRS (this is the default setting)
  - EYE-Start with the **right eye** being tested first. Select Monocular to test both eyes-right eye and then left eye sequentially.
  - From the Autorefraction, determine if the person is using their own spectacles or trial frames. Prepare the trial frames before the participant is positioned for testing.
  - Bring the Participant into the room
  - Turn lights off according to site instructions
  - **Important:** Use a general occluder to occlude the untested eye
  - Once you are ready to begin, select Start on the Monitor
  - On the tablet, touch “click for EVA”. Once you click, the test starts immediately!
  - When the test is complete, the results will appear on the EVA Monitor. Enter the test results into the REDCap BCVA form.
  - Select “Click for EVA to start the left eye”
  - Enter the results once the test is complete
  - Click “Finish” on tablet
  - Summary of results will appear
  - Click DONE on EVA Monitor located on the top left side of the monitor
  - For low luminance/ mesopic testing, follow the same procedures (exceptions noted below).
4. Mesopic (Low Luminance) EVA Testing: Starting with the right eye **Testing must be performed undilated**. The participant will view the screen using the ND-occluder
  - Place the ND filter so it is over the right eye (left eye is occluded)
  - On the monitor Select Non-Study
  - Select Find Patient to locate the Participant ID
  - Tech ID will be your Interviewer ID.
  - Test Selection will be eETDRS-this is the default setting
  - EYE-select right. Select Monocular to test both eyes.
  - Once you are ready to begin select Start on the Monitor
  - Select “click for EVA” on tablet-once you click this, the test starts immediately!
  - When test is complete, the results will appear on the EVA Monitor; write them down
  - Then, click Finish on tablet
  - Summary of results will appear
  - Click DONE on EVA Monitor located on the top left side of the monitor
  - Follow instructions test the Left eye for LLVA (right eye is occluded)
5. Turning off the device: (leave on if you have multiple visits scheduled for the day and turn off after the last visit)
  - Click BACK TO MAIN button on the EVA monitor
  - Click the EXIT button
  - Double Tap the red Shutdown Button located on top right corner of the EVA Monitor
  - Select OK to shutdown monitor
  - Now turn the tablet off by Pressing the white triangle button at bottom of the tablet screen
  - Select YES for Do you wish to exit application
  - Press and hold the Big Middle button on the tablet
  - The tablet will prompt to POWER OFF, select POWER OFF
6. Safeguards to reduce bias and ensure standardization across sites: Visual acuity examiner instructions to the participant are as follows:
  1. The participant should be told that there are only letters and no numbers and that each letter is “bracketed” by lines on all four sides.
  2. For participants with poor central vision, it may be suggested that the participant fixate eccentrically or turn or move his/her head to improve visual acuity. If the participant employs these maneuvers, care must be taken to ensure that the fellow eye remains covered and that the participant maintains the correct 3 meter distance from the screen.
  3. When the participant cannot read a letter, he/she is told to guess. If the participant states that a letter is one of two letters, then he/she is asked to choose only one letter and, if necessary, to guess.
  4. When the participant gives one response but then gives a second response before the first response has been finalized (i.e., before the examiner has verified the response as correct or incorrect and before the letter presentation on the EVA screen changes), the participant should be asked if that is his/her final answer. If the participant equivocates, the participant is asked to choose one letter. Once the examiner has verified the response and the letter presentation has changed on the EVA, no changes can be made in the participant’s response.

If experiencing issues with the screen luminance call M&S Technologies Tech Support at +1-847-763-0500 for assistance.

#### 5.10.5 Contrast Sensitivity under Photopic and Mesopic Conditions

This contrast sensitivity test measures how much contrast one needs to read a letter. Please see the instructions below for testing contrast sensitivity using the Mars Letter Contrast Sensitivity Test (Mars Perceptrix Corporation, Chappaqua, NY). **All data will be entered on the REDCap forms titled “Photopic Mars” or “Mesopic Mars”**.

##### Testing must be performed undilated

**Administering the Mars Letter Contrast Sensitivity Test – Overview:**

1. **Print out hardcopy forms for Photopic Mars and Mesopic Mars tests (**available at https://marsperceptrix.com/downloads/). It is recommended that CRCS use the hardcopy forms to manually record incorrect responses and the final correct letter. While this can be done in REDCap, CRCs may find it difficult to keep up with the participant if using the REDCap forms. **Important: the Mars card order has been standardized for all sites. Please use the card order for each eye as indicated in the REDCap forms**. The form order to be used is:
  a. Photopic, right eye (OD) = use Form 1 & general occluder
  b. Photopic, left eye (OS) = use Form 2 & general occluder
  c. Mesopic, right eye (OD) = use Form 3 & ND-occluder
  d. Mesopic, left eye (OS) = use Form 1 & ND-occluder
2. **Illumination:** For best results, the chart should be illuminated uniformly, with an optimal luminance in the chart’s white back-ground of 85 cd/m^2^. The chart’s small size facilitates this, and the lamp on a standard ophthalmic equipment stand will generally provide sufficient and sufficiently uniform illumination. Luminance should be at least 60 and less than 120 cd/m2 in all white areas of the chart. Luminance is best checked with a photometer. However, if one is not available, an inexpensive incident light meter can be used; illuminance should be in the range 189 to 377 lux, and optimally 267 lux. Testing should not be conducted through any coatings, laminations, or coverings on the chart, even if these are transparent or translucent.
3. **Viewing distance and correction:** The patient’s viewing distance to the chart is by design 50 cm (20 inches) but may range from the standard near refraction distance of 40 cm (15.75 inches) to 59 cm (23 inches). Patients should wear their appropriate near correction: reading glasses/ trial frames with an add of +2.00 D, and an occluder or patch on the untested eye. If a participant does not typically use reading glasses, they do not have to use them for the test if they prefer not to. The test is quite tolerant of small refractive errors since the letters are large (20/480 equivalent at 50 cm). **Testing should be performed with the eyes undilated**. For patients with very low visual acuity who cannot easily read the highest contrast letters, test distance may be shortened to 25 cm (increasing the add, if necessary, to +4.00 D); in this case care must be taken not to allow the patient’s head to occlude the light source illuminating the chart.
4. **Instructions to the patient:** Ask the patient to read the letters from left to right across each line of the chart. If the patient responds with a letter other than **C, D, H, K, N, O, R, S, V**, or **Z**, or with a numeral, do *not* score the response as incorrect. Instead, inform the patient of the restricted letter set, and ask for another response. This is in order to support the assumption that the probability of a guess is 1/10. ***Encourage the patient to guess even when they report that the letters appear too faint***.
5. **Recording responses and scoring:** On the score sheet, mark in the grid corresponding to the chart form used, an **X** for each letter incorrectly identified. Terminate testing only when the patient makes two consecutive errors or reaches the end of the chart. Do not terminate the test because the patient has given up and has stopped responding. If this happens, encourage the patient to guess, and score the guesses as ordinary responses. This will help to ensure that the score is based on what the patient *can* see and not on what the patient *believes* he or she can see.
6. Transfer all missed values to the charts on the REDCap forms “Photopic Mars” or “Mesopic Mars” as appropriate. **Enter** the CS value at the final correct letter and the number of errors prior to the final correct letter into REDCap. **REDCap will auto-calculate the log CS score** using the following formula:

The log contrast sensitivity (**log CS**) score is given by the log contrast sensitivity value **at** the lowest contrast letter just prior to two incorrectly identified letters, minus a scoring correction. The letter just prior to the two consecutive misses is called the **final correct letter**. If the patient reaches the end of the chart without making two consecutive errors, then the final correct letter is simply the final letter correctly identified.

##### Example scoring (auto-calculated in REDCap)

In the example below, the test terminates after the patient has read the first letter on the seventh row, because the consecutive letters **O** and **H** were missed. The log CS value at the final correct letter (**H**) is 1.40 and the number of misses prior to stopping is 1 (erred on the K). REDCap autocalculates the Log Contrast Sensitivity from these inputs.

**FORM 1**
| Row | Right eye |
| --- | --- |
| 1 | ☐ C (0.04) ☐ H (0.08) ☐ V (0.12) ☐ O (0.16) ☐ S (0.20) ☐ N (0.24) |
| 2 | ☐ D (0.28) ☐ S (0.32) ☐ Z (0.36) ☐ N (0.40) ☐ R (0.44) ☐ K (0.48) |
| 3 | ☐ N (0.52) ☐ D (0.56) ☐ R (0.60) ☐ H (0.64) ☐ V (0.68) ☐ Z (0.72) |
| 4 | ☐ C (0.76) ☐ S (0.80) ☐ O (0.84) ☐ N (0.88) ☐ K (0.92) ☐ H (0.96) |
| 5 | ☐ K (1.00) ☐ N (1.04) ☐ V (1.08) ☐ D (1.12) ☐ S (1.16) ☐ R (1.20) |
| 6 | ☐ Z (1.24) ☐ R (1.28) ☐ D (1.32) ☒ K (1.36) ☐ H (1.40) ☒ O (1.44) |
| 7 | ☒ H (1.48) ☐ Z (1.52) ☐ C (1.56) ☐ V (1.60) ☐ R (1.64) ☐ K (1.68) |
| 8 | ☐ S (1.72) ☐ C (1.76) ☐ Z (1.80) ☐ D (1.84) ☐ V (1.88) ☐ O (1.92) |

OD: Value of final correct letter
OD: Number of misses prior to stopping
OD: Log Contrast Sensitivity
1.3599999999999999 View equation

7. **Maintenance:** Charts should be stored in their portfolio case, to protect them from light, dust, and physical damage. Do not place other objects on the chart surface that can scratch or dent the charts, and try to avoid touching their front surface, especially in the area where the letters are printed.

#### Summary Procedure

##### Photopic MARS Testing (using general occluder)

###### Patient Set-up

Place MARS chart on bookstand and on top of wooden block on the table. Measure a viewing distance of 20 inches from viewing board to eye. Make sure they don’t lean forward.

**Form 1**= Right Eye **Form 2** = Left Eye

###### Glasses

- **Patient Wears <u>no</u> Glasses:** Use trial Frame and insert a +2.00 sphere for testing or provide +2.00 reading glasses. Patient may refuse to wear the +2.00 glasses if they indicate that they see better without them.
- **Patient wears Distance Glasses with Bifocal:** use current glasses and instruct patient to look through bifocal for testing
- **Patient has their Reading Glasses:** use glasses for testing
- If patient wears contact lenses: instruct them to leave them in for testing

###### Instructions for Patient

“Going from left to right I am going to ask that you tell me the letters that you can see on each row. Towards the end of the chart, you will notice that the letters start to fade away. Continue to stare at the faded letters for about 10 to 15 seconds to see if any of them will appear, if they don’t appear then move to the next row.”

##### Mesopic MARS Testing (using ND-occluder with low luminance filter lens)

- Same testing procedures but with a Filter Lens (will use an actual lens that has the filter attached if using the trial frame for testing; will use the occluder that has the filter if testing with patient’s current glasses)
  - Give the patient a few minutes before starting the Mesopic test.
  - **Form 3** = Right Eye **Form 1** = Left Eye
  - Use the Mars Mesopic data collection sheet; instructions for patient are the same
  - Enter measurement results into REDCap

### 5.11 Monitors and Sensors

Participants will be sent home with three monitoring devices (continuous glucose monitor, physical activity monitor, and an environmental sensor), which they will use for 10 continuous days before returning the devices to the CRC for data download. Each of these monitoring devices are described in the sections that follow. Table 4 is a summary of home monitoring devices used in AI-READI.

**Table 4.** Monitoring devices used in AI-READI, including the variables collected and exported data format.

| **Monitoring Device** | **Manufacturer** | **Data Variables Collected** | **Data Format** |
| --- | --- | --- | --- |
| Dexcom G6 Continuous Glucose Monitor (CGM) | Dexcom, Inc. (San Diego, CA) | Blood glucose measurements (mg/dL) every 5 minutes | .csv |
| Garmin VivoSmart 5 Physical Activity Monitor | Garmin Ltd. (Schaffhausen, Switzerland) | Number of steps<br>Heart rate<br>Sleep duration (circadian and diurnal rhythm)<br>Oxygen saturation | Exported in .FIT file format; converted to mHealth standard for the data set |
| Environmental Sensor | Custom designed, Karalis Johnson Retina Center, University of Washington (Seattle, WA) | Ambient temperature<br>Relative humidity<br>Nitrogen Oxides (NO and NO<sub>2</sub>)<br>Volatile Organic Compounds (VOCs)<br>Particulate Matter (PM1.0, PM2.5, PM4, PM10)<br>Multi-spectral light intensity measurements (11) | .csv |

#### 5.11.1 Continuous Glucose Monitoring - Dexcom G6

Continuous blood glucose monitoring of participants will be performed using a Dexcom G6 continuous glucose monitor (CGM; Dexcom, Inc., San Diego, CA). Participants will be given a general overview of the device and the sensor can be inserted at any time during the in-person visit. Participants will wear the Dexcom G6 continuously for 10 days.

##### Overview

The Dexcom G6 is a real-time, integrated continuous glucose monitoring system (iCGM) that directly monitors blood glucose levels without requiring finger sticks. It must be worn continuously in order to collect data. **Important:** the G6 transmitter will only save data for 30 days, so we must receive the transmitter back from participants and download the data within 30 days from activation or all data will be lost.

##### Dexcom G6 Training Tutorial

Each CRC should familiarize themselves with the Dexcom G6 system, including set-up, by watching the “Apply sensor and check status” video at this site: https://provider.dexcom.com/products/dexcom-g6-pro/training-resources#training-videos

Training Checklist:

– Sensor placement
– Removal
– Skin irritation/bleeding
– Steps for returning to study site
– Uploading
– Saving data

Dexcom Inc. instructions for use of the G6 CGM may be found here: https://provider.dexcom.com/education-research/cgm-education-use/product-information/using-your-dexcom-g6-guide

##### The Data Manager at each site must register for a Clarity Professional account

Go to https://clarity.dexcom.com/professional/registration/. Fill out all the required information in the registration form and click Submit. You will then be asked to create an account for yourself.

If you want to add another person for your site, go to the Staff tab once you log in to Clarity. You can assign them as a standard user or as an administrator. An automated email will be sent to them with an activation link.

To prepare for uploading data, follow the website instructions and install the Dexcom Uploader. You may need to restart your browser to finish the installation. Now you are all set to use Clarity.

##### Dexcom Supplies Waste

###### Dispose of Used Dexcom applicator

1. Sensor needle appropriately retracted in applicator
  a. Place Dexcom applicator in garbage bin
2. Sensor needle dislodged and sticking out of applicator
  a. Place Dexcom applicator in a large sharps container
  b. CAUTION: injury can occur if the needle is not retracted appropriately. Needle has been in contact with bodily fluid. Inform the research manager if you have been injured with a sharp that has been in contact with bodily fluid.

###### Dispose of Used Dexcom Sensor

1. Peel off sensor like a bandaid
2. Place in trash

###### Dispose of Used Dexcom transmitter (contains lithium battery - see local guidelines, UW below)

1. Wash hands, wear gloves before touching used, removed-from-body transmitter
2. Use sanitizing wipe to wipe off any bodily fluids from transmitter
3. Place in a designated small container labeled “Batteries”
4. Each site will dispose appropriately by following institutional policy

##### Study Visit Dexcom Script

Instructions for participants: “This is a Dexcom continuous glucose monitor (CGM). Continuous glucose monitoring is a way to measure your blood glucose in real-time throughout the day and night. A tiny filament called a glucose sensor is inserted under the skin by a skin prick to measure glucose levels in tissue fluid. This filament remains under the skin while you are wearing it. The internal sensor is connected to the transmitter that sits on top of the skin. It is about the size of a quarter and sticks to your skin with medical tape. The CGM records your glucose level every five minutes.

This is the applicator for the sensor. The sensor is what gets glucose information from your body. The sensor applicator inserts the sensor under your skin. You will feel a pinching sensation when the sensor is inserted and may feel discomfort from wearing the sensor. This is the transmitter. This device clips into the sensor and is where the glucose data is located. This is the part that you will mail back after the 10 days are up. Wearing the CGM is different and you may feel discomfort. Hopefully this will reduce as your body gets used to it. [**Note for CRCs**: when picking a sensor site, be mindful of where a participant’s pants waistline and belt falls – it is more comfortable to have the CGM in a spot where their pants waistband does not touch]. It is waterproof for showering, but long episodes of water exposure (swimming) may cause it to fall off. Do you have any questions?”

“At the end of the 10 days, it is time to remove your CGM. Simply peel off the adhesive like a band aid. After the adhesive, sensor and transmitter are removed from your body, disconnect the transmitter from the slot as shown in the instructions. Dispose the adhesive in the trash. Place the transmitter into the provided FedEx box along with the Garmin watch and environmental sensor. Drop the box off in a FedEx dropbox at your earliest convenience or you may bring them back to us in person.”

##### Notes for CRCs

Bleeding at CGM insertion site: Bleeding at the site will be rare, but the sensor should be replaced if it happens while the subject is still on site. Bleeding can often be prevented by not pushing too firmly on the applicator while applying the sensor. Applying too much pressure can cause the sensor to insert too deeply in the skin.

Go over the “Dexcom G6 CGM” instructions that are sent home with the participant, including contact information if they have questions or problems with the CGM and removal of the sensor/transmitter.

##### CGM Guidance

###### Participant comes in without a CGM on

- Explain and administer CGM per normal protocol

###### Participant comes in with a CGM that is not a Dexcom G6

- Explain and administer CGM per normal protocol
- Ensure that the new Dexcom G6 sensor is placed at least 3 inches away from the current CGM and injection site

###### Participant comes in already wearing a Dexcom G6 CGM

- Does the participant have the active Clarity app on their phone?
  - Yes
    § Participant’s device: Go to profile in the bottom right corner, click on “authorize sharing” and then “generate code” for 3 months.
    § Coordinator device: Go to https://clarity.dexcom.com/professional/ website and in the bottom right corner will be a section “View data shared from a smart device”. Enter the code from participant’s clarity app and and press “view reports”
    § This method can only grab previous data. If we want to get 10 days after the study visit, a coordinator could complete this over the phone 10 days after visit.
    § If the participant does not wish to share prior data with the project, proceed normally to attach a second CGM.
  - No
    § Proceed with other options below

##### Placing Dexcom G6 CGM Instructions

Follow the instructions below to set up and place a Dexcom G6 on a participant. If the participant is already wearing a CGM or pump device, ensure the sensor placement is at least 3 inches away from the CGM and/or pump and follow the instructions below.

Step 1: Set up supplies

– Get site reader and turn on by pressing and holding power button for 2-3 seconds and set aside
– Open the box and take out the applicator and transmitter. Set the transmitter aside.
– **Peel off the serial number sticker from the sensor/transmitter box and place it on the instructions sheet for “blinded” participation. Fill in all information on the instructions sheet as you go. You will also use this serial number to enter in the reader to check the connectivity status**.

Step 2: Use applicator to insert sensor

– Choose sensor site on the participant’s abdomen. Consult the Dexcom G6 product manual for an explanation of preferred sites.
  - inches away from injection/pump site
  - Avoid scarring, tattoos, hair, irritation
– Wash and dry hands, clean sensor site with alcohol wipe and let dry
– Remove both labels from the applicator, do not touch the adhesive
– Place applicator on sensor site, hold and break off safety guard
– Press button to insert. After inserting, the CRC should hold in place while counting to 10. **Note:** avoid pushing applicator into skin in order to prevent bleeding
– Discard applicator

Step 3: Attach transmitter

– Clean transmitter with alcohol and let dry
– Insert transmitter tab into slot
– Snap transmitter firmly into place
– Rub around patch 3 times with finger to secure
– Optional: a Dexcom overpatch may be used

Step 4: Check sensor status on receiver

– After the transmitter has been attached at least 90 seconds, select “Check patient status” on the reader
– Follow onscreen instructions and enter the serial number of the sensor inserted
– Wait for the reader to show that the CGM is paired. Once complete, it will show on screen and give you the date the sensor expires - share this with the participant and write it on the instructions handout sheet.
– **Log the serial number into REDCap using the “Device Distribution” form**
– Show the participant the “Device Information” handout that they will return with their home monitoring devices (Appendix C). Show them where you have provided the date that they may remove and return the CGM. Remind them that they should not remove the device prior to that date.
– Make sure the return date is clearly indicated on the “Device Information” sheet. Instruct them that they are to return the sheet with their devices after 10 days of monitoring.
– Double check that the serial number entered into REDCap is correct

- Remind the participant that the battery has a finite lifespan and when the battery dies, the data is lost. We need for them to return the transmitter back to the CRU quickly after the 10 days so that the data is not lost.

Removing the Dexcom G6 Device: see “Removing Dexcom CGM” instructions that are sent home with the participant. Show the participant where this information may be found in the items they are given at their visit.

#### 5.11.2 Physical Activity Monitoring - Garmin Watch

All participants will be issued a Garmin Vivosmart5 watch (Garmin Ltd., Olathe, KS) for monitoring their physical activity for the 10-day monitoring period. The Garmin device will monitor the number of steps taken, heartrate, sleep duration, and oxygen saturation (SpO2 or pulseOx). There is no built in GPS and connected GPS is disabled. The Garmin watch will be placed on the participant during the in-person visit and worn continuously for 10 days, except for periods of charging. Below are the procedures that must be followed when distributing the Garmin Vivosmart 5.

##### 1) Record the activity monitor information in REDCap

This would ideally be done before the Participant leaves the CRU, but could be recorded after the participant leaves. **Record the distribution date and serial number from the back of the watch in REDCap using the “Device Distribution” form**

##### 2) Check the date and time on the watch and erase the .FIT files from the device

Before handing out the watch to a participant, the following must be done: (Important: If this watch was previously worn by a participant, you must do these steps after exporting the data for local storage and uploading to Azure Storage Explorer as described in section 5.11.5)

- **Check that the date and time are correct on the watch for your time zone**. If they are not correct, the watch needs to be reset (as described in 5.11.5) and you should select a different watch and continue the following steps.
- Sanitize the watch.
- Charge the watch to 100%
- Remove *.FIT files from the device. **This should be done the day of the participant visit, as close as possible to the time of placing it on the participant because the watch generates these files everyday even if not worn**.
- Use the charging cable to connect the watch to a computer.
- Open the mounted Garmin drive

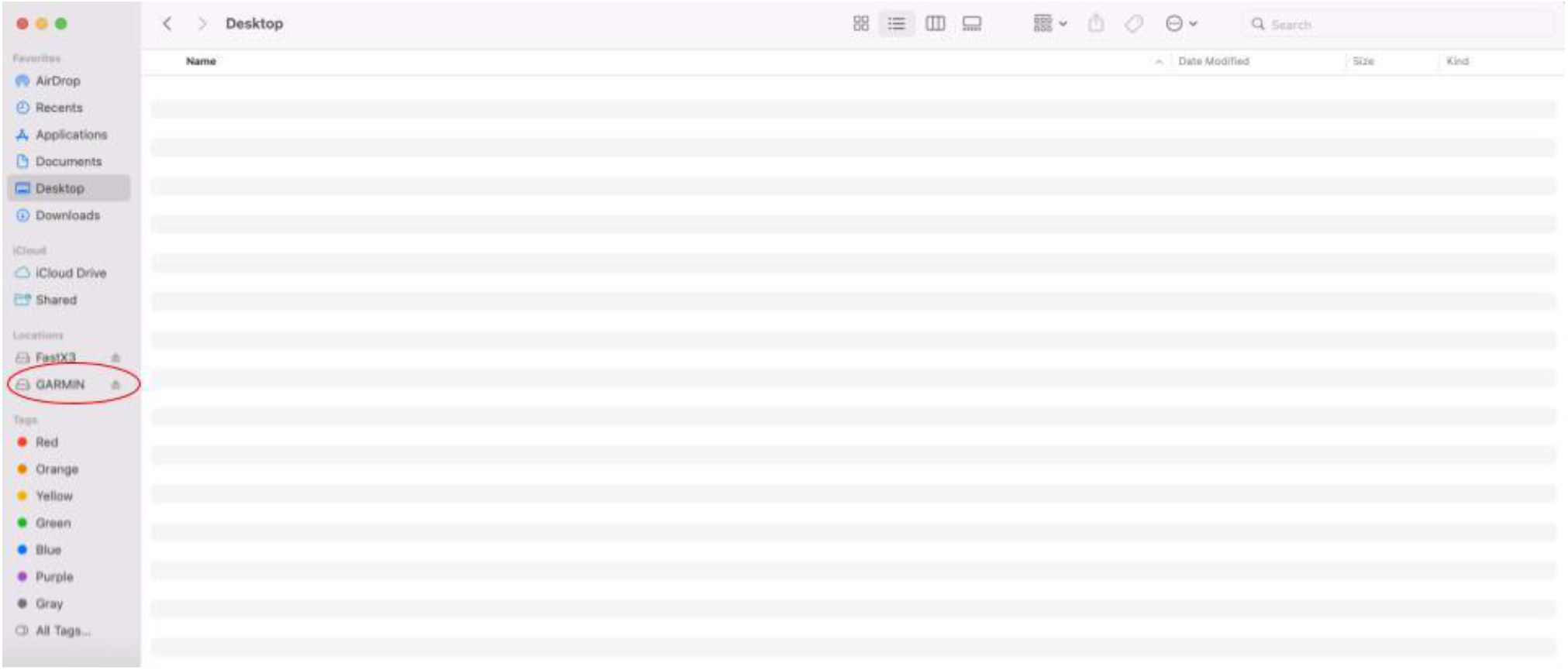
- Open the “GARMIN” folder

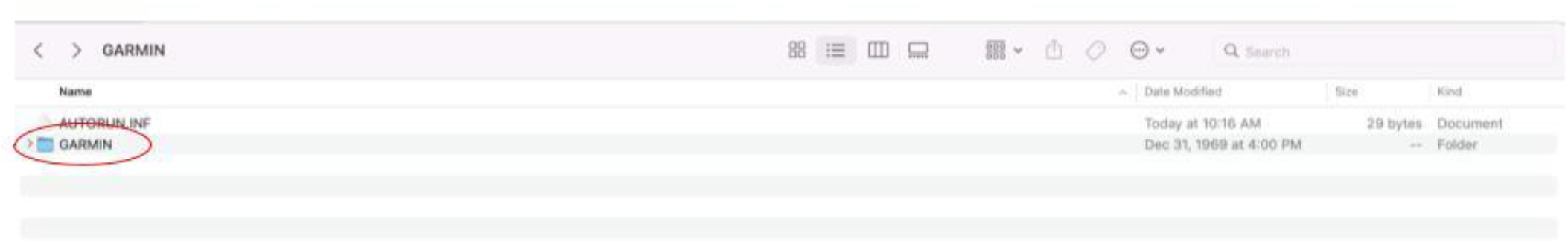
- Open the “Monitor” and “Sleep” folders and erase any *.FIT files from the “Monitor” and “Sleep” folders

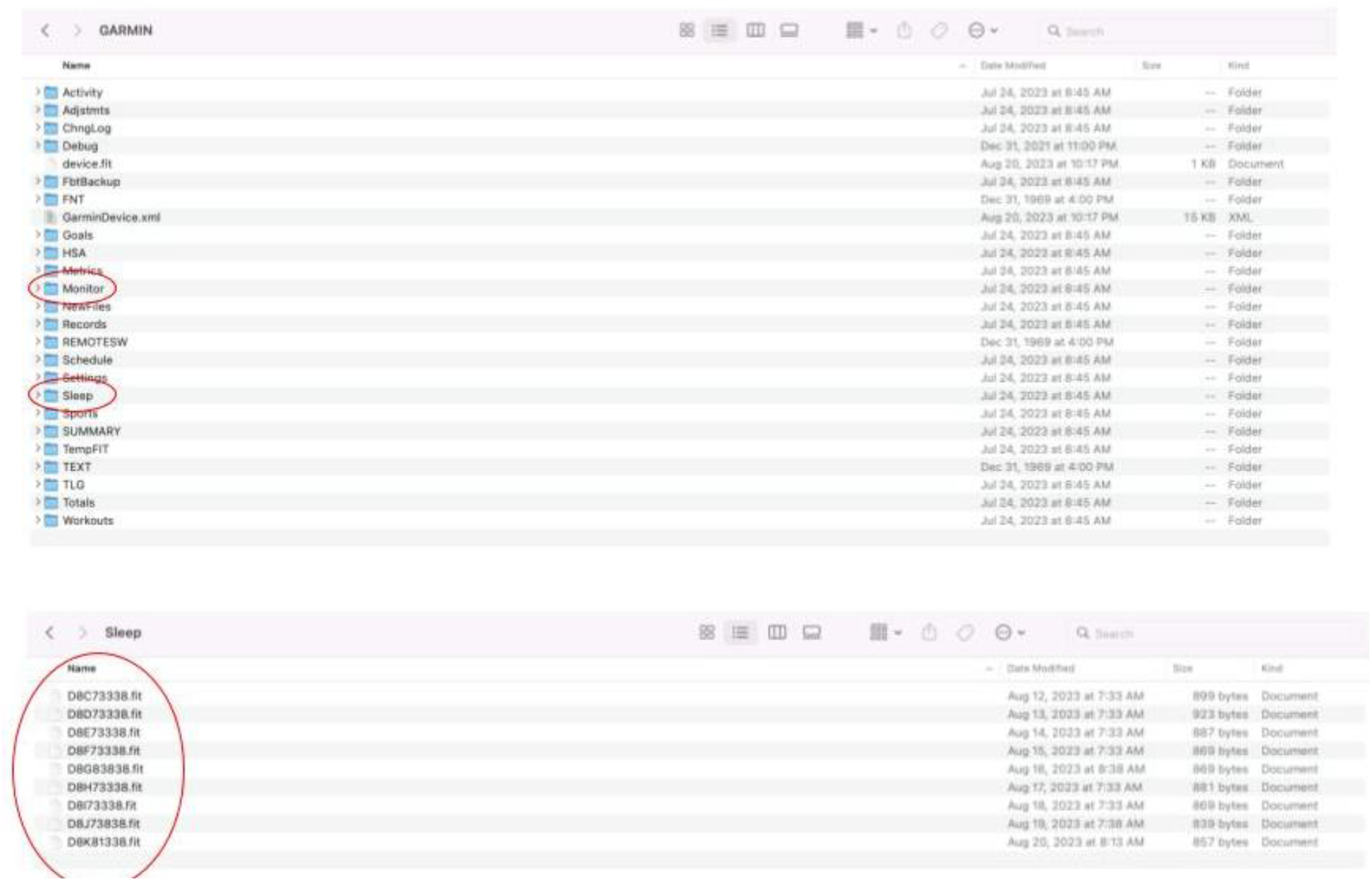
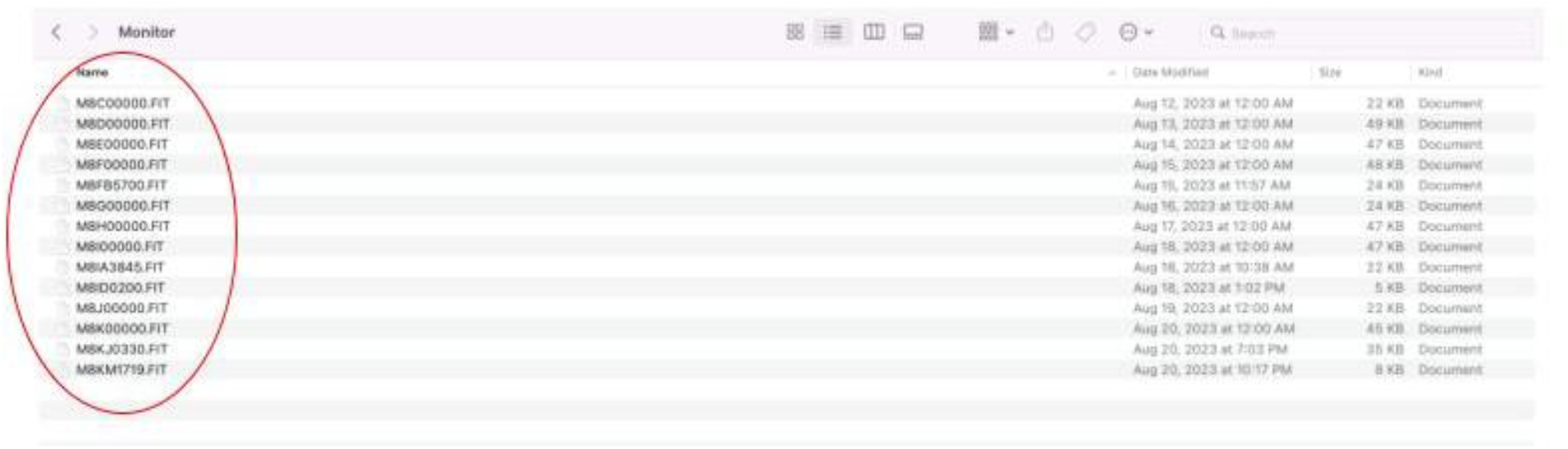
- **Log the serial number on the back of the watch in REDCap**.

##### 3) Demo the watch for participants

###### Script for use with participants

“We are asking you to wear this wrist watch on your left wrist for 10 days (__/__/__ - __/__/__). You may adjust the watch band for comfort, so that it fits snugly on your wrist with just enough room for your right index finger. It should not be so tight that it leaves a print on your skin, or that your movements feel restricted. It will continuously record data pertaining to your physical activities, such as your heart rate and step counts.

The watch takes about 1-2 hours to charge, and we ask you to do this when the watch battery is at around 20% (after 3-4 days of wearing it). Be sure to put the watch back on when it is charged to 100%. Simply plug in the charger, and place the connector to the back of the watch.

At the end of the 10-day period, please check the % battery charge on the watch. If it is less than 50% charged, please charge the battery one last time before returning it to us. Wrap the watch in the bubble wrap/place the watch in the box we provided, and send it back together with the CGM transmitter and the environmental sensor with the prepaid FedEX label. [Show the participant the “Device Return” handout; Appendix C]. You will need to return this handout in the box with your home monitoring devices. Please write in which wrist you wore the fitness tracker on, as well as your dominant hand.” [Show the participant where to place this information].

“If you have any technical issues with the watch please contact the study staff. Information is provided to you in the instructions I will give you to take home.”

#### 5.11.3 Environmental Sensor

The environmental sensor used in AI-READI was custom developed by the Lee Lab (Karalis Johnson Retina Center, University of Washington, Seattle, WA). The environmental sensor monitors various aspects of air quality and does not have audio or video capability (see Table 4, Section 5.11 for a list of variables detected by the sensor). During the in-person visit, provide the participant with the take home instructions for the environmental sensor and review the instructions together. As you read through the instructions, you must demonstrate the device using the steps outlined below.

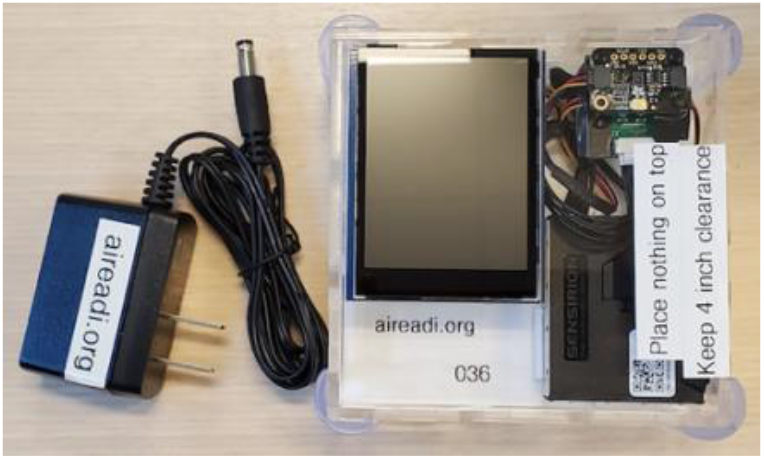

**Demo step 1**: Set up supplies

– Environmental sensor
– Power adapter (be sure you have a wall outlet or a power strip nearby)
– Take Home instructions and “Device Information” sheet

**Demo step 2:** Prepare for power on

– Read the instructions about power up
– Point out that there will be diagnostic messages to watch for
– Point out that the screen will then show current data
– Point out that the screen will go dark on its own after 30 seconds

**Demo step 3**: Power on

– Plug the adapter into the environmental sensor
– Plug the adapter into the wall or power power strip
– Confirm that the sensor powers on, shows diagnostic messages, shows data, and then goes dark
– Demonstrate that the participant can opt to gently touch the screen to see the current data

**Demo step 4**: Review placement and care

– Review the rest of the Take Home sheet regarding placement and care
– Point out the fan on the right side and emphasize the need to leave a 4” space around the device for ventilation (4” is about the width of the sensor)
– Point out that the light sensor is also on top, so nothing should be placed on top
– Point out the openings in the case to explain that it is not watertight and should not be cleaned, sprayed, or placed in an area prone to spills or very high humidity
– Point out that the case should not be placed in direct sunlight or near a heat source

**Demo step 5**: Power down

– Unplug the device to power down
– Point out that the device can be powered down if there is an error message or if the initial location is no longer a good option. The device can be moved (if needed) and plugged in; it will restart data collection automatically.
– **Record the 3 digit unit number for the sensor (e.g. “041”) in the “serial number” field in the REDCap “Device Distribution” form**.

If any of the following issues are observed during the demonstration, please select a different environmental sensor to give to this participant and report the problem to the Data Manager at UW.

- An SD card error that is not resolved by removing and reinserting the SD card
- A current date that is clearly incorrect, e.g. 2000-01-01, or a date that is a week in the future
- Any other error displayed on the screen
- A screen that will not go dark after 30 seconds
- Any flashing or intermittent lights

##### Demo step 6

Show the “Device Information” sheet (Appendix C). Remind them again that this sheet needs to be returned in the box along with their 3 home monitoring devices. Show the participant where to write in the location where they chose to keep the sensor during the 10-day monitoring period.

#### 5.11.4 Return of Monitoring Devices - Instructions for Participants

During their in-person visit, participants will be given instructions on how to use all three monitoring devices. At the conclusion of their visit, they will be given a box, handouts of instructions on the use and return of devices, and a prepaid FEDEX shipping label for returning the devices to the CRU. **Important: Record the FedEx tracking number in REDCap using the “Device Distribution” form**. Be sure to include the “Device Information” handout (Appendix C) along with the instruction sheets. On the “Device Information” handout, write in the date that the participant should remove the CGM and Garmin watch and ship all 3 monitoring devices back. The date provided by CRCs should be 11 days from the day of the in-person visit. Remind the participant to use that date for return of devices.

CRCs may phone the participant with a reminder to return devices and verbally walk them through the process. CRCs and Data Managers at each site should use available reports in REDCap to monitor device returns. FEDEx tracking numbers entered into REDCap can be monitored by CRCs to determine when shipments are initiated. Participants may also return the devices in person. The participant will receive their honorarium only when the devices are returned and received by the CRU and their receipt is logged in REDCap. **Use the REDCap form titled “Device Return” to log the return of all home monitoring devices. If a device was not returned or is returned without data, indicate this on the device return form and provide an explanation in a comment linked to the Device Return form**.

##### Script for instructing participants

At the end of your visit, “we have provided you with a FedEX Express box (FedEx Box size: 8-3/4” x 2- 5/8” x 11-1/4”, FedEX Express box S2, part number 167027), a prepaid FedEX shipping label, and 3 labeled bubble wrap sleeves, one for each of the items below

- The environmental sensor and power cord
- Dexcom CGM (Continuous Glucose Monitoring) transmitter
- Garmin watch / physical activity monitor
- Instructions for returning the devices to us on a sheet titled “Device Information”

Please place the environmental sensor, Dexcom CGM transmitter, and the Garmin watch fitness tracker in the box provided and mail back to the study center on or slightly after the date indicated on the “Device Information” sheet. Be sure to include the chargers for the Garmin watch and environmental sensor in the box. You have been provided with a labeled bubble wrap sleeve for each of these devices to protect them during shipping. Please be sure you have written in which wrist you wore the fitness tracker on, what your dominant hand is, and where you placed the environmental sensor in your home on the “Device Information” sheet and include it in the box as well.”

#### 5.11.5 Receiving Returned Monitoring Devices and Extracting Data - Instructions for CRCs

There are procedures that must be followed when monitoring devices are returned from participants and received in the CRU, to ensure that the devices are properly readied for the next user.

##### REDCap

Any CRC may receive and log returned devices. When a CRC receives devices from a participant, they should immediately unpack the box and log the date of return in the REDCap form titled “Device Return” for that participant. The location of the environmental sensor and wrist information for the Garmin watch should also be notated in this REDCap form. Data extraction from returned monitoring devices must be performed immediately after arrival, because there is the potential for data loss from both the Dexcom and Garmin devices if the battery is drained. **Note**: Data Managers should monitor the device returns using available reports on REDCap. Coordinators should contact participants who have not returned devices >3 weeks after their in-person visit date. Devices not returned or lost in transport should be documented in the “Device Return” form using the questions regarding data export from monitoring devices.

##### Dexcom G6 CGM

**Step 1:** Log the date of transmitter receipt in the Dexcom G6 section of the “Device Return” REDCap form.

**Step 2: Immediately** download data from transmitter to reader. Store data files on a local hard drive or server for later upload to Azure Storage Explorer. To export data collected by the Dexcom G6 PRO sensor, you will need the reader and the transmitter.

- Each transmitter has its own serial number; you can find this number on a sticker in the box, or on the back of the transmitter itself. This is the same number you had used during the sensor insertion.
- Perform the following to download data from the transmitter to the reader. The transmitter and the reader should be placed close to each other for this step.
- Push the oval button on the reader. Press “OK” on the touchscreen when it says it is ready for the next patient
- Press “Download Data”
- Find the six-digit serial number for the returned transmitter. Press “Next” and then enter the serial number using the touchscreen. Verify that the serial number is correct and press “yes”. The reader will pair with the transmitter and the data will be downloaded.
- Connect the Reader to your computer with a micro-USB cable. Visit https://clarity.dexcom.com/professional/patients/ and find the Add New Patient button on the upper right corner.
- When adding a new patient, enter the following information:
  – **First Name**: enter Participant ID#
  – **Last Name**: enter AIREADI
  – **DOB**: enter 01/01/actual birth year
  – **Patient ID**: enter AIREADI-Participant ID#
- Follow the simple instructions on the website and upload the data.
- Once the data is uploaded, export the participant data as a .csv file and save on a local computer and/or server. **Do not rename the** .**csv file. We will use the name that exports with the data from Clarity**. All CGM data files should be saved to local storage for later uploaded to Azure Storage Explorer (described in Section 8.4.7).

**Step 3**: Download the pdf reports of results. To do so, select “Save or Print Reports”, make sure that all reports are highlighted, then select “Save as PDF” and store locally. These are the reports that will be provided to participants.

**Step 4**: Send Dexcom pdf reports to participants along with the “Interpreting your CGM Report” guidance handout, which explains how to interpret those results (see Appendix D).

**PDF reports should be sent securely to participants** to help prevent the possibility of re-identification in the event of an email hack, since the reports reference the AI-READI Participant ID#. Sites may choose to send through an encrypted email server or PHI-compliant Sharefile server (or equivalent) for protection. We recommend that each site enlist the help of their IT department for determining the safest way of distributing participant Dexcom files. A paper form of the report (with subject ID redacted) may also be provided in person or via mail.

**Step 5:** Discard transmitter only after you have verified that data is successfully downloaded from Clarity and saved to local storage.

##### Environmental Sensor

When the environmental sensor is received back from the participant, **log the date of receipt and the location of the sensor in the home on the REDCap form titled “Device Return”**. Next, follow the instructions below for exporting the data and preparing the sensor for the next participant.

After the environmental sensor is returned, a CRC will need to retrieve data from the environmental sensor using the following steps:

**Data retrieval step 1**: Set up supplies

- Environmental sensor
- Blunt nosed tweezers (supplied by UW)
- Mini SD card reader
- Computer with upload capability (on a network)
- Hands should be clean
- Reduce the likelihood of electric discharge by using a clean solid work surface, avoid touching fabrics such as sweaters or blankets

**Data retrieval step 2**: Remove the mini SD card

- Hold the environmental sensor upside down with one hand
- Using the back end of the blunt nosed tweezers, gently press on the SD card and release. The SD card slot will make a small click and the SD card will extend out a few mm from the slot.
- Using the blunt nosed tweezers, gently grasp the end of the SD card and remove it from the environmental sensor. Avoid touching the small metal contacts on the SD card.
- Place the mini SD card into the card reader; connect the card reader to the computer.

**Data retrieval step 3**: Creating a storage folder and moving environmental sensor files

- Participant data will be stored in a series of files that are named by date and time. Most of the files will be the same size (about 145 KB) and the last file is likely a smaller size. It is not a problem if files are of various sizes.
- **Create an outer folder to hold the data files on your local computer** Name the folder using the participant ID and the environmental sensor ID in this format: **ENV-pppp-nnn** where pppp is the 4 digit participant ID (1001 to 9999) and nnn is the 3-digit environmental sensor assembly ID. Important checklist
  - Keep ENV as the first 3 letters
  - Use only the dash symbol - when separating parts of the folder name
  - Keep all 3 digits, e.g. 042, for the environmental sensor. Do not shorten.
  - Keep all 4 digits of the patient ID. Do not add or remove any characters.
  - Make sure that the folder does not contain any subfolders. Only files ending in csv should be in this folder.
  - **Example for patient 1234 and environmental sensor 042: ENV-1234-042**
- Open the folder in the SD card containing the environmental sensor files
- Copy or move all of the files into your newly created outer folder (discussed above).
- Store the data folder on your local computer or server for later upload to Azure Storage Explorer (described in Section 8.4.7).

**Data retrieval step 4:** Prepare the card for the next participant

- Double check that all files were successfully copied from the SD card to your new folder (from Step 3).
- Delete all files on the **SD card**. Do not reformat the card. Be sure to empty the trash/recycling bin on the computer before ejecting the SD card.
- Holding the environmental sensor upside down, gently place the mini SD card into the slot with the text facing up.
- Using the back end of the blunt nosed tweezers, gently press the SD card into the slot until you hear a “click” or feel the card snap into place.
- Optional: For the first few sensors, you may want to confirm that the SD card is properly seated. Plug in the device, watch the initial diagnostic messages, and if it reaches “All finished” then you know that the SD card is properly positioned. If instead you get a red error message regarding the SD card, unplug the sensor, remove the card, and re-insert.

##### Physical Activity Monitor (Garmin Vivosmart5)

After receiving the returned device, **log the date of receipt, the wrist used for data collection, and the participant’s dominant hand in the REDCap form “Device Return”**.

Next, perform the following to ensure that the data is extracted from the watch and the watch is prepared for the next participant:

**Step 1:** Use the charging cable to connect the watch to a computer.

- The watch should appear as a mounted drive.

**Step 2:** Make a folder called **“FIT-ParticipantID”** on your Desktop or harddrive (where ParticipantID is the 4 digit study ID used for that participant)

**Step 3:** Open the mounted Garmin drive.

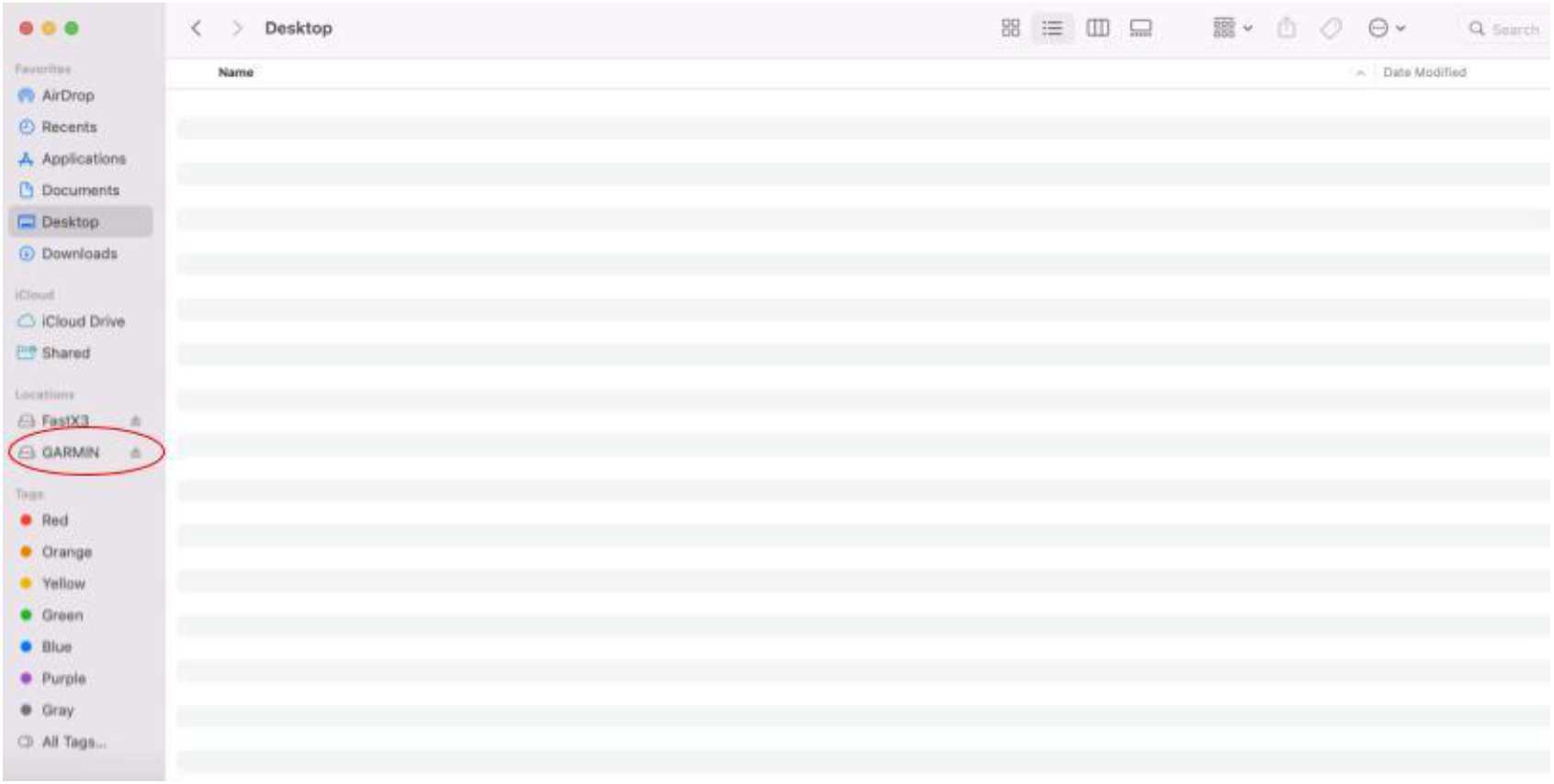

**Step 4:** Copy the entire “GARMIN” folder to the new FIT-ParticipantID folder you created on your Desktop or harddrive

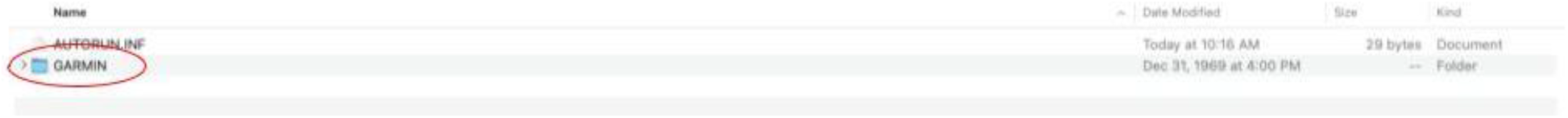
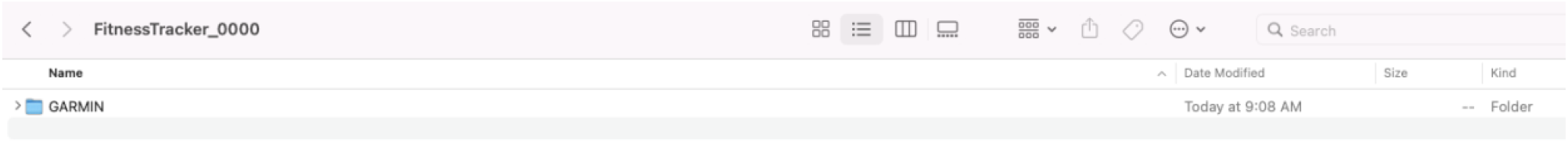

**Step 5**: Save the FIT-ParticipantID folder on your local computer and/or server for later upload to Azure Storage Explorer (described in Section 8.4.7).

**Step 6:** Prepare the Garmin watch for the next participant

- Verify that the Garmin folder was saved appropriately to your new folder
- Open the Garmin folder (from the the mounted drive representing the watch; NOT the Garmin folder in your newly created folder!)
- Delete the .FIT files in the “Monitor” and “Sleep” folders before assigning it to a new participant, using the instructions outlined in Section 5.11.2.
- <u>Check the date and time on the watch</u>. If the watch has the incorrect time or date for your time zone, remove it from circulation until you have followed the instructions below to reset the watch.

**\*If a watch comes back with the wrong date or time (or if there is a time change), you will need to factory reset the watch and re-establish proper settings**.

**\*\*Do not perform a factory reset until you are certain that you have saved the data from the participant. First you must copy data from the previous participant using the instructions above.\*\***

##### Factory reset the watch

1. Push oval button on watch (may need to push twice to get to menu)
2. Scroll up to Settings
3. Scroll up to System
4. Scroll up to Reset
5. Choose Delete Data and Reset Settings
6. Click the screen when the warning appears
7. Click the ✓

##### Set the watch up again (Garmin Express)

1. Download Garmin Express for Mac or Windows: https://www.garmin.com/en-US/software/express/windows/
2. Open the Garmin Express app
3. Plug in the watch to a USB drive on your computer using the black cord
4. Click “Add Device” in the Garmin Express app:
5. It will detect the watch you plugged in. (You may receive a message that Garmin Express would like to access a removable device - click OK)
6. Click “Add Device”:
7. Sign into the shared Garmin account using the login information provided to each site.
8. Click Yes that this the primary wearable
9. Click Next on the next two pages asking about settings and completing your profile (do not add any information here; use default settings)
10. Make up any name for the Nickname, you can’t reuse nicknames
11. When the screen shows the message “You’re up to date!”, the watch is set up
12. Eject the watch on your computer and unplug the watch from the USB cord. **Do not push any button until you read the section below**.
13. Click the Home Button in the upper left of Garmin Express app.
14. Right click on the watch icon and “Remove from Express”:

##### Settings on the watch

1. If it offers to update software, decline.
2. Push the oval button the watch, scroll down to Settings and set the following as specified (click ✓ to set):
  a. Wrist Heart Rate -> Pulse Ox -> During Sleep
  b. Phone -> Status Phone: Off
  c. Morning Report -> Status: Off
  d. System ->
    i. Brightness -> 1
    ii. Alert Vibration -> Off
    iii. Gesture -> Mode -> Off
    iv. USB Mode -> Mass Storage

**Note: You cannot change the USB mode if the charging cable is attached to the watch**.

### 5.12 Retinal Imaging

Retinal imaging for each device will be done OD (right eye) and OS (left eye). Fundus photos of each eye will be acquired when undilated using the Optomed Aurora handheld camera. The remaining retinal imaging will be performed on dilated participants using procedures outlined here. **All retinal imaging must be logged in REDCap using the form titled “Retinal Imaging”**. If a scan is unable to be performed, an explanation must be provided in REDCap. A retinal imaging checklist has been created for technicians to use during a participant’s visit (Appendix E). **Note:** if the participant wears contact lenses, they must be removed at this time. The CRU should make available contact lens cases and contact solution for temporary storage.

#### 5.12.1 Undilated Fundus Photography (Optomed Aurora)

Undilated fundus photos will be taken using the Optomed Aurora IQ camera (Optomed, Oulu, Finland). **This imaging is to be done BEFORE the patient is dilated**.

Comprehensive instructions on the use of the Optomed Aurora may be found here: https://www.optomed.com/us/wp-content/uploads/sites/2/2020/12/60000888-R-7.0-Optomed-Aurora-User-Manual.pdf. The users manual provides helpful images showing camera alignment and software functions.

1. Pick up Aurora from dock, if camera does not auto power on, press power button.
2. Using Optoroller joystick to create new patient study or find patient in list.
3. Select “Four image sequence”
4. Then select “New Patient Study”
  a. **For Name**: enter Participant ID#
  b. **For Patient ID**: enter AIREADI-Participant ID#
5. If possible, dim room lights this will help with pupil size, but is not 100% necessary.
6. The photographer and patient should be on the same level, this is ideal to have level alignment: 1) photographer eyes through 2) camera to 3) patient eyes (see user manual for images). Seated is recommended. Facing the patient and seated in proximity by placing one knee between the patient’s knees. Have patient sit forward in chair if needed, or whatever is best for the photographer to be in a relaxed and comfortable position. **Start with the right eye (OD)**.
7. Instructions to patient:
  a. Keep head still, while sitting still in general.
  b. Keep both eyes open while covering fellow eye with palm of hand (having both eyes open assists in keeping test eye open wide).
  c. Focus subject eye on fixation target/red light.
8. The acquisition should automatically start on the Right Eye Disc
9. Hold the Aurora handle with dominant hand, place finger on trigger.
  a. Other hand supports the front of camera and makes any fine tune adjustments. There is a divot for your thumb on the bottom of the camera barrel.
  b. Back hand provides stable base to move camera in/out only. Try to hold back hand steady.
10. Stabilize front of camera by placing fingertips of your front hand on patient’s forehead. **Make micro adjustments with the front hand while keeping the camera perpendicular to the patient’s eye.
11. Keep camera level, perpendicular to patient. Slowly move straight on toward the patient’s eye. Eye cup will press around the examined eye as you approach.
12. Use the screen to guide you to the retina. The retina will appear light gray through the pupil. The closer you get the retina view will open filling the screen. **PRO TIP: Center the gray retina reflection seen through the pupil 12:00 spot as soon as you see it. Keep this centered on the approach, trying to align and capture in one motion. The correct imaging distance from the eye is about 2 cm or ¾ inch. The flexible cup keeps unit from touching patient.
13. Alignment:
  a. Aim help circle (parentheses) on the screen will turn green when the retina is fully in view and fills the screen.
  b. The focusing rectangle will appear when autofocus is engaged and will turn green when focal distance is achieved (you can initiate the autofocus by pressing trigger half-way down). Move in another 1 MM for optimal positioning.
14. Completely press down trigger (hold down momentarily) to capture image.
15. Review the image for complete capture of desired image and pathology. Then push down on the button to:
  a. Toggle to “Continue”, if you’re satisfied with the image. The sequence will move on to the next image type in the four image sequence. The acquisition sequence is right eye disc (target is nasally), left eye disc (target is nasally), right eye macula (target is central) and left eye macula (target is central).
  b. Or toggle to “Retake”, if you’re not satisfied with the image.
  c. Note: if you’re using the “Four image sequence” when you create the subject, you should not have to assign the eye as right or left. When the eye is assigned, the file name will start with ‘OD’ or ‘OS’. If the eye is not assigned, the file name will start with ‘IMG’.
16. At the end of the sequence, you will see a question “Sequence completed. Start a new sequence?” Select “No” to end the session.
17. Double check image quality using the traffic light analyzer. 3 green lights is best. 2 green and 1 yellow is acceptable. Refer to detailed image quality analysis below. Note: If there is reflection in the upper part of the image the imaging distance is too short/close. If there is reflection in the lower part of the image the imaging distance is too long/far away.
18. Repeat steps 5-13 for the left eye (OS).

Patient may see the red target light filling vision. Correct depth position.

If you need additional assistance or troubleshooting: refer to the user manual provided in the Aurora camera case (or link provided above) or quick guide. You may also access training videos and submit requests for help on the Aurora Product Page at https://www.optomed.com/us/products-and-solutions/fundus-cameras.

#### 5.12.2 Pupillary Dilation

Effective pupillary dilation must be achieved to obtain high quality retinal imaging. AI-READI has an extensive retinal imaging protocol and dilation of each eye is essential. Dilation involves the administration of dilation drops by CRCs. To ensure that these protocols are carefully followed, a guideline for pupillary dilation is provided below. Questions regarding this information should be directed to the PI at your site or your site manager.

Many individuals have had dilated eye examinations previously. However, it is important to provide a clear explanation to the participant.

1. **Explanation of dilation procedure**
  a. Before administering the dilation drops to the participant, the study coordinator should sufficiently explain what they are giving the subject as well as why they are giving them the drops. For example, “…I am now going to give you some dilating drops. These drops will dilate your pupils allowing us to gather information about the back part or retinal layers of your eyes. These drops are no different from the dilation drops used in your normal eye exam with your eye care professional.” The study coordinator should explain to the participant that their eyes will remain dilated for approximately 4-6 hours and during that time they will be sensitive to light.
  b. Any allergies or physiological conditions existing within the participant’s medical history that are related to pupillary dilation should be thoroughly discussed with the subject and an authorized medical practitioner prior to administration of the dilation drops to ensure proper adherence to the study protocol.
1.5 **Administering the proparacaine hydrochloride ophthalmic solution drops.** This is a site-specific decision, as not all data collection sites will utilitze these drops
  a. Before administering proparacaine hydrochloride ophthalmic solution, tell the patient to sit firmly in a chair with proper overhead lighting.
  b. Ask the subject if they have had this done before and if they had experienced any problems with the drops. If the participant did have problems from proparacaine hydrochloride ophthalmic solution, discuss with the subject and determine if further inquiry should be followed based upon study protocol. Confer with the PI or data manager for further instructions. If not, continue to c.
  c. Locate the proparacaine hydrochloride ophthalmic solution drops and visually confirm by reading the label that they are in fact the correct drops and are not expired.
  d. Provide a tissue to the subject for blotting excess tears as necessary.
  e. Have the participant lean their head back slightly and have them look back or up as much as possible.
  f. Starting with the right eye (OD), with the index finger of your free hand, gently pull down the lower eyelid. With the hand holding the drops, place 1 drop of proparacaine hydrochloride ophthalmic solution within the lid margin and conjunctiva from approximately 1 to 2 inches above the eye.
  g. Move over to the left eye (OS) and repeat step f.
  h. With a single drop, the onset of the effect begins within 30 seconds and persists for 15 minutes or longer. Start the dilation process after at least 30 seconds from the administration of proparacaine hydrochloride ophthalmic solution drops.
2. **Administering the dilation drops**
  a. When dilating the participant, have them sit firmly in a chair with proper overhead lighting.
  b. Ask the subject if they have had their eyes dilated before and if so, did they have any issues with dilation. If the participant did have problems from dilation, discuss with the subject and determine if further inquiry should be followed based upon study protocol and confer with the PI or data manager for further instructions. If not, continue to c.
  c. Locate the dilation drops and visually confirm by reading the label that they are in fact dilation drops and are not expired.
  d. Provide a tissue to the subject for blotting excess tears as necessary.
  e. Have the participant lean their head back slightly and have them look back or up as much as possible.
  f. Starting with the right eye (OD), with the index finger of your free hand, gently pull down the lower eyelid. With the hand holding the drops, gently rest the lower part of the palm on the subject’s forehead and without touching the eye or surrounding tissue, place 1 drop of Neofrin 2.5% (phenylephrine HCL) within the lid margin and conjunctiva from approximately 1 to 2 inches above the eye.
  g. Move over to the left eye (OS) and repeat step f.
  h. Repeat steps f and g with Mydral 1% (tropicamide) in the OD and OS.
3. **Problems or consequences**
  a. Dilation usually takes up to 15 minutes to fully take effect, but in some instances may take up to 20 minutes.
  b. The subject may experience a stinging sensation in the eye. Should this occur, reassure the subject that this should only last a few seconds and will subside quickly.
  c. The subject may also experience some mild to moderate photophobia or sensitivity to light following dilation. If the subject has no sunglasses of their own, provide them with a pair of disposable sunglasses available within the CRU before they depart the visit.
  d. If any allergic reaction such as redness, inflammation, or visual disturbance occurs, contact the PI or site manager for further instructions.
  e. In the event that the participant experiences dry eye symptoms during retinal imaging procedures, artificial tears may be administered to ameliorate the condition.

#### Note to CRCs

The retinal imaging protocol is not a comprehensive eye examination, and thus participants should be informed that it does not replace a comprehensive eye examination that they can receive from their own ophthalmologist or optometrist. Retinal imaging technicians are not clinicians and cannot comment on retinal images for participants, other than the 3 conditions for which we have IRB approval to discuss with the participant. See “Incidental Findings” in Section 9.2 for more information.

### 5.12.3 Dilated Retinal Imaging

Multiple imaging modalities are used in the AI-READI protocol. Detailed instructions for the use of each device may be obtained from the manufacturer. Table 5 below summarizes the imaging devices used, the types of scans, and the data format provided in the AI-READI dataset. Exact accounting of the scans taken for each participant will be entered into REDCap using the Retinal Imaging Checklist found in Appendix E.

**Table 5.** Retinal Imaging Devices, acquired scan types, and data format.

| Imaging Device | Manufacturer | Scans Collected | Number of images per participant | Data Format |
| --- | --- | --- | --- | --- |
| Aurora IQ | Optomed (Oulu, Finland) | Fundus: macula and disc centered (CFP) color fundus photo (undilated) | 4 | DICOM |
| EIDON widefield truecolor confocal fundus system | iCare USA, Inc. (Raleigh, NC) | Ultra-wide Field Central CFP<br>Ultra-wide Field Nasal CFP<br>Ultra-wide Field Temporal CFP<br>Ultra-wide Field Central IR<br>Ultra-wide Field Central FAF<br>Mosaic of Ultra-wide Field images | 12 | DICOM |
| Spectralis HRA optical coherence tomography (OCT) and optical coherence tomography angiography (OCTA) | Heidelberg Engineering GmbH (Heidelberg, Germany) | Optic Nerve Head-Radial Circle-High Resolution (OCT)<br>Posterior Pole Macula-HR-61 lines (OCT)<br>Macula-20x20-HS-512 lines (OCTA) | 6 | DICOM |
| Maestro2 3D OCT-1 | Topcon Healthcare (Itabashi-ku, Tokyo, Japan) | 3D Wide(H) 12x9-512x128 (OCT)<br>3D Macula 6x6-512x128 (OCT)<br>Macula 6x6-360x360 (OCTA) | 6 | Exported in .fda file format; converted to DICOM |
| Triton DRI OCT | Topcon Healthcare (Itabashi-ku, Tokyo, Japan) | 3D(H)+Rad 12x9-512x256 (OCT)<br>Macula 6x6-320x320 (OCTA)<br>Macula 12x12-512x512 (OCTA) | 6 | Exported in .fda file format; converted to DICOM |
| Cirrus 5000 | Carl Zeiss Meditec AG (Jena, Germany) | Disc Cube, - 200x200 (OCT)<br>Macula Cube, - 512x128 (OCT)<br>Macula, -6x6 (OCTA)<br>Disc, -6x6 (OCTA) | 8 | DICOM |
| Fluorescence Lifetime Imaging Ophthalmoscopy (FLIO) | Heidelberg Engineering GmbH (Heidelberg, Germany) | Macula-centered | 2 | Exported in .sdt file format; converted to DICOM |

### 5.12.4 Quality Recommendations for Structural OCT Images

Please note that the instrument quality scores do not reflect all possible quality issues that may occur

#### Current recommendations for Topcon structural OCT scans

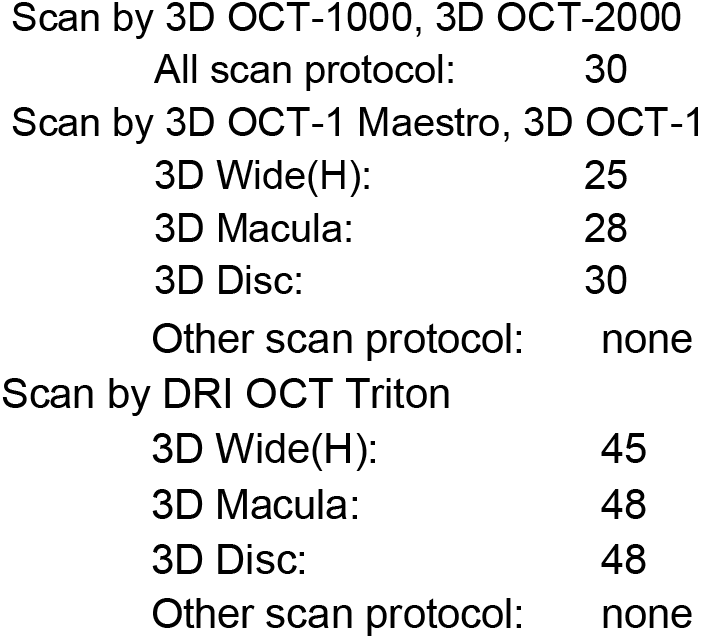

#### Cirrus quality recommendations

**OCT:** Signal strength 6/10 or higher for both macula and ONH scan generally indicates a good quality scan

#### Spectralis quality score recommendations

**<H5>OCT**

Qualtiy Score:

15 or less,generally indicates a poor scan quality.

15 – 25 the scan quality is considered marginal.

25 or above the scan quality is considered good.

## 5.13 Biospecimen Collection

Blood samples will be collected and processed for both clinical lab testing and biorepository storage, using the procedures outlined in this chapter. All three Data sites will use a standardized approach to clinical lab testing: plasma, serum, whole blood and urine samples will be stored frozen at -80°C (locally) and then batch-shipped on dry ice to UW’s Diabetes Institute Clinical Research Unit (CRU). The samples for clinical testing will be stored frozen at -80°C until testing is performed. After receiving samples from each site, the UW CRU staff will store the boxes in a -80C freezer until the UW’s Nutrition Obesity Research Center (NORC) staff is able to transfer them for testing quarterly. UW’s NORC lab will perform all clinical lab testing including urine, except for the CBC test. The complete blood count (CBC) test must be performed at a lab local to each Data site on fresh whole blood. For biorepository samples, each Data site will process the blood according to the instructions described below. CPT Tubes (for Peripheral Blood Mononuclear Cell (PBMC) samples) and PAXgene RNA tubes must be processed and shipped daily at ambient temperature to the UAB Biorepository for next day delivery. All other biorepository samples are to be processed, frozen and stored at -80°C at each site and then batch-shipped on dry ice to the UAB Biorepository. The frequency of batch shipments to UW and UAB will be decided by each Data site, but should at minimum occur quarterly. A summary of biospecimen processing, and purpose is provided below (Table 6).

**Table 6.** Biospecimen collection, processing, and purpose.

| Specimen Collection | Sample Type | Location | Processing Details | Purpose |
| --- | --- | --- | --- | --- |
| EDTA vacutainers |  |  |  |  |
|  | Whole blood | Local clinical lab | None | Complete Blood Count (CBC) analysis |
|  | Whole blood | UW NORC*† | None | Testing for HbA1c |
|  | Plasma | UW NORC*† | processed locally at data site on the day of collection | Testing for NT-proBNP§, troponin-T, C-Peptide, and insulin |
|  | Plasma | Biobanking at UAB CCTS‡ | processed locally at data site on the day of collection | Future proteomics and metabolomics studies |
|  | Buffy coats | Biobanking at UAB CCTS‡ | processed locally at data site on the day of collection | DNA extractions for future genomics |
| Serum Separator (SST) vacutainers |  |  |  |  |
|  | Serum | Biobanking at UAB CCTS‡ | processed locally at data site on the day of collection | Future proteomics and metabolomics studies |
|  | Serum | UW NORC*† | processed locally at data site on the day of collection | Testing for CRP-HS§, glucose, BUN§, creatinine, carbon dioxide, total protein, albumin, globulin, bilirubin, alkaline phosphatase, AST§, ALT§, electrolytes§, and lipids§ |
| Urine collection kit | Urine | UW NORC*† | processed locally at data site on the day of collection | Testing for creatinine and albumin |
| CPT Mononuclear Cell Preparation Tubes | PMBCs¶ | Biobanking at UAB CCTS‡ | UAB – processed locally on day of collection<br>UCSD/UW – tubes sent to UAB for next day processing | Future immunological studies; generation of iPSCs |
| PaxGene RNA vacutainers | Stabilized, un-isolated RNA | Biobanking at UAB CCTS‡ | UAB – processed locally on day of collection<br>UCSD/UW – tubes sent to UAB for next day processing | Future gene expression studies |

\* **UW NORC:** University of Washington Nutrition Obesity Research Center
† Samples analyzed by the UW NORC laboratory (serum, plasma, whole blood, and urine) are first stored locally at UAB and UCSD at -70° C, then batch shipped to UW on dry ice approximately once per quarter.
‡ **UAB CCTS:** University of Alabama at Birmingham Center for Clinical and Translational Science
§ **NT-proBNP:** N-terminal pro-B-type natriuretic peptide; **CRP-HS:** C-Reactive Protein, High Sensitivity; **BUN:** Blood Urea Nitrogen; **AST:** aspartate aminotransferase; **ALT:** alanine aminotransferase; **Electrolytes:** sodium, potassium, chloride, calcium; **Lipid Analysis:** total cholesterol, HDL cholesterol, LDL cholesterol, triglycerides
¶ **PMBCs:** Peripheral Blood Mononuclear Cells

All biorepository samples will be held at UAB’s Biorepository laboratory in the UAB Center for Clinical and Translational Science (CCTS). Limited inventory lists will be provided to fairhub.io with each data release.

### 5.13.1 Preparation

The following vacutainers are to be collected from each participant **in the order of draw recommended** by the manufacturer (https://www.bd.com/a/92195). Blood and blood derivatives will be used for either same day testing (CBC), batch clinical lab testing at the University of Washington NORC, or biobanked at UAB for future research purposes. The numbers in parentheses next to each tube type are Becton, Dickinson and Co. (BD) SKU numbers. The following tubes must be organized and labeled prior to a participant’s visit, using instructions in the sections that follow.

· 2x 5.0ml SST gold top (BD 367986)
· 2x 6.0ml purple (EDTA) top (BD 367863)
· 1x 3.0ml purple (EDTA) top (BD 367856)
· 2x sodium citrate CPT tubes (BD 362761)
· 1x Paxgene RNA (BD 762165) – keep upright after inverting/mixing
· Urine (BD Vacutainer Urine Collection Kit)

Each site may need to complete requisition forms for phlebotomy services and/or local lab testing for the CBC test. These requisitions should be prepared prior to the participant’s visit.

### 5.13.2 Vacutainer Tube Labeling

Vacutainer labels: UAB will provide pre-printed labels for blood collection. These labels will contain the participant ID and site name (UW, UCSD or UAB).

There will be 10 collection devices (8 vacutainers, urine cup, urine vacutainer) per participant that must be labeled.

- Example of one set of labels (1.5” x 0.75” in size) to be provided:

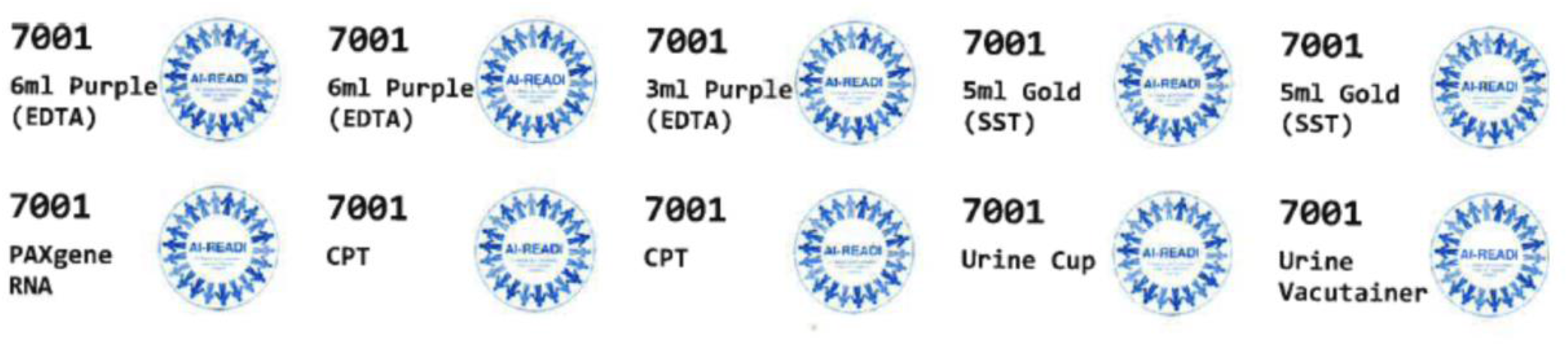

Used to label:

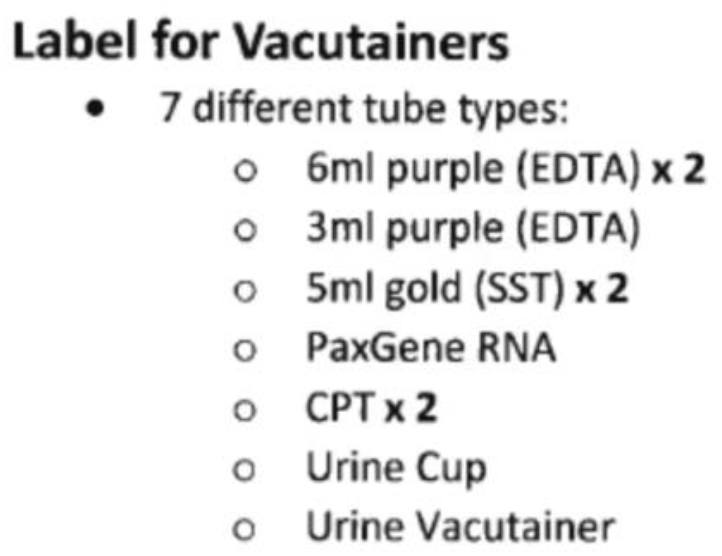

Sufficient labels (2”x1”) for 1400 participants (10 labels per participant) from each site will be provided. Additional labels beyond this number will be provided upon request. Blank labels will also be provided for any additional labeling needs. Use a permanent waterproof marker for any handwritten labeling needs. Each vacutainer used for phlebotomy and urine collection must be labeled appropriately with the Participant ID, collection date, and collection tube type.

### 5.13.3 Blood Collection

As described in Section 5.13.1, the order of blood draw will be:

· 2x 5.0ml SST gold top (BD 367986)
· 2x 6.0ml purple (EDTA) top (BD 367863)
· 1x 3.0ml purple (EDTA) top (BD 367856)
· 2x sodium heparin CPT tubes (BD 362753 )
· 1x Paxgene RNA (BD 762165) – keep upright after inverting/mixing

**These tubes must be organized and labeled prior to the participant’s visit, as described in Sections 5.13.1 and 5.13.2**.

#### IMPORTANT NOTE

Each phlebotomy vacutainer should be allowed to fill completely inverted 8-10 times immediately after collection to ensure complete mixing and to prevent clots in the anti-coagulated tubes. Any vacutainers that are not able to be filled to capacity should be processed as per protocol and clearly notated in REDCap. Any tubes not filled must be indicated in REDCap. Use the Specimen Management form to document the blood draw and sample processing.

Participants will have blood drawn according to procedures implemented at each data site.

### 5.13.4 Urine Collection

A midstream urine sample will be collected from each participant and stored according to procedures established at each Data site. Urine will be collected using a BD Urine Collection kit. Urine samples should be frozen immediately after processing and stored at -80°C until batch-shipping to UW’s NORC lab.

### 5.13.5 Safety Considerations

The risks of blood collection are minimal. Some participants may feel lightheaded or dizzy after blood collection and will be offered rest, fluids, and snacks. All collection tubes will be handled and disposed of according to established guidelines.

## 5.14 Biospecimen Processing, Storage, and Shipment

This chapter covers AI-READI biospecimen processing for both clinical lab testing and biorepository storage.

### 5.14.1 Overview

Figure 6 (below) outlines the overall scheme for biospecimen processing, storage, and shipping.

**Figure 6.**
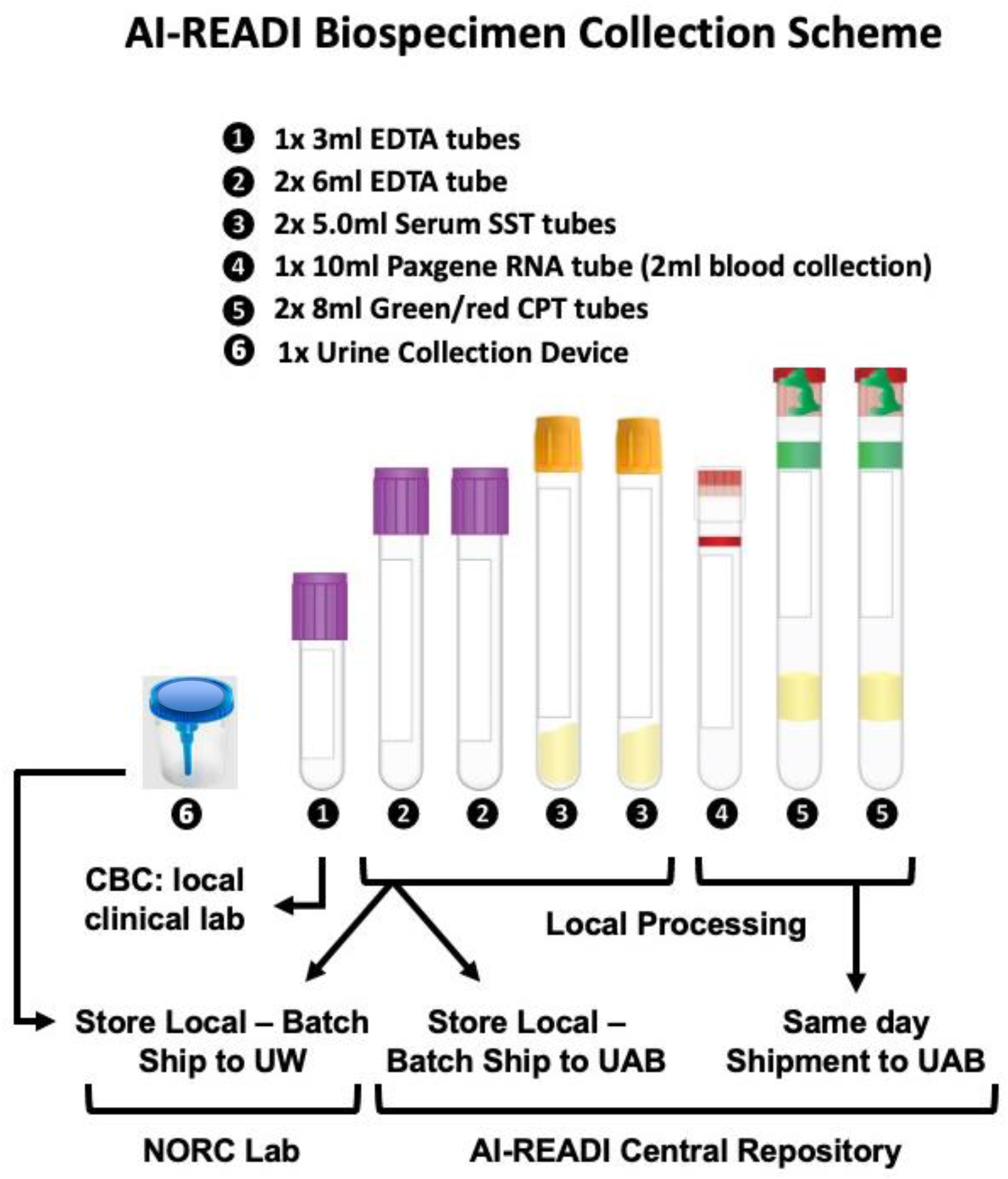
The collection and processing of biospecimens for AI-READI will involve blood and urine collection, local processing of samples, and daily and batch shipping of samples to UAB (central repository) and UW (NORC lab - clinical testing).

### 5.14.2 Preparation of Vials for Specimen Processing and Storage

This section describes the preparation and labeling of all tubes to be used in biospecimen processing and storage. Biospecimen processing refers to all steps of processing after specimen collection and before storage at -80°C until batch shipping on dry ice to UAB or UW.

For each participant, the following **storage tubes** should be prepared with labels:

- 2x 0.6ml plasma storage – 0.75 Micronic tube (Micronic, Lelystad, AR)
  - **Blue** color caps will be used
- 4x 0.4ml plasma storage – 0.75 Micronic tube
  - **Purple** color caps will be used
- 2x 0.6ml serum storage – 0.75 Micronic tube
  - **Red** color caps will be used
- 4x 0.4ml serum storage – 0.75 Micronic tube
  - **Orange** color caps will be used
- 2x buffy coat – 2.0 Micronic tube
  - **Gray** color caps will be used
- 4x 2mL ThermoFisher storage tubes
  - Clear caps (ThermoFisher) with O-rings will be used

#### Each of these tube types will be stored in separate boxes

Detailed box layouts by participant ID have been provided to each Data site.

#### Specimen storage tubes summary

- 0.75mL Micronic pre-barcoded tubes used for 0.6ml and 0.4ml plasma or serum aliquots for biorepository storage
- 2.0mL Micronic pre-barcoded tubes for buffy coats (up to 1.5mL) for biorepository storage
- 2mL Thermo-Fisher storage tubes used for plasma, serum, whole blood and urine for UW NORC analysis.

Micronic tubes: These tubes are pre-coded with barcodes (bottom and side) that will be used to identify each individual tube. In addition, preprinted labels will be provided that contain the participant ID and the specimen type. These preprinted labels will be applied to the side of the tube opposite the barcode.

2ml Fisher storage tubes: Preprinted labels will be provided containing the participant ID, the specimen type and a 1D barcode. These labels will be applied to the side of the tube with the barcode in a horizontal position.

A. **Plasma and Serum mixing tube** (to allow for combining the plasma and the serum from the respective 2 collection tubes into single plasma and serum aliquots to ensure complete mixing). · A single tube will be used to mix plasma or serum from the multiple vacutainers prior to specimen aliquoting for storage. A 5mL snap cap Eppendorf tube or a 15mL screw top conical tube (various sources available) can be used to pool plasma or serum prior to aliquoting into storage tubes. 1 plasma + 1 serum = 2 tubes per participant UAB to provide pre-printed rolls of labels (0.75” x 0.4” in size)
  · Example (left shows fields to be included on the label/right shows completed example):

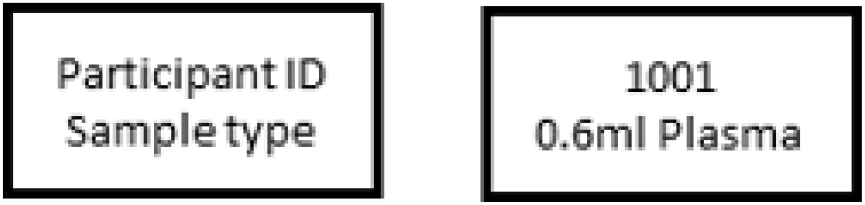
B. **Micronic Aliquot tubes for UAB biorepository** Each Micronic tube will contain a unique barcode (in 2D format on the bottom and 1D format on the side), together with the corresponding human readable barcode number on the side. In addition, UAB will provide preprinted freezer compatible labels (x” x y”) that can be affixed to the side of these tubes (not covering the barcode area). These labels will contain the participant ID (10 labels per participant) for 1400 participants. Blank labels will also be provided for any additional labeling needs. Use a permanent waterproof marker for any handwritten labeling needs. Example tube: 6 plasma + 6 serum + 2 buffy coats = 14 tubes per participant UAB to provide pre-printed rolls of labels (0.75” x 0.4” in size)
  · Example:

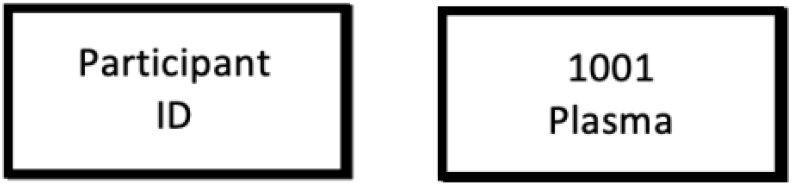
C. **UW NORCAliquot tubes** UW Storage tubes: UAB will provide freezer compatible labels for the storage vials and storage boxes being shipped to the UW NORC. Example storage tube 4 tubes per participant UAB to provide pre-printed rolls of labels (1” x 0.5” in size)
  · Example: (note – Barcode is site + ID – eg UW1001 per Andy’s request)

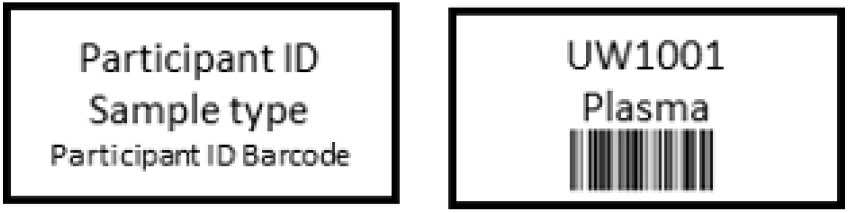

### 5.14.3. Biospecimen Processing Instructions

This section details the processing steps to be performed on all blood and urine sample prior to storage and shipping. Figures 7-9 provide overviews of processing for the different types of samples collected under the AI-READI protocol.

**Figure 7.**
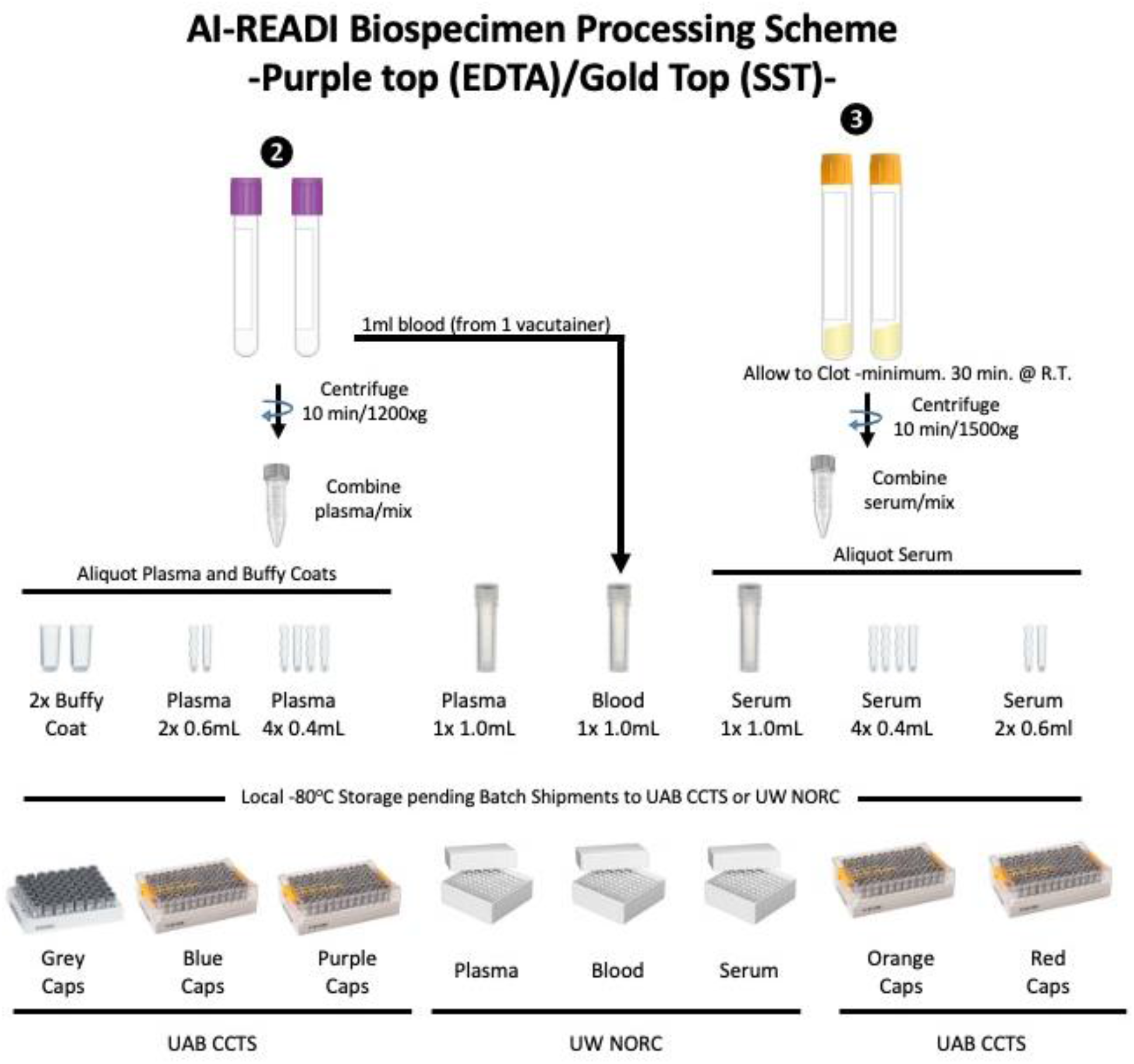
Processing of 6 ml EDTA and 5 ml SST vacutainers.

**Figure 8.**
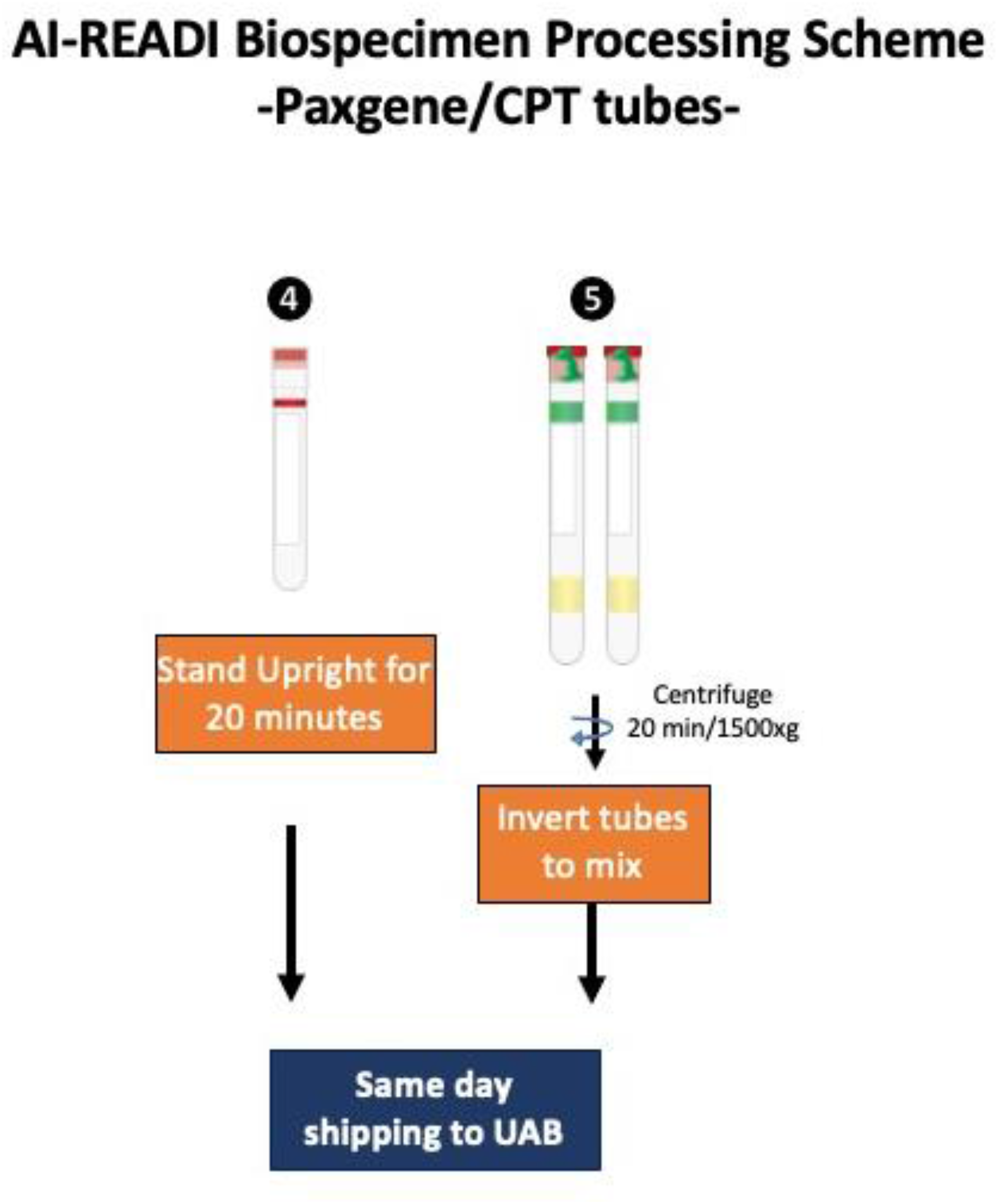
PAXgene RNA tubes and CPT tubes require special handling and same-day ambient shipping to UAB CCTS.

**Figure 9.**
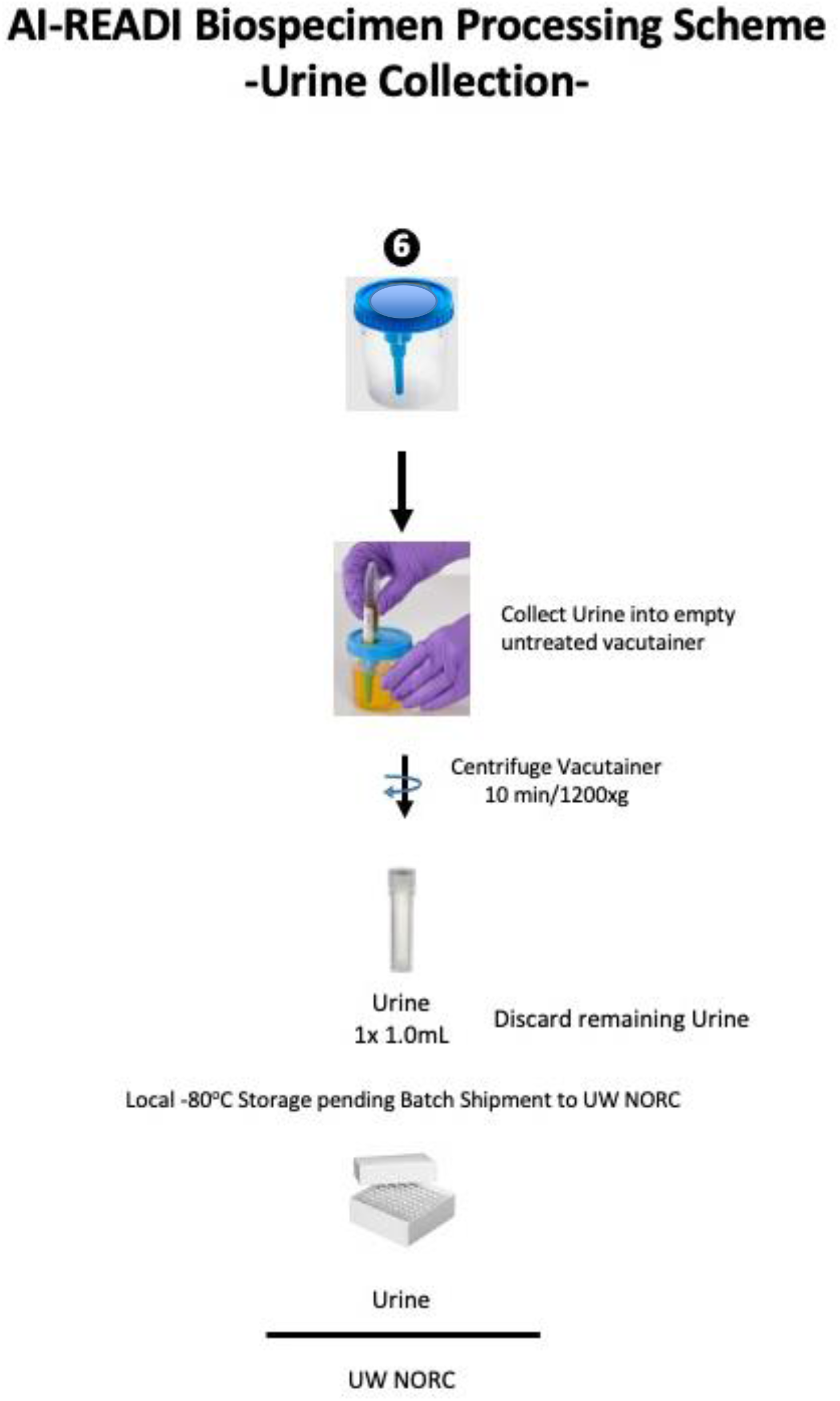
Urine is collected, centrifuged, and aliquots stored at -80°C until batch shipment to UW (NORC Lab for clinical testing).

#### Detailed Blood/Urine Processing Instructions

Each vacutainer is to be processed as soon as possible after collection (within 2 hours whenever possible). It is important to ensure that the Paxgene tube is kept upright after collection. If processing cannot be initiated within 2 hours of collection, the urine and purple/top specimens should be refrigerated. The gold top/SST, CPT and Paxgene tubes should not be refrigerated. All centrifugation should be performed at room temperature in swinging bucket rotors. Care should be taken to ensure that tubes are balanced in the centrifuge prior to centrifugation. Processing that is initiated 2 hours or more after collection will be considered a deviation and should be appropriately noted on the participant’s collection sheet and the specimen logs.

1. **Gold top/SST vacutainers** should be held at room temperature for a minimum of 30 minutes to allow for complete clotting of the blood. Tubes should then be centrifuged:
  · Gold top/SST centrifugation: 1500xg (RCF) for 10 minutes at room temperature
    ∘ Eppendorf 5702: 1500xg = XXXX RPM
    ∘ Beckman XXXX
2. **Purple top vacutainers (6ml size)** should be centrifuged as soon as possible after collection (note that 1.0ml of whole blood should be removed from one of the vacutainers prior to centrifugation – ensure that the vacutainers are appropriately balanced prior to centrifugation):
  · 6ml purple/EDTA centrifugation: 1200xg (RCF) for 10 minutes at room temperature
    ∘ Eppendorf 5702: 1200xg = XXXX RPM
    ∘ Beckman XXXX

#### Buffy Coat Isolation

Using a transfer pipet, remove the concentrated leukocyte band (this is the buffy coat), plus a small portion of the plasma using a circular motion with the tip of the pipet at the top of the buffy coat layer. Inclusion of a small number of erythrocytes at the buffy coat erythrocyte interface is acceptable to facilitate complete harvest of the buffy coat layer. The total volume isolated is typically 0.5-1.0mL. The absolute volume is not critical (but do not exceed 1.5mL) and it is more important to ensure complete harvest of the buffy coat.

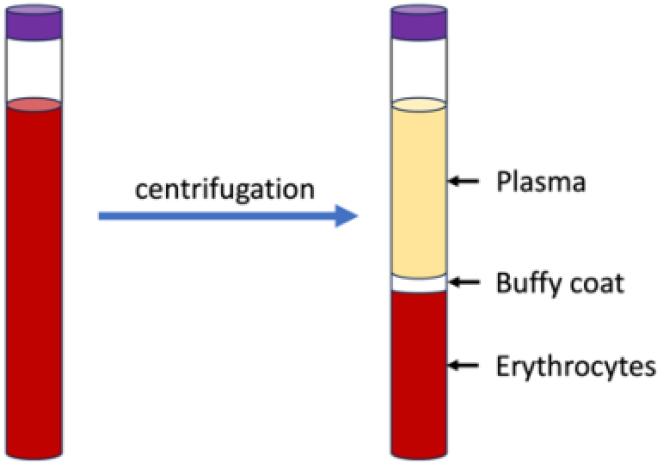

3. **Purple top vacutainer (3ml size)** is intended for local CBC determination. Each data site will use instructions provided by the local testing lab for the submission of fresh blood for CBC testing.
4. **CPT vacutainers** should be centrifuged as soon as possible after collection. These tubes must be kept at room temperature:
  · CPT centrifugation: 1500xg (RCF) for 20 minutes at room temperature. The upper layer containing the plasma and the peripheral blood mononuclear cells (PBMC) at the gel interface should be free of red blood cells (RBC). If RBCs are still present in the upper layer, the sample can be centrifuged for another 4 minutes. Additional details regarding the handling of CPT tubes can be found at: https://www.bdbiosciences.com/content/dam/bdb/products/global/blood-collection/blood-collection-tubes/362753_base/pdf/VDP40393.pdf
  · After centrifugation, the tubes are inverted to resuspend the cells and then shipped to UAB on the day of collection
    ∘ Eppendorf 5702: 1500xg = XXXX RPM using XXX buckets
    ∘ Beckman
    ∘ **NOTE** – these tubes are taller than most vacutainers and require special buckets to accommodate their height. Failure to follow these instructions can result in tube breakage during centrifugation.
5. **Paxgene RNA vacutainer** must remain at room temperature (18°C – 25°C) for a minimum of 2 hours (maximum of 72 hours). These tubes are then shipped ambient to UAB on the day of collection. Complete product Paxgene RNA tube information and instructions can be found at https://www.bdbiosciences.com/content/dam/bdb/products/global/blood-collection/blood-collection-tubes/762165_base/pdf/VDP40400.pdf.

#### Aliquoting Instructions for blood/plasma/serum (purple top and SST vacutainers)

Figure 7 outlines the plasma and serum aliquoting needs. Specimens for clinical lab testing (UW NORC) will be aliquoted in 2ml storage tubes (Fisher catalog #s 02-681-343 (tubes) and 02-681-358(caps)) and placed in cardboard freezer boxes (03-391-515 or equivalent). **IMPORTANT NOTE: clinical samples to be sent to the UW NORC lab should be organized by sample type. Serum, plasma, whole blood, and urine samples should be placed in separate boxes that are clearly labeled**. Please see below for instructions on maintaining proper spreadsheet logs of clinical lab testing samples. Each site is responsible for properly labeling, storing, and logging their participant samples.

Specimens for biobanking will be aliquoted into 0.75ml Micronic storage tubes (plasma/serum) or 2.0ml Micronic storage tubes (buffy coats) and placed into Micronic boxes (96 tube format for plasma/serum or 48 tube format for buffy coats). Detailed Micronic box layouts are supplied in section 5.14.7 (below). Each participant has designated tube locations in each storage box that must be followed. If an aliquot cannot be made due to insufficient material, the corresponding aliquot tube location should be left empty.

Order of processing should be:

1. 1.0 ml of mixed purple top/EDTA whole blood from the first EDTA vacutainer should be added to a 2ml Fisher storage tube prior to centrifugation.
2. After centrifugation of the purple top/EDTA tubes, the following aliquots are prepared:
  a. Transfer plasma, the upper aqueous portion from the tube, from each Purple-top vacutainer into the labeled plasma transfer tube (5ml Eppendorf or 15mL conical tube).
  b. Cap transfer tube and mix by inverting.
  c. Using a micropipette, prepare aliquots (in this order:1.0ml into the Fisher storage tube for the UW NORC; 4x 0.4mL and 2×0.6mL into the appropriate Micronic tubes).
  d. Transfer the Micronic tubes into the correct Micronic storage box (see Micronic box layouts).
  e. Transfer the UW NORC tube into the correct cardboard storage box (see UW NORC box layouts). Residual plasma can be saved by each site if space permits for IRB approved future usage.
    • 4x ∼0.4ml plasma from the first vacutainer into Micronic tubes in the designated Micronic box
    • 2x ∼0.6 ml plasma from the second vacutainer into Micronic tubes in the designated Micronic box
    • 1x 1.0 ml plasma from the second vacutainer into a Fisher storage tube for the UW storage box. **Place in a storage box that will only contain plasma samples**.
    • *Residual plasma - TBD*
    • buffy coats from each vacutainer into separate 2.0ml Micronic tubes in the designated Micronic box
3. After centrifugation of the gold top/SST tubes, the following aliquots are prepared:
  a. Transfer serum, the upper aqueous portion from the Gold-top tube, from each Gold-top vacutainer into the labeled serum transfer tube (5ml Eppendorf or 15mL conical tube).
  b. Cap transfer tube and mix by inverting.
  c. Using a micropipette, prepare aliquots (in this order:1.0ml into the Fisher storage tube for the UW NORC; 4x 0.4mL and 2×0.6mL into the appropriate Micronic tubes).
  d. Transfer the Micronic tubes into the correct Micronic storage box (see Micronic box layouts).
  e. Transfer the UW NORC tube into the correct cardboard storage box (see UW NORC box layouts). Residual serum can be saved by each site if space permits for IRB approved future usage.
    • 4x ∼0.4ml serum from the first vacutainer into Micronic tubes in the designated Micronic box
    • 2x ∼0.6 ml serum from the second vacutainer into Micronic tubes in the designated Micronic box
    • 1x 1.0 ml serum from the second vacutainer into a Fisher storage tube for the UW storage box. **Place in a storage box that will only contain serum samples**.
    • *Residual serum is to be disposed of by the data site following institutional policy*.
4. **Urine** is collected in a sterile urine collection bottle by the participant. Conical tubes (empty untreated vacutainers) are attached to the collection bottle and urine is transferred by inversion. The urine is centrifuged at 1200xg for 10 minutes to remove particulates and one ml of clarified urine is transferred to a 2 ml. labeled storage tube (Fisher catalog #s 02-681-343 (tubes) and 02-681-358(caps)). Urine is stored at -80°C until batch shipment to the UW NORC Lab. Place urine samples into a properly labeled freezer storage box that **only contains urine samples** and log into the sample spreadsheet as discussed below.

#### NOTE

There will be 3 different anticipated types of specimen shipping, as described in sections 5.14.4, 5.14.5, and 5.14.6). Shipping of human biospecimens should only be performed by individuals who have completed appropriate IATA training for the shipping of Biological Substance, Category B (diagnostic specimens).

Micronic tubes containing biospecimens to be sent to UAB’s biorepository will be logged according to barcode and maintained in spreadsheet format as described in Section 5.14.7.

### 5.14.4 Specimen Inventory Tracking

Serum, plasma, and urine aliquots to be batch-shipped to UW’s NORC lab are to be entered into a spreadsheet (see below). Precise organization of samples is imperative. Specimens destined for the NORC lab are to be stored according to specimen type and the spreadsheet must be supplied to the NORC lab upon shipment. **Serum, plasma, whole blood, and urine samples must be stored in separate freezer boxes and logged accordingly**. Please see below for an example of the spreadsheet format. Each site may make a copy of the spreadsheet format from a master copy shared with the site managers. The identity and location of each specimen aliquot tube needs to be recorded in the provided spreadsheet templates.

**For the UW NORC aliquots** (in 2mL Fisher storage tubes), the following data points need to be included:

- Box name
- Site (UW, UCSD or UAB)
- Participant ID (4 digit assigned number)
- Sample Type (plasma, serum, blood, urine)

NORC sample spreadsheet: example provided below:

Box position (1-81 within each box)

| A | B | C | D | E | F | G | H | I | J | K | L | M |
| --- | --- | --- | --- | --- | --- | --- | --- | --- | --- | --- | --- | --- |
| Sample Information |  |  |  |  |  |  |  |  |  | NORC Analysis |  |  |
| Box Label | Site | Participant ID | Sample Type | Box position | Barcode | volume (ml) | Date of Collection | Date of Processing | Time Deviation | BNP (pg/mL) | C-Peptide (ng/mL) | Notes: |
| UW EDTA Plasma Box 1 | UW |  | EDTA Plasma | 1 |  |  |  |  |  |  |  |  |
| UW EDTA Plasma Box 1 | UW |  | EDTA Plasma | 2 |  |  |  |  |  |  |  |  |
| UW EDTA Plasma Box 1 | UW |  | EDTA Plasma | 3 |  |  |  |  |  |  |  |  |
| UW EDTA Plasma Box 1 | UW |  | EDTA Plasma | 4 |  |  |  |  |  |  |  |  |
| UW EDTA Plasma Box 1 | UW |  | EDTA Plasma | 5 |  |  |  |  |  |  |  |  |
| UW EDTA Plasma Box 1 | UW |  | EDTA Plasma | 6 |  |  |  |  |  |  |  |  |
| UW EDTA Plasma Box 1 | UW |  | EDTA Plasma | 7 |  |  |  |  |  |  |  |  |
| UW EDTA Plasma Box 1 | UW |  | EDTA Plasma | 8 |  |  |  |  |  |  |  |  |
| UW EDTA Plasma Box 1 | UW |  | EDTA Plasma | 9 |  |  |  |  |  |  |  |  |
| UW EDTA Plasma Box 1 | UW |  | EDTA Plasma | 10 |  |  |  |  |  |  |  |  |
| USCD EDTA Plasma Box | SD |  | EDTA Plasma | 1 |  |  |  |  |  |  |  |  |
| UAB EDTA Plasma Box 1 | UA |  | EDTA Plasma | 1 |  |  |  |  |  |  |  |  |

**For Micronic tubes**, the following data points need to be included:

- Well (A1 through H12)
- Scanned barcode
- Participant ID
- Sample Type
- volume (ml) – except for buffy coat that are recorded as 1 each.
- Date of Collection
- Date of Processing
- Time Deviations Specimen type

Micronic samples spreadsheet: example shown below:

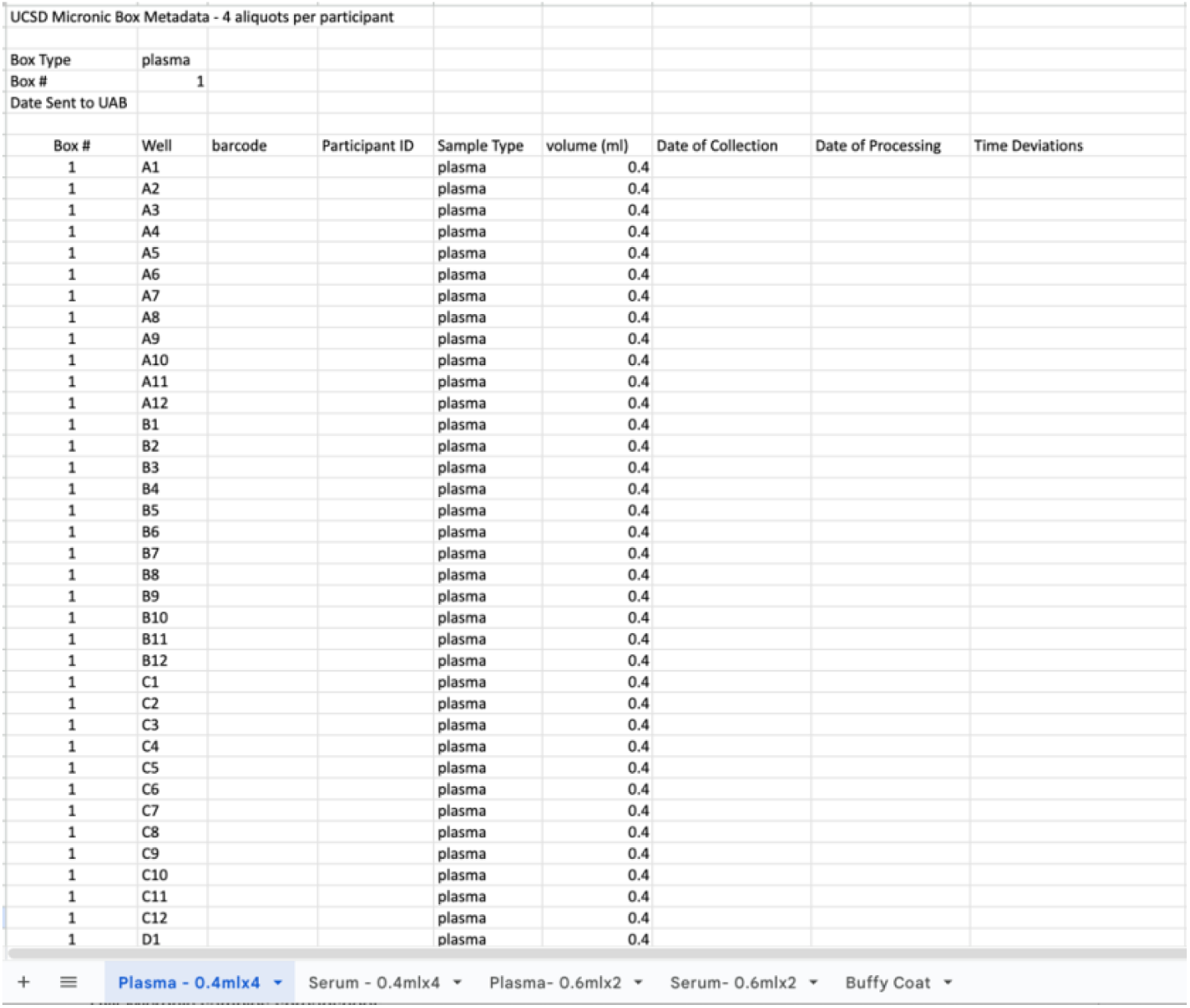

### 5.14.5 Storage Boxes: Labeling, Preparation, Loading

#### A. Specimen Storage Boxes Summary

- 96 tube format boxes (plastic): used to hold 0.75mL Micronic pre-barcoded tubes used for 0.6ml and 0.4ml plasma or serum aliquots
- 48 tube format boxes (plastic): used to hold 2.0mL Micronic pre-barcoded tubes for buffy coats (up to 1.5mL)
- 2” tall standard freezer rack boxes (cardboard): used to hold 2mL Thermo-Fisher storage tubes containing specimens for UW NORC analysis. Separate boxes will be used for each specimen type (plasma, serum, whole blood and urine).

## B. Micronic Tube Freezer Storage Boxes

Each Micronic box contains a unique 1D barcode on the side of the box. In addition, UAB will provide labels containing a box identifier (specimen type and box #) to be applied to a side of each box prior to storage to facilitate box identification in the freezer.

UAB to provide pre-printed rolls of labels for each box (note that one side of the box is already barcoded for tracking purposes (2.125” x 0.27” in size)

Boxes will store plasma, serum and buffy coats

· 2 labels per box will be provided to allow for labeling of both the top and bottom of the box for ease in identification.
· Example:

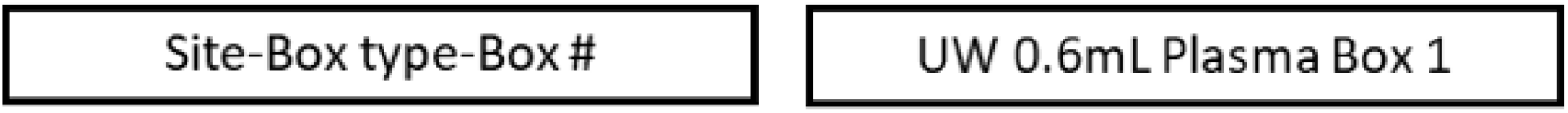

## E. UW NORC Freezer Storage Boxes

Important: Each Data site will use separate freezer boxes for plasma, EDTA blood, serum and urine for NORC analysis

UAB to provide pre-printed label sheets (same as vacutainer labels)

· 4 labels per box will be supplied to allow labeling of both the top and bottom of boxes (pending further input from Andy regarding placement of box labels)
· Example:

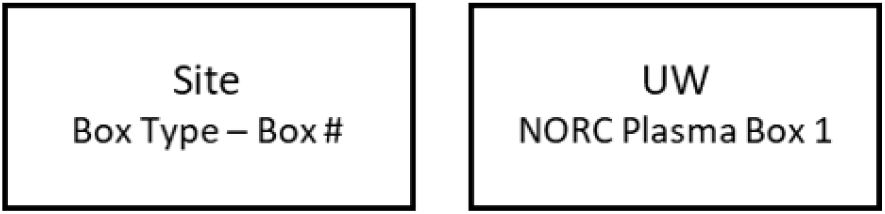

**Note: All storage boxes must be properly inventoried using the spreadsheets provided to the site managers at each data collection site**.

### 5.14.6 Biospecimen Inventory Tracking (UAB CCTS)

Weekly receipts of biospecimens will be provided by UAB CCTS and shared with all project team members. Biorepository inventory will be made available on fairhub.io and accessible under the controlled access database.

### 5.14.7 Same Day Shipping of PAXGene and CPT Tubes

#### Ambient (no ice) Day of Collection

shipping of CPT and Paxgene tubes are shipped to the UAB CCTS. FedEx Clinical Paks should be used for Priority Overnight service. Do not request Early Morning Delivery or Saturday Delivery.

Please use the following procedure for daily shipments:

1. Provide sample and tracking information in the Biospecimen Shipping log. Each site has their own spreadsheet to track shipments.
2. Send an email notification to staff at the UAB CCTS to expect a shipment
3. Ambient Shipments should be sent to the UAB Center for Clinical and Translational Science (CCTS) using the address provided to the data manager at each site.
  a. NOTE for Day of Collection Shipping– if the day of collection shipping is not possible, then the blood tubes should be shipped next day. If a Friday collection occurs too late in the day for same day shipping, every effort should be made to tendering the package to a FedEx office on Saturday for Monday delivery. **NOTE**: only certain specified FedEx offices accept Category B specimens, some are inconveniently located and may have different hours.
  b. Suggested shipping container is Fisher 03-530-35 (Sonoco 409). One per patient will be needed. Follow FedEx instructions for packaging the shipping containers into one appropriately sized FedEx Clinical Pak shipper. Each Data site will need to determine the shipping containers and packaging material that works best for their purposes, based on their participant recruitment and visit schedule. (https://www.fedex.com/en-us/shipping/how-to-ship-clinical-samples.html#1). FedEx will usually provide their shipping supplies at no cost to account holders.

## 5.14.8 Batch Shipment to UW NORC Lab and Transfer of Inventory Data and Deviations

This section describes the shipping of processed and frozen plasma, serum and whole blood to the UW data team for NORC analysis. FedEX Priority Overnight service should be requested. Do not request Early Morning Delivery or Saturday Delivery. **Note**: shipments should only be sent on Mondays-Wednesdays to ensure proper delivery when receiving staff is available.

Freezer boxes containing aliquot samples should be placed in one of 3 types of dry ice shippers (see Table 8), based on the number of boxes to be shipped. Each site will determine their shipping needs based on their local storage capacities and frequency needs for shipment. The boxes must be packed with dry ice to ensure that samples remain frozen during shipment. A hardcopy of the spreadsheet with sample information should be included with the shipment.

**Table 7.** Recommended shipping supplies for daily ambient shipments of CPT and PaxGene tubes to UAB CCTS.

| Items | Catalog Number | Mfr. No. | Capacity |
| --- | --- | --- | --- |
| **Specimen Tube Mailers**<br>**(CPT/Paxgene RNA tubes)** | Fisher 03-530-35 | Therapak™ 409 | 7, 10, 15 or 25 mL |
| **Absorbent Sheet** | Fisher 22-130-039 | Therapak™ 10300 | Absorbent 3x4" - 50 mL capacity |
|  | Fisher 22-130-040 | Therapak™ 10306 | Absorbent 3x6" - 120 mL capacity |
| **Biohazard Specimen Transport Bags** | Fisher 22-311-100 | Minigrip LLC SBL2X810B5 | 10 x 8 in. (25.4 x 20.32cm) |
| **Packing Tape** | Staples 886404 | Any sealing packing tape |  |

**Table 8.**
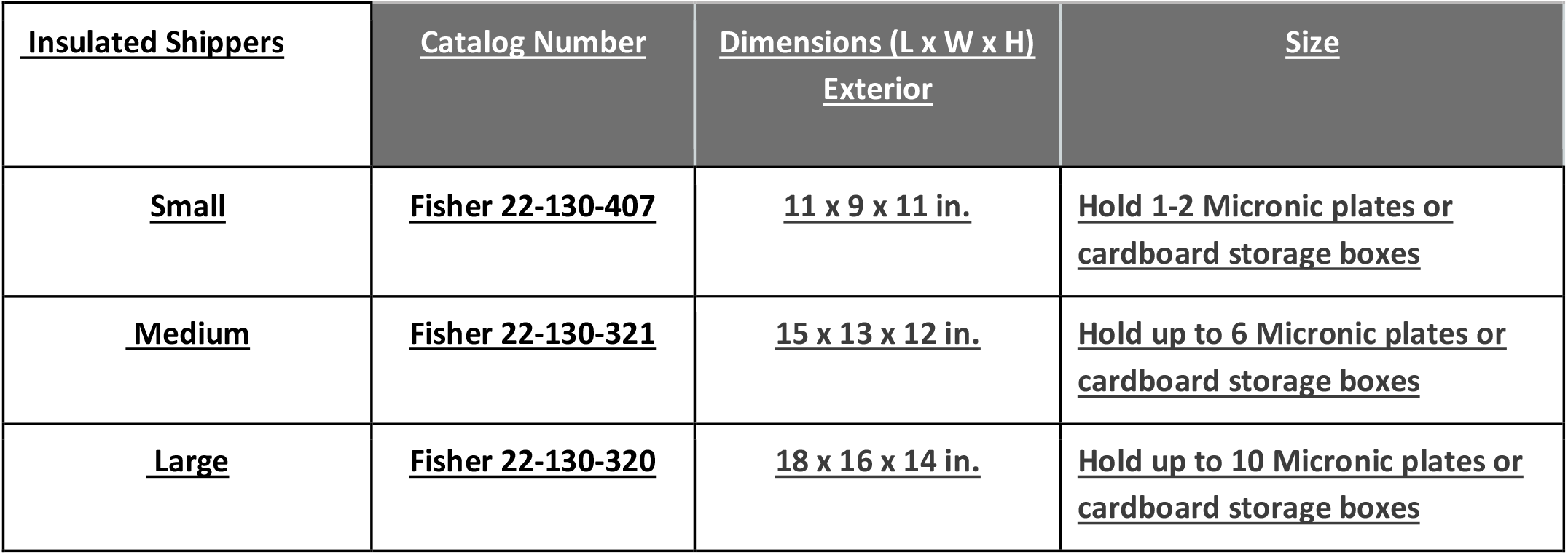
Recommended shipping supplies for biospecimen batch shipments on dry ice.

Please follow the procedure below for batch shipments to UW for NORC lab analysis:

1. Provide sample and tracking information in the Biospecimen Shipping log. Each site has their own spreadsheet to track shipments
2. Call the UW data manger the day before the shipment is sent and let them know to expect your packages.
3. Ship to the UW Medicine Karalis Johnson Retina Center using the specific address provided to each site manager.

Frequency of shipment will be dictated by local storage limitations. **Only full specimen boxes should be shipped**.

Once received by the CRU at UW, the sample boxes will be unloaded from the shipping containers and stored in a -80°C freezer until delivery to the NORC lab is arranged (described in more detail below in Section 5.14.6). **IMPORTANT**: It is each Data site’s responsibility to be sure that the samples are properly labeled and organized in boxes, and that the organization matches the spreadsheet that accompanies each shipment (described in detail in Section 5.14.3). UW is not responsible for the quality control of batch-shipped samples. They will simply unload and store at -80°C until testing.

### Pre-shipment at sites

- Ensure all boxes are packed correctly with sufficient amounts of dry ice to keep samples frozen
- Ensure that all samples are properly logged into the sample spreadsheet and that the spreadsheet is forwarded to supervising staff in the NORC lab
- Ensure a copy of the spreadsheet of samples is in the shipping box.
- Email the site manager at UW to notify a box is full and **schedule a date** for shipping out the boxes. **Note**: shipments should only be done M-W for overnight delivery. Do not ship without first arranging with Cari and Isha at UW. Staff work a hybrid schedule and shipments must be scheduled in advance
  - Be mindful of hot weather and anything else that could disrupt shipment.
- Log the date shipped, shipping label tracking number, and planned delivery date into REDCap so that notifications may be sent to update UW staff.

## 5.14.9 UW CRU Sample Box Receiving Protocol (for NORC samples)

### Receiving sample boxes at CRU

- UW CRU will receive samples from FedEx.
- UW CRU will track on the Sample Shipment Log:
  - Date received
  - Which institution the boxes are from
  - Which boxes came in
  - Participant ID numbers received
- After tracking samples in the sample shipment log, CRU staff will initial and place boxes in designated spot in freezer.

#### 5.14.10 Batch Shipment to UAB CCTS (Central Repository) and Transfer of Inventory and Deviations

This section describes the shipping of processed and frozen plasma, serum and buffy coats in Micronic boxes to UAB. **Only full specimen boxes should be sent**. Each box should be placed into a reclosable bubble bag (such as ULINE S-5098) to prevent breakage during shipment. FedEX Priority Overnight service should be requested. Do not request Early Morning Delivery or Saturday Delivery. **Note**: shipments should only be sent on Mondays-Wednesdays to ensure proper delivery when receiving staff is available.

Freezer boxes containing aliquot samples should be placed in one of 3 types of dry ice shippers (see Table 9), based on the number of boxes to be shipped. Each site will determine their shipping needs based on their local storage capacities and frequency needs for shipment. The boxes must be packed with dry ice to ensure that samples remain frozen during shipment. A hardcopy of the spreadsheet with sample information should be included with the shipment. Frequency of shipment will be dictated by local storage limitations.

**Table 9.**
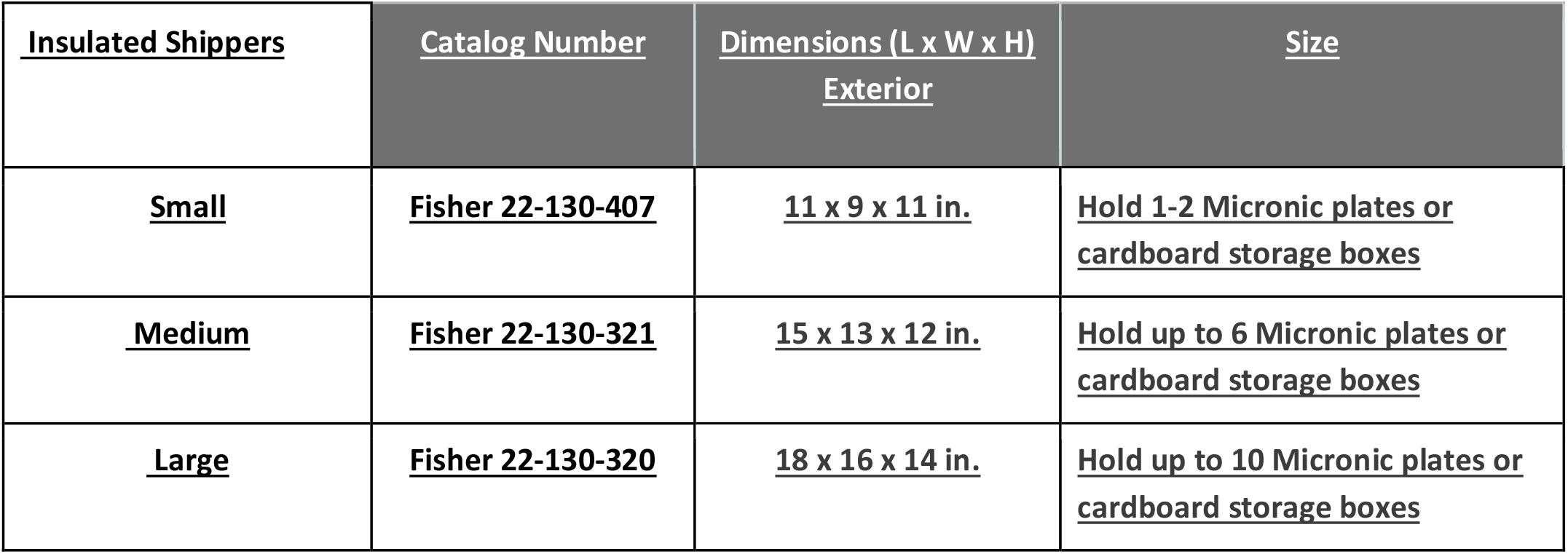
Recommended shipping supplies for micronic tube samples that are batch-shipped to UAB on dry ice.

Please follow the procedure below for batch shipments to UAB’s CCTS:

1. Provide sample and tracking information in the Biospecimen Shipping log. Each site has their own spreadsheet to track shipments.
2. Send an email notification to CCTS staff using the contacts provided to each site’s data manager.
3. Batch shipments should be sent to the address provided to each site’s data manager.

#### 5.14.11 UAB Biobanking Procedures and Inventory

Micronic tube plasma, serum and buffy coat aliquots: As aliquots are prepared, a box specific spreadsheet will be maintained that will document the physical box location of each aliquot, the barcode associated with each aliquot tube (to be scanned from the tube directly into the spreadsheet), the participant ID, the aliquot volume (in ml), the date of collection, the date of processing (which should be the same as the date of collection) and any deviations in the time that elapsed between collection and processing (more than 2 hrs). If an aliquot is not prepared due to insufficient blood, note the volume as “0”, leave date of processing blank and not insufficient volume in the deviation column. Examples of biobanking manifests are provided below.

##### Plasma/Serum – 4x 0.4ml aliquots per participant

| Micronic Box Metadata - 4 aliquots per participant |  |  |  |  |  |  |  |  |
| --- | --- | --- | --- | --- | --- | --- | --- | --- |
| Box Type | plasma (or serum) |  |  |  |  |  |  |  |
| Box # | 1 |  |  |  |  |  |  |  |
| Date Sent to UAB |  |  |  |  |  |  |  |  |
|  | Well | barcode | Participant ID | Sample Type | volume (ml) | Date of Collection | Date of Processing | Time Deviations |
| 1 | A1 | 7515128983 | 1001 | plasma | 0.4 | 1/17/2023 | 1/17/2023 |  |
| 2 | A2 | 7515128984 | 1001 | plasma | 0.4 | 1/17/2023 | 1/17/2023 |  |
| 3 | A3 | 7515128985 | 1001 | plasma | 0.4 | 1/17/2023 | 1/17/2023 |  |
| 4 | A4 | 7515128986 | 1001 | plasma | 0.4 | 1/17/2023 | 1/17/2023 |  |
| 5 | A5 | 7515128876 | 1002 | plasma | 0.4 | 1/18/2023 | 1/18/2023 | processed 3 hrs after collection |
| 6 | A6 | 7515128877 | 1002 | plasma | 0.4 | 1/18/2023 | 1/18/2023 | processed 3 hrs after collection |
| 7 | A7 | 7515128878 | 1002 | plasma | 0.4 | 1/18/2023 | 1/18/2023 | processed 3 hrs after collection |
| 8 | A8 | 7515128879 | 1002 | plasma | 0.4 | 1/18/2023 | 1/18/2023 | processed 3 hrs after collection |

##### Plasma/Serum – 2x 0.6ml aliquots per participant

Plasma/Serum – 2x 0.6ml aliquots per participant:
| Micronic Box Metadata - 4 aliquots per participant |  |  |  |  |  |  |  |  |
| --- | --- | --- | --- | --- | --- | --- | --- | --- |
| Box Type | plasma (or serum) |  |  |  |  |  |  |  |
| Box # | 1 |  |  |  |  |  |  |  |
| Date Sent to UAB |  |  |  |  |  |  |  |  |
|  | Well | barcode | Participant ID | Sample Type | volume (ml) | Date of Collection | Date of Processing | Time Deviations |
| 1 | A1 | 7515128997 | 1001 | serum | 0.6 | 1/17/2023 | 1/17/2023 |  |
| 2 | A2 | 7515128996 | 1001 | serum | 0.6 | 1/17/2023 | 1/17/2023 |  |
| 3 | A3 | 7515128848 | 1002 | serum | 0.6 | 1/18/2023 | 1/18/2023 | processed 3 hrs after collection |
| 4 | A4 | 7515128849 | 1002 | serum | 0.6 | 1/18/2023 | 1/18/2023 | processed 3 hrs after collection |

##### Buffy coats– 2 aliquots per participant

Buffy coats– 2 aliquots per participant:
| Micronic Box Metadata - 4 aliquots per participant |  |  |  |  |  |  |  |
| --- | --- | --- | --- | --- | --- | --- | --- |
| Box Type | Buffy Coat |  |  |  |  |  |  |
| Box # |  |  |  |  |  |  |  |
| Date Sent to UAB |  |  |  |  |  |  |  |
|  | Well | barcode | Participant ID | Sample Type | Date of Collection | Date of Processing | Time Deviations |
| 1 | A1 | 4048286044 | 1001 | Buffy Coat | 1/17/2023 | 1/17/2023 |  |
| 2 | A2 | 4048286045 | 1001 | Buffy Coat | 1/17/2023 | 1/17/2023 |  |
| 3 | A3 | 4048286076 | 1002 | Buffy Coat | 1/18/2023 | 1/18/2023 | processed 3 hrs after collection |
| 4 | A4 | 4048286077 | 1002 | Buffy Coat | 1/18/2023 | 1/18/2023 | processed 3 hrs after collection |

Biorepository inventories will be reported periodically to fairhub.io as explained in Section 7.3.

### 5.15 Clinical Lab Results

#### 5.15.1 Lab Testing Requirements and Standardization

The clinical lab tests performed for all 3 data sites will yield results with standardized units. All testing, with the exception of the CBC test, will be performed at the UW NORC lab. The CBC test will be performed locally at each site, but the units reported are standardized between sites. Table 10 (next page) summarizes all the reference intervals for all clinical testing performed under the AI-READI protocol and associated LOINC codes.

**Table 10.** Units and reference intervals for all lab tests performed on blood and urine samples.

| Test | Units | Reference Interval | LOINC |
| --- | --- | --- | --- |
| EDTA Plasma Tests: |  |  |  |
| NT-proBNP | pg/mL | Varies by age | 33762-6 |
| Troponin-T | ng/L | Female <11; Male <16 | 67151-1 |
| C-Peptide | ng/mL | 1.1-4.4 | 1986-9 |
| Insulin | ng/mL | 0.0-24.9 | 20448-7 |
| Serum Tests |  |  |  |
| CRP-HS | mg/L | 0.0-10.0 | 30522-7 |
| Total Cholesterol | mg/dL | <200 | 2093-3 |
| Triglycerides | mg/dL | <150 | 2571-8 |
| HDL Cholesterol | mg/dL | >39 | 2085-9 |
| LDL Cholesterol (calculated field) | mg/dL | <130 |  |
| Glucose | mg/dL | 62-125 | 2345-7 |
| BUN | mg/dL | 8.0-21.0 | 3094-0 |
| Creatinine | mg/dL | Female: 0.38-1.02, Male: 0.51-1.18 | 2160-0 |
| BUN/Creatinine ratio |  |  |  |
| Sodium | mEq/L | 135-145 | 2951-2 |
| Potassium | mEq/L | 3.6-5.2 | 2823-3 |
| Chloride | mEq/L | 98-108 | 2075-0 |
| Carbon Dioxide, Total | mEq/L | 22-32 | 2028-9 |
| Calcium | mg/dL | 8.9-10.2 | 17861-6 |
| Protein, total | g/dL | 6.0-8.2 | 2885-2 |
| Albumin | g/dL | 3.5-5.2 | 1751-7 |
| Globulin, total (calculated field) | g/dL |  |  |
| A/G Ratio (calculated field) |  |  |  |
| Bilirubin, Total | mg/dL | 0.2-1.3 | 1975-2 |
| Alkaline Phosphatase | IU/L | See table below | 6768-6 |
| AST | IU/L | 9-38 | 1920-8 |
| ALT | IU/L | Female: 7-33, Male Age 0-49: 10-64, Male Age 50+: 10-48 | 1742-6 |
| Whole Blood Tests |  |  |  |
| HbA1c | % | 4.0-6.0 | 4548-4 |
| WBC | x10E<sup>3</sup>/mL | 3.4-10.8 |  |
| RBC | x10E<sup>6</sup>/mL | 3.77-5.80 |  |
| Hemoglobin | g/dL | 11.1-17.7 |  |
| Hematocrit | % | 34.0-51.0 |  |
| MCV | fL | 79-97 |  |
| MCH | pg | 26.6-33.0 |  |
| MCHC | g/dL | 31.5-35.7 |  |
| RDW | % | 11.6-15.4 |  |
| Platelets | x10E<sup>3</sup>/mL | 150-450 |  |
| Urine Tests |  |  |  |
| Urine creatinine | ug/mL | N/A | 65634-8 |
| Urine Albumin | ug/mL | N/A | 34535-5 |

#### 5.15.2 Data Reporting and Data Transfer of Clinical Lab Results

NORC lab results will be returned to the Data Manager at UW on the same manifests used initially for storing, cataloging, and shipping biospecimens. The manifests are in .csv format. UW will upload clinical lab results to Azure Storage Explorer for all sites (explained in section 8.4.7).

#### 5.15.3 Reference Ranges for Clinical Lab Tests

The reference ranges for all clinical lab tests performed by the UW NORC lab (and CBC test performed at local labs for each Data site) are shown in Table 10 of Section 5.15.1. These reference intervals will be made available to participants when they are given access to their lab results (Appendix F). It is important that the Tools team is notified if a reference interval has changed, so that the dataset may be updated to reflect the new values.

## 6. Protocol Safety Considerations

CRCs are required to complete all training and certification components described in Sections 3.2 and 3.3. Training and certification is essential for CRC and participant safety, as well as proper execution of the protocol. CRCs will follow the sanitation procedures outlined in Section 5.2 prior to, during, and after all participant visits to minimize the potential spread of communicable disease. All staff who handle biospecimen samples must comply with guidelines outlined in the Bloodborne Pathogens and Shipping and Transporting Regulated Biological Samples training courses (Section 3.2), and properly dispose of all biological waste according to their institution’s guidelines.

All sites will keep a stock of snacks and beverages (water or juice) and will offer these to participants during the in-person visit. This is a long protocol and some participants will require snacks as part of their glucose management program. Snacks and drinks should always be offered after a participant’s blood is drawn. Chairs will be provided throughout the protocol and elevators should be available on routes involving travel from the CRU and other locations used for elements of the protocol. Disposable sunglasses will be offered to participants after dilation due to sensitivity to light, and participants will be warned that some activities (for example, driving) may be temporarily impacted by light sensitivity.

## 7. Variables, Data Entry, and Data Flow

### 7.1: Overview of Data Flow in AI-READI

AI-READI data will take different paths before final transfer to our data dissemination platform fairhub.io. This section provides a summary of the AI-READI data flow (depicted in the flow chart provided in Figure 10 on the next page).

**Figure 10:**
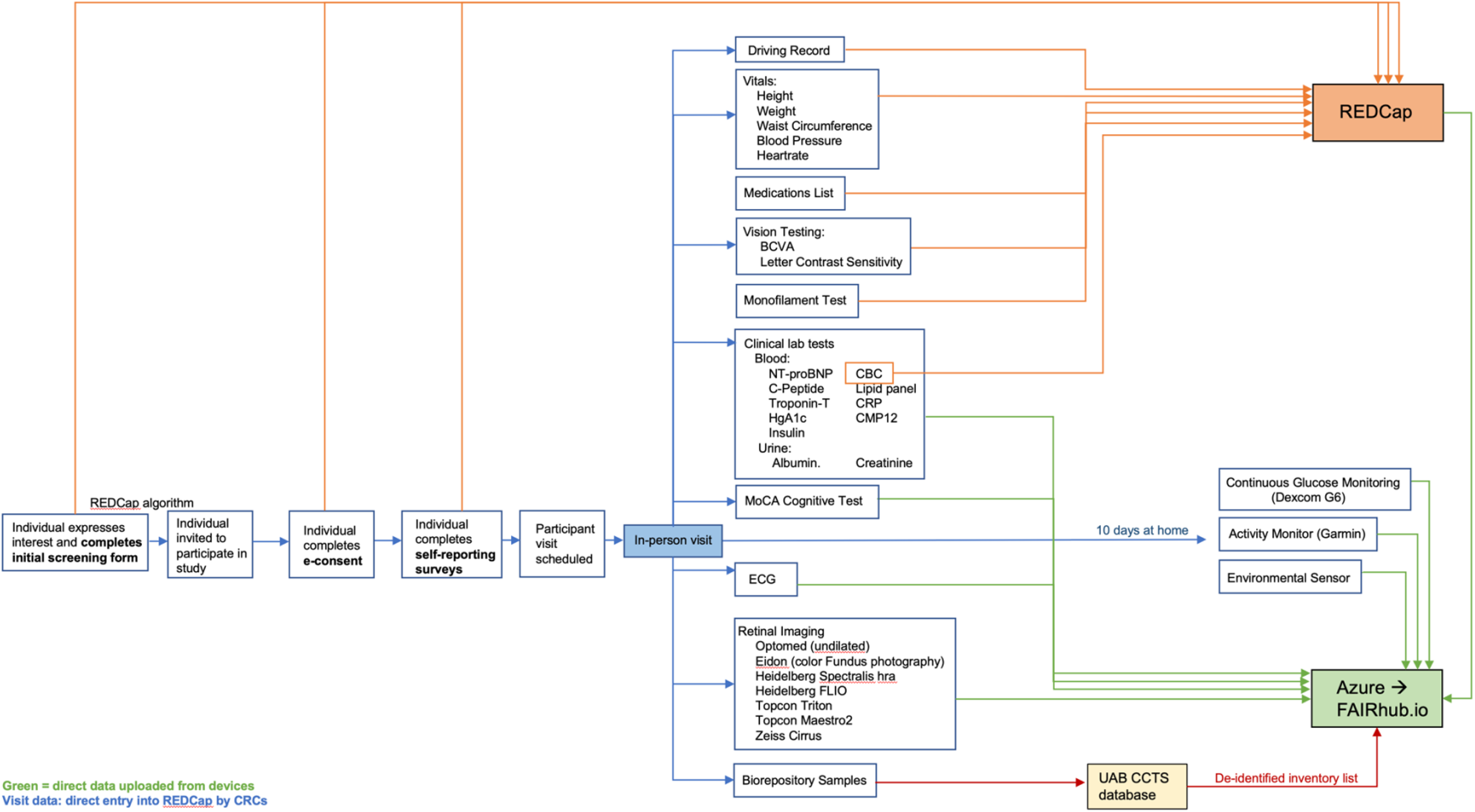
Data collected prior to and during the in-person visit will be either entered into REDCap or saved on local servers and batch loaded directly into Azure Storage Explorer. Azure Storage Explorer is a cloud-based storage platform used by AI-READI for collating, validating, and transforming data prior to its transfer to fairhub.io for dissemination to researchers. Fairhub.io is explained in more detail in section 8.2.

### 7.2 Data Entered Into REDCap

Data entered into REDCap comes from three sources: the EHR, the participant, or the CRC.

#### From the EHR

1. Name
2. Demographics (used for sampling, but not exported to fairhub.io)
3. Contact information

#### From the participant

1. Initial screening form
2. E-consent
3. Demographics, current contact information/preferred mode of contact
4. Health and functioning questionnaires completed electronically, **or**
5. Health and functioning questionnaires that are completed at the visit but into an iPad, or in hardcopy and entered into REDCap by CRCs.

#### From the CRC (entered through computer or iPad)

1. Driving habits, license information, and motor vehicle collision data
2. Physical assessment
3. Medications
4. Autorefraction
5. Visual acuity (photopic and mesopic)
6. Contrast sensitivity (photopic and mesopic)
7. Monofilament test
8. CBC lab results
9. Checklist of all data elements, including retinal imaging details and data transfer details.

Data exported from each site’s EHR will be collated, filtered, and then imported into REDCap to generate personalized access codes and urls to be used by potential participants for REDCap access. Hardcopy letters will be mailed to selected contacts and include a generic URL (https://redcap.ITHS.org/surveys/) and personalized survey access code that will allow participants to enter the AI-READI REDCap database and begin the screening process. Individuals who do not respond to the hardcopy letter (do not engage with our REDCap interface) within 7-14 days will receive an email from REDCap with a survey link that is unique to them. Individuals may click on the personalized link within the email and enter the REDCap database to read about the study and complete the screening questionnaire, if interested.

Data collected from self-reporting questionnaires will be entered directly into REDCap by participants prior to their in-person visit. Participants who complete e-consent in REDCap will be immediately presented with all questionnaires for self-completion. Individuals may use any device with internet access to complete these questionnaires. Individuals who do not complete questionnaires online will complete them during their in-person evaluation on iPads or in hardcopy. Clinical data will be entered directly into our REDCap database by CRCs using iPads or laptops. The iPads and laptops used for this purpose must have internet access. Hardcopy forms should always be available in the CRU in case of technology outages. If hard copy forms are used, the data must then be manually uploaded into REDCap by CRCs after the visit.

The AI-READI REDCap database will be housed on the UW server and CRCs will log into the database using their username and passwords. Transfer of REDCap data into Azure Storage Explorer (a cloud-based storage platform used for AI-READI data) will be performed on a schedule determined by the REDCap team, as described in section 8.3. Site Data Managers will verify that all participant records are complete in REDCap before indicating that the data is ready for data transfer to Azure. **Data Managers will use the REDCap form titled “Data Management” to indicate that an individual participant’s data is ready for transfer to Azure at two key stages:** 1) after the in-person visit is complete; and 2) after the 10-day home monitoring period is complete and devices are returned. All REDCap data transfer to Azure Storage Explorer will be handled by the REDCap team.

### 7.3 Data Uploaded Directly to Azure Data Explorer

Data that is not entered into REDCap is directly uploaded to Azure Data Explorer to allow for quality control and data transformation processes prior to final upload to fairhub.io and release to researchers. Site Data Managers are responsible for storing this data on local computers or servers prior to (and after) uploading to Azure. Each data site has their own storage container within Azure and site Data Managers are responsible for monitoring their Azure container to ensure that all non-REDCap participant data has been uploaded. Data that is uploaded directly to Azure from local storage is:

1. Continuous glucose monitor (Dexcom G6)
2. Activity monitor (Garmin watch)
3. Environmental sensor
4. ECG
5. Retinal imaging
6. MoCA Cognitive Screening
7. Clinical lab tests (except CBC results)

The procedure for data transfer from devices to Azure is addressed in section 8.4.7.

### 7.4 Inventory of Biorepository Samples at UAB

All collected biospecimens will be tracked by Participant ID and stored in a HIPAA compliant enterprise database hosted by the UAB Biorepository. No clinical or research data will be entered into the biorepository database. The only information stored will be specimen availability per Participant ID and the date the samples were collected. Reports of biospecimen availability will be periodically exported and made available on fairhub.io, along with instructions on how investigators may access stored specimens.

### 7.5 Missing Data

Participants will always have the choice of not answering a question on questionnaires. In order to enable the monitoring of form completeness in REDCap, each question in self-reporting questionnaires will include the option of selecting “prefer not to say” and/or “don’t know”, with the exception of validated surveys that have calculated summary scores. Individuals will need to choose the best answer for every question (or an opt-out answer) in order for REDCap to accept the questionnaire as complete. A single skipped question will render the questionnaire incomplete and the individual will not be able to submit and move on to the next questionnaire. Similarly, each data collection form in REDCap will have an option for the CRC to indicate when a participant has refused to answer or participate in an element of the protocol. CRCs are required to provide an explanation for why a protocol element was not performed. REDCap reports will indicate missing data on questionnaires and data collection forms. Missing data on self-reporting questionnaires may be addressed during the in-person evaluation and the answers added/forms edited by the CRC. Missing data on clinical forms must be explained by the CRC in REDCap. For every data collection form, CRCs must indicate if all data fields have been completed. If not, they must provide an explanation using the associated text field or in a comment linked to the data collection form. Site Data Managers will monitor REDCap reports for missing data and issue data queries to CRCs if necessary.

### 7.6 Data Editing

Clinical data that falls outside of expected min/max ranges will be flagged by REDCap, which will be visible in reports viewed by CRCs and Data Managers. Using these REDCap reports as guides, Data Managers and CRCs must then examine the participant record and determine if an error is likely. Data must be checked for the following and edited if errors are detected:

1. Credibility, based on range checks to determine if all responses fall within a prespecified reasonable range
2. Incorrect flow through prescribed skip patterns
3. Missing data that can be directly filled from other portions of an individual’s record
4. The omission and/or duplication of records

Editing should use available information and logical assumptions to derive substitute values for inconsistent values in a data file. This should only be done under the guidance and approval of the site PI and/or site Data Manager. If corrected data is available from elsewhere in the respondent’s answers, the error may be corrected. If there is no logical or appropriate way to correct the data, the site PI and/or site data manager should review the values and make decisions about whether those values should be removed from the database.

### 7.7 Retention of Study Documentation

Data uploaded into REDCap will be stored on the AI-READI REDCap database (UW server) for the duration of the study, even after data transfer to fairhub.io. All participant data stored on fairhub.io will be retained for a period that has yet to be defined. It is recommended that data from devices be stored on local computers or secure servers at the data collection site for the duration of the study.

### 7.8 Source Documentation

Copies of the informed consent document and all questionnaires, data collection forms, return of results documentation, home monitoring device instructions, and recruitment letters are located in the Appendix of this manual.

## 8. Data Management

This section outlines procedures for data extraction from devices and transfer of data to Azure Storage Explorer and fairhub.io.

### 8.1 Overview

Data protection will be facilitated by the Data Stewards at each institution. The central Information Technology (IT) group for each Data site’s institution will protect institutional data that is stored and maintained on servers at the site. The central IT group at UW, which will maintain the server that hosts the AI-READI REDCap database, will be responsible for ensuring safe data transfer from institutions to the REDCap database. Data Use and Data Share agreements between data sites have been established for the transfer of data (and PHI) between institutions. Azure Storage Explorer and fairhub.io are managed by the AI-READI Tools team (section 8.2). Data collected under the AI-READI protocol will be accessible via limited dataset access license (survey data, clinical measurements, retinal images, blood and urine tests, ECG, physical activity levels and heart rate, continuous blood glucose results, and environmental sensor data) or a controlled dataset access license (all items listed above, plus zip code, genomic sequencing, race/ethnicity, sex, medications, past health records, and motor vehicle accident reports).

### 8.2 FAIR Data ihub (fairhub.io)

The AI-READI Tools team has developed an open-source cloud-based data collection, curation, and sharing platform called fairhub.io.^2^ The platform was developed to support researchers in preparing and sharing their data according to the Findable, Accessible, Interoperable, Reusable (FAIR) data principles. Fairhub.io provides a convenient interface to upload data on-the-go, as it is being collected, from any web-browser. Researchers from the Data module will be walked step-by-step through our FAIR tools for making their data FAIR and AI-ready as per the guidelines that were established by the AI-READI Standards and Tools teams. Prior to the transfer of data to fairhub.io for dissemination, all AI-READI data will first be stored on Azure Storage Explorer to enable extensive quality control and data transformation processes. Validated data will be transferred to fairhub.io in discrete data set releases.

### 8.3 Transfer of REDCap data to Azure Storage Explorer

Transfer of REDCap data into raw storage containers within Azure will be automated using the REDCap API on a schedule to be determined. Site Data Managers will perform quality control checks on all participant data, verify that all participant records in REDCap are complete, and ensure that text fields are free of any personal information before indicating that the data is ready for transfer to Azure. **Data Managers will use the REDCap form titled “Data Management” to indicate that an individual participant’s data is ready for transfer**. All REDCap data transfer to Azure will be handled by the Tools team. **Note**: Data Managers should monitor the home monitoring device returns using available reports on REDCap (for example, “Device Return Information” and “Data Managers: Ready to release to Fairhub.io?“reports). Coordinators should contact participants who have not returned devices > 3 weeks after their in-person visit. If devices are not returned or lost in transport, this should be documented in the “Device Return” form using the questions asking about data export from monitoring devices. Once documented, the Data Management form may be marked as complete.

The automated data extracts are based on validated reports in REDCap that filter the data for different purposes - dashboards, OMOP mapping, interim/final REDCap datasets in CSV format, etc. All datasets will have a timestamp and will initially be stored in the raw storage container in Azure Storage Explorer. It will then be possible to limit reports in Azure by the participant’s study date, which will allow for discrete time snapshots for generation of the periodic data releases.

Periodic and final datasets will be validated by the Tools and Data teams prior to release for public dissemination. REDCap raw datasets will be moved to the final staging container in Azure Storage Explorer after the Data team has performed validation. A data dictionary will also be available for the REDCap datasets and included as a separate file in the final staging container in Azure Storage Explorer.

### 8.4 Transfer of Data from Devices to Azure Storage Explorer

Data from devices (Dexcom G6, Garmin watch, environmental sensor, retinal imaging devices, and ECG) will be directly uploaded from institutional storage to each site’s raw storage container in Azure at a frequency determined by the site. MoCA and clinical lab testing data will be uploaded directly to UAB and UW’s storage containers in Azure, respectively. It is recommended that CRCs or technicians initially upload data at a frequency of once per week so that any problems associated with data transfer are identified and remedied. Users will be given a URI to establish a secure connection to their site’s raw storage container within Azure. Data should be stored on local institutional servers prior to and after transfer to Azure. The sections below describe the process of downloading data from devices prior to upload to Azure. Device data will be moved to an interim staging container within Azure to enable quality control and data transformation processes to be performed. Once validated, device data will be moved to the final staging container in Azure prior to release to fairhub.io.

#### 8.4.1 Exporting Data from Imaging Devices

##### NOTE ABOUT EXPORTING IMAGES

Please export ALL images, including images that were retaken, and upload to the appropriate folder in Azure.

Export the data from each instrument and structure it into folders using the following site and instrument specific subfolders configuration. **Please note, capitalization is important and each folder has a unique filename**.

Folder structure (with UW as an example)

UW:

> “UW_Instrument”

>>“UW_Instrument_YYYYMMDD-YYYYMMDD” The dates will reflect the date range for the export (from date 1 to date 2).

>>> contents of these subfolders will be the data exported from the applicable instrument, without file renaming or reorganization (e.g. without creating subfolders per subject).

##### Optomed Aurora

- NOTE 1: AVENUE Sync is the proprietary software used for capturing and storing Optomed camera data. **DO NOT** manually delete image files from the computer–doing so will erase the patient identifiers on the camera itself the next time it is connected to the computer.
- NOTE 2: Storage on the Optomed Aurora should be periodically cleared (approximately every 1-2 months) to avoid syncing issues with the AVENUE Sync program. To avoid confusion, instructions for clearing the camera storage are included at the end of this section.

##### ONLY DO THESE STEPS THE FIRST TIME

– Open AVENUE Sync without the camera connected by clicking “Start by not connecting device”.
– Go to options and make sure that the field labeled “Local file storage” is exporting a to a folder called “C:\Users\{name of user directory}\Documents\imagesdicom”. You will need to create a new folder in “Documents” called “imagesdicom” for the images to export to.
– Also, make sure that “Show logs” is selected.
– Once you have set up the export folder, close the program and connect the camera to your computer.

##### DO THESE STEPS FOR EVERY EXPORT

– Make sure the camera is plugged into the computer and recognized as a storage device.
– Open AVENUE Sync.
– Select the camera from the camera list and make sure it is highlighted in blue.
– Click ‘Start by connecting device’.
– Go to the “Logs” tab to check that the dicom files are automatically exporting to the correct folder.
– You should see log entries similar to the image below for each dicom file that is exported.

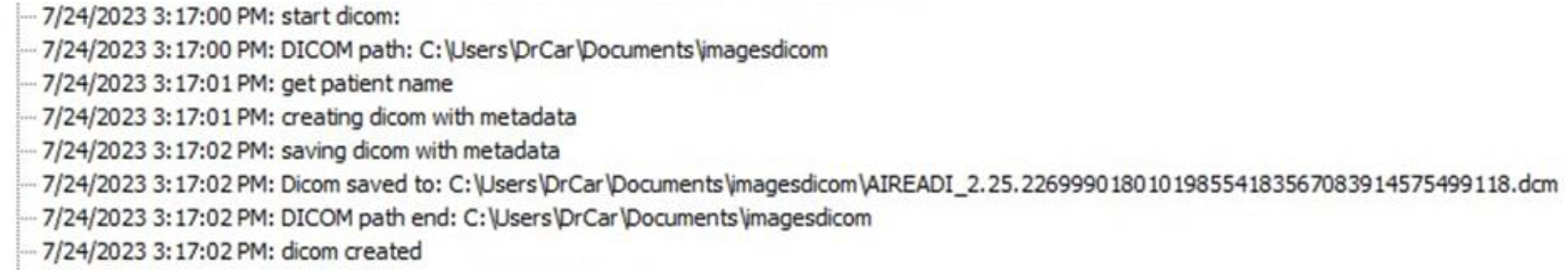
– Image files will look like this in your folder:

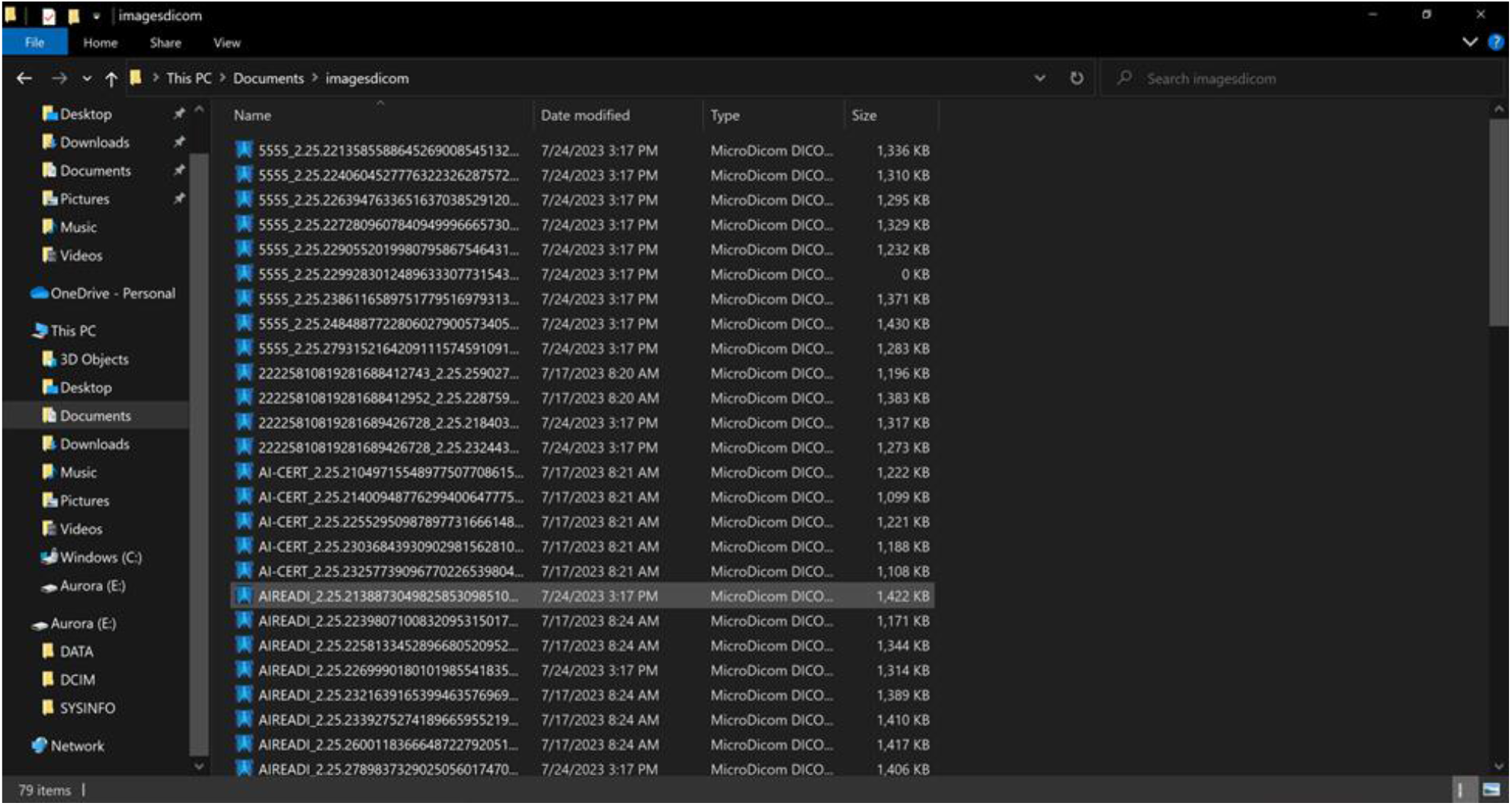
– Important:
  – Be sure you have used the “Four image sequence” option when creating a new subject to ensure the images are labeled correctly for which eye.
  – Images where the eye was not specified are labeled with “IM” at the beginning. This can be fixed manually on the camera.
  – If you need to delete any images, you will need to manually delete them on the camera before connecting to AVENUE Sync.

##### CLEARING CAMERA STORAGE

- Make sure that you have backed up all of the existing exports and images.
- On the Optomed Aurora, scroll over to settings (the little gear icon) and select “Camera.
- In the camera settings menu, pick “Erase image memory.”

Folder structure (with UW as an example)

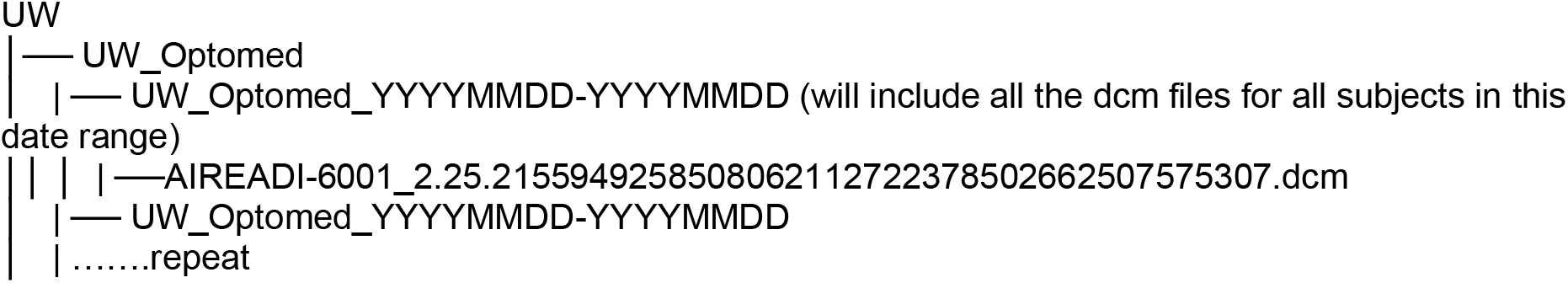

##### Topcon Maestro2

- IF YOUR DEVICE IS ONLY USED FOR AI-READI:
  - Go to the folder where the fda files are stored and access the ‘Data’ folder.
  - Copy the fda files, within the date range you need, to your USB or external harddrive.
- IF YOUR DEVICE IS USED FOR STUDIES OTHER THAN AI-READI:
  - Open IMAGEnet6 on the desktop
  - Under the ‘Basic’ tab, you can pick ‘All’ to view the list of patients and then scroll through to the date range you want or select the date range you want to only view the patients within a specified time period.
  - Find the ID in the list for the patient whose images you want to export. Double click on the patient ID to see the thumbnails of their images. You can also do so by manually clicking the Thumbnail button.
  - Hover or right-click on the thumbnail of the image to see the filename (e.g., ‘2006.fda’)
  - Go to the folder where the fda files are stored (at UW this is ‘Archive (E:)’) and go into the ‘Data’ folder.
  - Copy the fda files you need to your USB or external harddrive.

Folder structure (with UAB as an example)

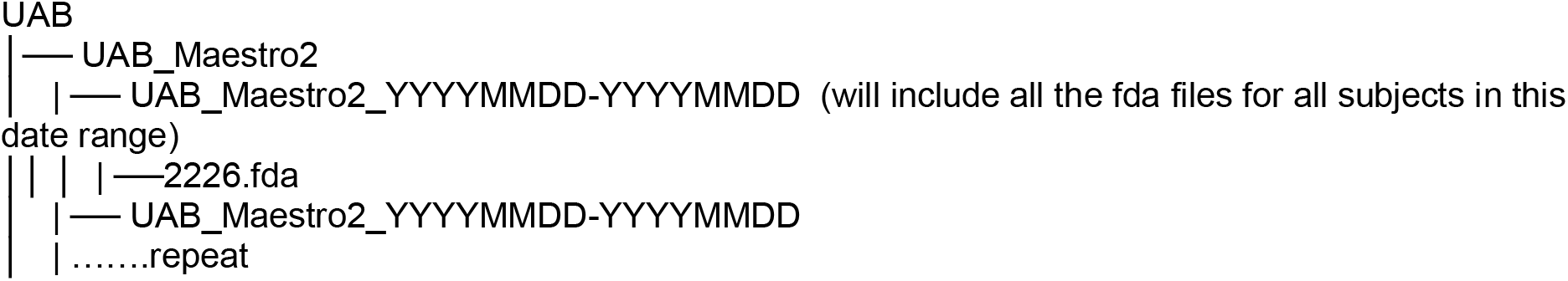

##### Heidelberg FLIO

- Open the file you have specified for storing FLIO data.
- Open the folder called ‘FLIO_DATA’.
- Select the files you need based on the options below.
  - If your device is only used for AI-READI, you can grab the files within your desired date range.
  - If your device is used for studies other than AI-READI, verify that the files you grab only have AI-READI ID’s.
- Copy the entire patient folder into your USB or hard drive. This patient folder should already be named in this fashion: [somenumber]_AIREADI_[participantID].
- **Upload the entire patient folder to fairhub**.

Folder structure (with UW as an example)

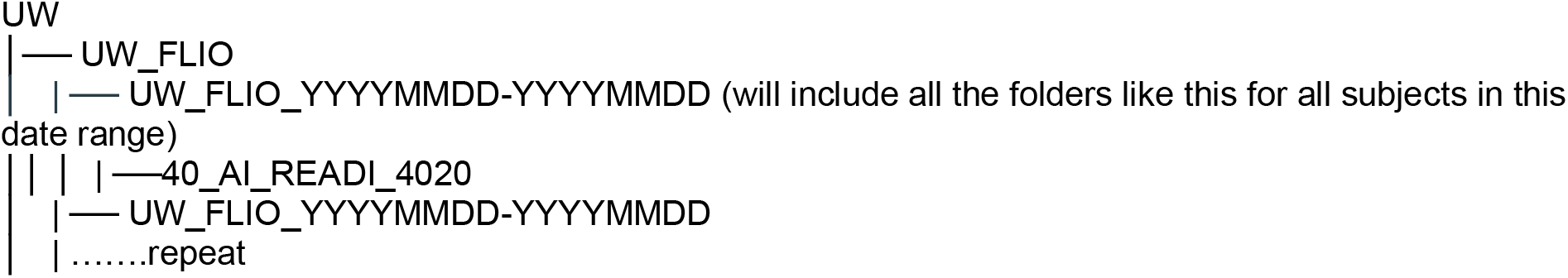

##### Topcon Triton

- IF YOUR DEVICE IS ONLY USED FOR AI-READI:
  - Go to the folder where the fda files are stored and access the ‘Data’ folder.
  - Copy the fda files, within the date range you need, to your USB or external harddrive.
- IF YOUR DEVICE IS USED FOR STUDIES OTHER THAN AI-READI:
  - Open IMAGEnet6 on the desktop
  Under the ‘Basic’ tab, you can pick ‘All’ to view the list of patients and then scroll through to the date range you want or select the date range you want to only view the patients within a specified time period.
  - Find the ID in the list for the patient whose images you want to export. Double click on the patient ID to see the thumbnails of their images. You can also do so by manually clicking the Thumbnail button.
  - Hover or right-click on the thumbnail of the image to see the filename (e.g., ‘2006.fda’)
  - Go to the folder where the fda files are stored access the ‘Data’ folder.
  - Copy the fda files you need to your USB or external harddrive.

Folder structure (with UCSD as an example)

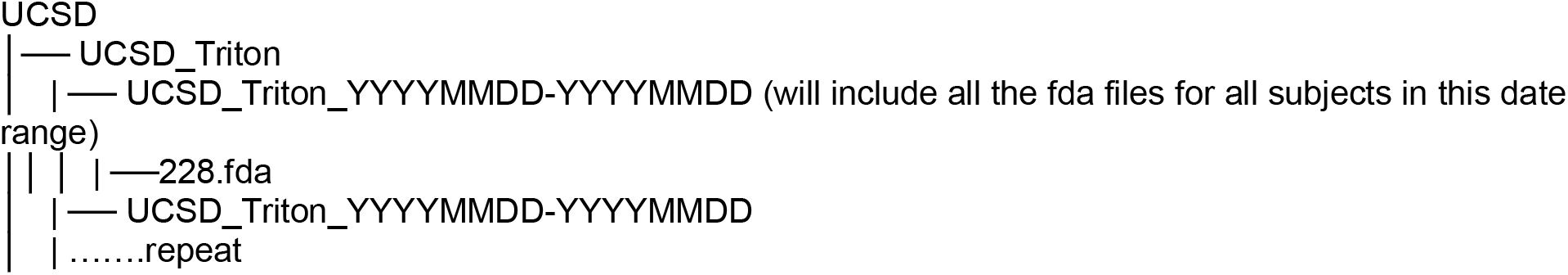

##### Heidelberg Spectralis

- Open HEYEX2 on the Desktop. Log in with your institutional credentials. You should be automatically directed to the “Review” tab.
- On the right side of the screen, find the Export Template. To export DICOM files, select the ‘DICOM’ tab in the lower part of the window.
- Look for your participants by filling out the study prefix in the “ID” field. Further narrow down your search by selecting the desired date range (past week, past 2 months, etc.)
- A list of patients should show up on the left side of the browser. Highlight the patients for whom you wish to export images from the list; hold down the left button on your mouse and drag the participants’ to the “Export” panel on the very right, and “drop” them on the logo that says “Export to Drive”. You can also click on the patient to see a list of images they have, and select all of the images you wish to export and drag and drop them on the “Export to Drive” logo.
- The export template symbol turns to blue.
- Drag-and-drop the export template symbol to the destination drive. Note that exporting directly to a portable drive may cause HEYEX 2 to crash; it is recommended that you choose one of the drives built into the computer.
- Browse to the desired folder and click OK to confirm.
- The export window is displayed. The export is completed if “**Finished successfully”** is displayed.
- Please note that this exports the images into multiple files that do not have file labels; the folder should be called DICOM. Copy the files onto the USB or hard drive you are using for exports.
- A DICOM viewer may be used to verify that all participant files were successfully exported.

Folder structure (with UW as an example)

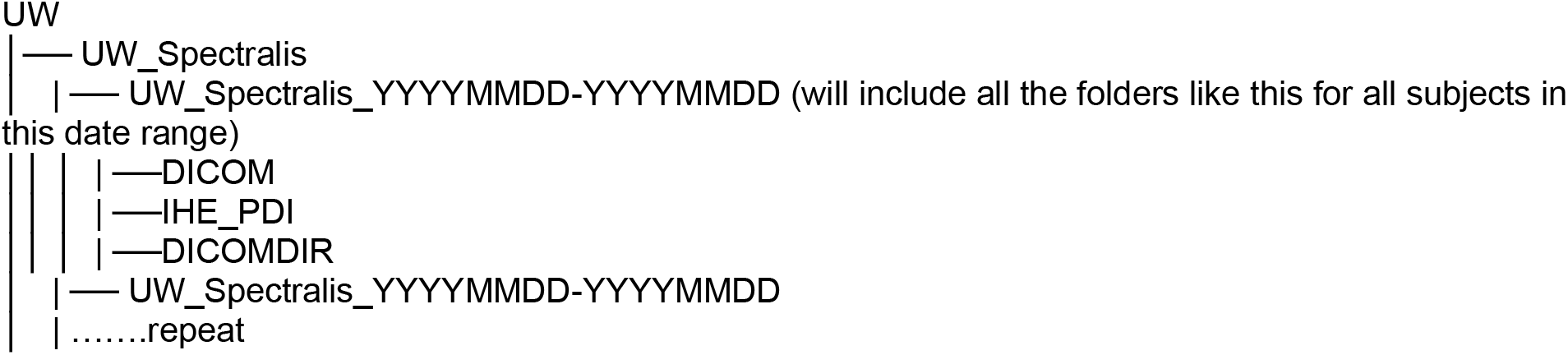

##### Eidon

**NOTE**: In the Admin account, go to the configurator and make sure you have selected to export DICOM images to the shared folder.

- On the tablet, log into either the Admin or Doctor account.
- Find your patient on the Eidon FA list (this list should be visible once you have logged into the program).
- Press the ‘Select’ button.
- Tap each image that you want to export; they should be highlighted in orange once they are selected.
- Tap on the paper airplane button and select ‘Export DICOM objects to shared folder’.
- In the shared folder, the exported files will be named starting with the last name then first name.
- Select the files you want to export and then copy them to your usb or hard drive.

Folder structure (with UW as an example)

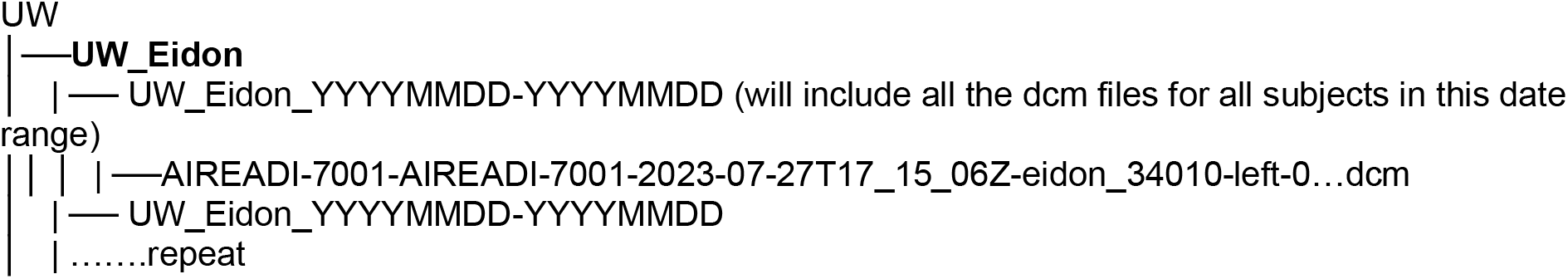

##### Zeiss Cirrus 5000

- Open Cirrus HD-OCT and log in
- Click ‘Records’ and select ‘Export Exams…’
- Click ‘Browse’ to select your export folder, which can be on the desktop or directly onto a USB or external harddrive.
- **Make sure ‘Export to Zip Format’ is NOT selected**
- Enter the last name, ID, and/or date range (by selecting Interval) into the appropriate boxes and
- click ‘Search’
- Select the participants you want from the list
- Click ‘Export’.
- Once the files have finished exporting, you should be able to find them in the location you picked for exporting.
- There should be a folder for each of the images.
- If you did not export the files directly onto a USB or external harddrive, then you can copy and paste the folders to a proper storage location.
- The data files will not be visible by participant ID. A DICOM viewer may be used to verify that all participant images are included in the batched folder.

Folder structure (with UW as an example)

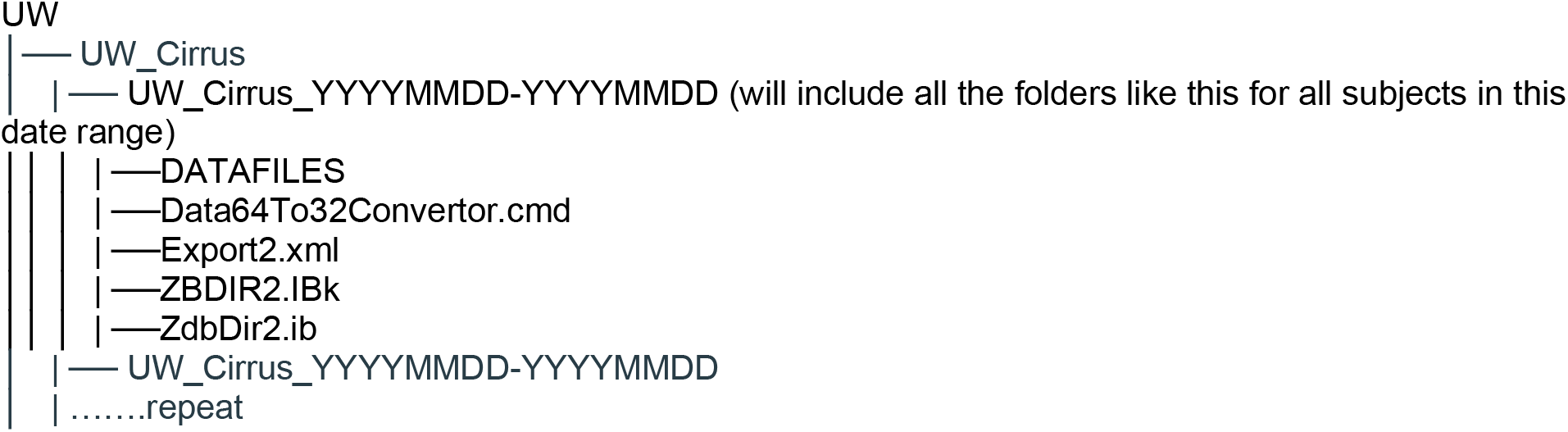

#### 8.4.2 Exporting Data from Dexcom G6 Transmitters / Dexcom Clarity CGM

Detailed instructions for the removal of the transmitter from the sensor is explained in Section 5.11.1. Once the transmitter is received from the participant, **Immediately download data from transmitter to reader**, following the instructions in Section “5.11.5 Receiving Returned Monitoring Devices and Extracting Data - Instructions for CRCs”. The transmitter has limited battery storage and data could be lost if data transfer is not performed quickly. Store data files on local harddrive or server for later upload to fairhub.io.

- Once the data is uploaded to Clarity.com, export the participant data as a .csv file and save on a local computer or server.
- **No renaming is necessary for the** .**csv file**. The data file will be uploaded to Azure in the format it is exported from Clarity. Do not change the file name.
- **Data will not be placed in date range subfolders**. Individual participant files will be uploaded to a CGM-specific folder in Azure as devices are returned.

#### 8.4.3 Exporting Data from Philips Pagewriter ECG Devices

Participant ECG files should be exported from the Philips Pagewriter at least weekly and saved on a local hard drive or secure server prior to uploading to Azure. **Importantly, previously saved ECGs must be periodically deleted from the device to provide space for new ECG files to be stored. You may not be warned that there is insufficient space for new file storage**.

##### Downloading and Saving ECG Data

- Insert a USB drive into the USB port on the right side of the ECG device
- Select “Archive” on the home screen
- Under “Transfer Destination”, make sure that “XML to USB memory” is selected
- Select the participant(s) you wish to download (scroll to right to see Participant ID and other headers)
- Select “Transfer” and wait until the transfer process is complete
- Remove USB drive and download files to a local computer or server for storage. **Each exported file will have a machine generated name ending in** .**xml (there is no need to rename files**; **we will upload files in the form they are exported from the device**). Note: ECG files will be uploaded to Azure in date-range folders, similar to those used for retinal imaging. See Section 8.4.7 for more details. Batch-exported ECG files must be placed in folders using the following nomenclature: “Site_ECG_startdate-enddate”, using YYYYMMDD format for dates representing the date ranges included.
- Upload to Azure according to instructions in Section 8.4.7

#### 8.4.4 Exporting Data from Environmental Sensors and Garmin Physical Activity Monitors Environmental Sensor

Follow the detailed instructions in Section “5.11.5 Receiving Returned Monitoring Devices and Extracting Data - Instructions for CRCs” for removal of the SD card and data retrieval.

##### Garmin Watch

Follow the detailed instructions in Section “5.11.5 Receiving Returned Monitoring Devices and Extracting Data - Instructions for CRCs” for retrieving Garmin watch data.

**Note:** Data from environmental sensors and Garmin watches will not be batched uploaded to Azure or placed in date range folders. Instead, individual participant files will be uploaded in their raw format to Azure as the devices are returned.

Environmental sensor and Garmin watch data should be stored on local computers/servers in folders named according to participant ID as described in Section 5.11.5. Data from these devices will be uploaded to Azure according to instructions in Section 8.4.7.

#### 8.4.5 Exporting Data from the MoCA Duo Application

The MoCA duo account is licensed to the AI-READI study under our IRB approval number. Thus, all data sites are approved under the same licensing account. MoCA test results are saved by the duo app for all 3 sites and can only be exported as cumulative reports containing all tests performed. For this reason, the Data Manager at UAB will be responsible for downloading all MoCA test results (for all data sites) and uploading the cumulative reports to Azure. Coordinators at each data site are only responsible for saving the test results for each participant on the app itself. For Azure, MoCA data will be batch exported and saved in a folder using the following naming rules:

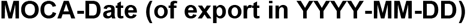

MoCA results will be uploaded to the MoCA folder within the UAB site-specific directory in Azure Storage Explorer.

**Important:** If the paper version of the MoCA was used, please contact the Data team Project Manager for further instructions.

#### 8.4.6 Clinical Lab Data from the UW NORC lab

Clinical lab data will be returned to UW after analysis by the UW NORC lab, using the manifest documents originally used to inventory/ship the samples. Files must be saved in .csv format and renamed as:

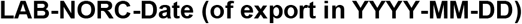

Lab data results will be uploaded to the Lab_NORC folder within the UW site-specific directory in Azure Storage Explorer.

#### 8.4.7 Uploading Data to Azure Storage Explorer

Each Data site will have their own designated site-specific directory within the raw storage container on Microsoft Azure Storage Explorer. Data managers at each site are sent a secure URI for their site and following the provided instructions, will have access to the Azure raw storage staging area containing an upload folder for their site only.

Data containing subfolders for each device will be generated as described in the export sections above. Within the raw storage area of Azure, the following folders are available within each site-specific directory for data uploads:

Example for UW:

**UW_Cirrus**

**UW_Optomed**

**UW_Triton**

**UW_Spectralis**

**UW_Eidon**

**UW_FLIO**

**UW_Maestro2**

**UW_EnvSensor**

**UW_CGM**

**UW_FitnessTracker**

**UW_ECG**

**MOCA** (UAB only)

**Lab_NORC** (UW only)

Data from devices can be uploaded into the appropriate device subfolders on a biweekly, weekly, or monthly basis. The frequency of exporting and uploading can vary by site. There are advantages to exporting and uploading on a monthly basis as the date ranges are clear and there is less chance for error. However, given the high monthly recruitment targets, monthly exports and uploads may be very time consuming, requiring instrument downtime to the exports and monitoring of the completeness of the uploads, and therefore the frequency of site exports and uploads can vary by site and instrument. Refer to sections covering each device for specific instructions on file structure/upload procedures. **While retinal imaging data may be uploaded in date-range subfolders (“Site_Device_startdate-enddate” (date format is YYYYMMDD), home monitoring devices (environmental sensor, CGM, fitness tracker) will be uploaded individually to appropriately named top level folders in Azure as they are returned and will NOT be uploaded in date-range subfolders. ECG data will be uploaded to Azure in date-range folders, using the same format as retinal imaging folders (“Site_ECG_startdate-enddate”). MoCA and NORC lab results will be uploaded in batches in date-range folders**.

Prior to uploading, Data Managers should perform quality control on all data files to be sure that they conform to proper protocol conventions and are in the correct format.

### 8.5 Quality Control Procedures

#### REDCap data

All data entered into REDCap will be monitored closely by the CRCs and Data Manager at each site. Any missing items in questionnaires, data collection forms, or other electronic documents will be flagged within REDCap and then resolved by the CRC either during the in-person visit or after (as described in Section 5). If missing data cannot be resolved, then an explanation in REDCap must be provided. Data entered directly into REDCap from iPads will not have hardcopy backups, which precludes the ability to have 2x data entry quality control. However, REDCap entries should be double-checked and any values that lie outside of expected ranges should be identified and notated in REDCap as potential input errors. Data Managers at each site should use the multiple QC reports available to them in REDCap to validate all data.

#### Quality control of wearables, environmental sensor, ECG device, and Clinical Lab Tests

CGM: CRCs will verify that the G6 device is inserted properly into the skin, is initiated properly, and the transmitter is properly paired to the sensor prior to leaving the CRU. After the transmitter is received back from the participant, the CRC will verify that data was collected for 10 days of glucose monitoring by downloading the data from the receiver and looking for gaps in data collection. Any missing data should be indicated in REDCap.

Garmin watch/activity monitor: CRCs will verify that the Garmin watches are positioned correctly on the participant’s body and that the instructions for use are understood. When the watch is returned, the data will be downloaded by the CRC and transferred to Azure. If CRCs find a low number of files on the returned watch, it must be notated in REDCap and followed up with testing of the watch for functionality. Comments about missing data should be provided using both the “comments” function in REDCap (link the comment to the “Return” form) and in the disposition form (deviations from protocol; but not an adverse event). Verification of proper date and time, as well as proper working order must be performed on each watch **prior** to being given to a participant. This is especially important because Garmin watches will be reused.

Environmental Sensor: There will be no real-time analysis of data quality during the monitoring period. CRCs will verify that the sensor is properly operating prior to sending it home with a participant and that data download is working normally. If the CRCs are notified by the participant that the sensor is not working, they may arrange for a replacement and notate the problems in REDCap. Each environmental sensor should add approximately 18-20 files per day during the monitoring period. Sensors returned with fewer than expected files should be notated in REDCap using both the “comments” function in REDCap (link the comment to the “Return” form) and in the disposition form (deviations from protocol; but not an adverse event). The sensor should be tested for functionality before returning it to circulation. Verification of proper working order must be performed on each device **before** it is given to a participant. This is especially important because the environmental sensors will be reused.

ECG: CRCs will verify that the Philips ECG device and data download is working properly prior to use on a participant. The proper functioning of the ECG device should be verified on each participant following the manufacturers guidelines. If there are any issues with data collection or data download from the device, it must be adequately described, in detail, in REDCap. ECG data will be uploaded as raw .xml files to Azure. Prior to data upload to Azure, PDF files of participant ECG traces should be viewed for quality, looking for wandering baseline, missing leads, or low precordial voltage. It is important to provide feedback to CRCs when the aforementioned quality issues are observed. The best representative ECG file for each patient should be chosen for upload to Azure.

Clinical lab results: Upon receipt of lab results from the NORC lab, data will be examined for extreme out-of-range values and any obvious batch effects (e.g. values in undetectable ranges for a continuous series of participant IDs). If either of the aforementioned issues are detected, re-testing may be requested for validation.

Quality control of Azure uploads: During the data validation period that precedes a new data release, Data Managers at each site will be given a list of participant IDs to be included in the new data release. Data Managers must verify that the data for each participant is present in the appropriate folders of Azure and that all folders are named using the correct format. Only after validation will data be transferred to the final staging area. Secondary, independent data validation checks will be performed by members of the Tools and Standards teams prior to release to fairhub.io.

## 9. Reports

### 9.1 Potential Adverse Events

Adverse events may occur during the course of data collection. For this specific protocol, examples of adverse events may include the following. We are anticipating these adverse events to be rare.

#### Mild

Dilation lasting more than several hours and/or excessive irritation and tearing from the dilating drops: This is more than minor discomfort that dissipates very rapidly in the vast majority of persons. This is not an event that can be anticipated for a given participant.

Skin irritation from the continuous glucose monitor

Skin irritation from the Garmin watch.

Dry eye symptoms, blurry vision, and headaches are possible adverse events associated with this protocol.

Bruising and pain at the site of blood draw.

Fainting after blood draw is possible, especially in patients with T2DM. To help mitigate this issue, we provide small snacks following the blood collection. We also offer the option of reclining during the blood draw to minimize this risk

#### More serious

Participants could fall or be involved in a motor vehicle accident due to blurry vision after dilation. Although these potential events are exceedingly rare, we will warn participants about walking carefully and encourage them to be picked up from their visit in order to help prevent them from occurring. We will provide disposable sunglasses to all participants to reduce the effects of light sensitivity.

Other rare events that have been associated with pupillary dilation are angle closure attacks, allergic reactions, and systemic reactions such as increased blood pressure, tachycardia, arrhythmia, and dizziness. If any of these events occur, appropriate medical intervention will be immediately sought. Also extremely rare, the venipuncture site may become infected, which could lead to hospitalization or death.

Serious adverse events (SAE) will be reported immediately to the UW IRB, the IRB of record for this multi-site study. SAEs would be hospitalization, disability, or death of a participant, and must be reported within 24 hours of the team learning of it. Because this is a cross-sectional study, as soon as the subjects complete the visit and the 10-day monitoring period, they are considered off the study. To report SAE’s, UW has an online form which falls under “promptly reportable information” submission. On that form there is a list of items to check. Depending on what is checked, additional forms may be activated and need to be completed by the study team. For information on reportable events at UW, go to this site: https://www.washington.edu/research/hsd/study-activities/report-events-and-new-information/guide-to-reporting-new-information/

Although all SAEs must be reported to the central IRB at UW, there may be additional reporting conditions that must be met at UAB and UCSD, based on institutional guidelines.

### 9.2 Incidental Findings

It is possible that technicians and CRCs will observe potential abnormalities in the participant data they collect. Examples could include possible abnormal heart rhythms in ECG traces and retinal images that are consistent with retinal disease. Participation in the in-person visit does not constitute a proper exam under a physician’s care and there will be no consultation with physicians on findings (in most cases we would not have a primary care physician to contact). Technicians/CRCs are not qualified to make medical diagnoses. Thus, we will not be following up on incidental findings with the participant or health care provider. However, important exceptions to this policy exist (see below).

#### Incidental findings during the Physical Assessment

Blood pressure and heart rate data collected during the in-person visit will be provided to the participant on a data return card (See Appendix F). If a participant presents with blood pressure and/or heart rate values that are outside of published normal ranges, coordinators will follow the procedures listed below:

##### Systolic Blood Pressure

- Readings of >180 mmHg or <100mmHg (with any symptoms of hemodynamic instability): patient is immediately advised of the need for emergent care, stop visit and recommend patient go to the emergency department.
- Readings of 130-180 mmHg: recommend that patient seek medical care and notify their provider.

##### Diastolic Blood Pressure

- Readings of >120 mmHg or <60 mmHg (with any symptoms of hemodynamic instability): patient is immediately advised of the need for emergent care, stop visit and recommend patient go to the emergency department.
- Readings of 110-120 mmHg: recommend that patient seek medical care and notify their provider.

##### Heart Rate

- <60 or >100 bpm, not known to be usual for the participant (with any symptoms of hemodynamic instability): patient is immediately advised of the need for emergent care, stop visit and recommend patient go to the emergency department.
- <60 or >100 bpm and hypotension (systolic blood pressure of <90) without symptoms of hemodynamic instability: patient is immediately advised of the need for emergent care, stop visit and recommend patient go to the emergency department.
- Asymptomatic heart rate <50 bpm or >120 bpm, not known to be usual: recommend that patient seek medical care and notify their provider.
- Heart rate >100 bmp and irregular pulse unable to be determined via digital cuff: recommend that patient seek medical care and notify their provider (unless confirmed by participant that an irregular heart rhythm is already a known condition)

#### Incidental findings during retinal imaging

Retinal imaging technicians may notice potentially serious eye conditions during the imaging procedure. Because we do not have an ophthalmologist evaluating our participant images and technicians do not have the expertise or credentials to diagnose ophthalmic conditions, we plan to intervene only when life-or vision-threatening conditions are implicated. This includes retinal detachment, tumor, and optic disc edema. Imaging technicians will be trained to detect these conditions at the time of image acquisition and if detected, follow the site-specific procedures for notifying the participant. **Participants with suspected disc edema will be referred to the Emergency Department for immediate care. Participants with a suspected retinal detachment or tumor will be advised to seek immediate care with their ophthalmologist and/or provided a referral to an ophthalmologist**. Our approach to handling these conditions is based on balancing the need for immediate care with the understanding that participants may have insurance that does not cover services at our institutional hospitals/clinics (or they may prefer to use providers outside of our healthcare system). Each data site has specific procedures to be followed for eye-related incidental findings.

### 9.3 IRB

The initial IRB approval at the University of Washington was received on December 20, 2022. Under FWA #00006878, the IRB approved activity for the AI-READI study from 12/16/2022 to 12/15/2023 and has extended approval through 10/23/2026. The UW IRB is the central IRB on record with UAB and UCSD ceding review through reliance agreements. The AI-READI study is required to (1) obtain IRB approval before making any changes (modifications) to the study, and (2) provide the IRB with any reportable new information such as breaches of confidentiality or unanticipated problems.

IRB approval is required for UW and a separate approval notice is provided to each individual participating site. Approval is for the number of subjects described in the application (in total and for each group). A modification to request an increase in the number of approved subjects, if necessary, is required. Exceeding the IRB-approved number (over-enrollment) will be considered non-compliance. Before enrolling non-English speaking subjects, the IRB must receive all translated consent materials that will be provided to subjects in written or electronic form.

The IRB made a determination that the following waivers are granted for this study:

- Waiver of consent for pre-screening subjects to determine eligibility
- Waiver of HIPAA authorization for pre screening to determine eligibility.

*Note that any granted waivers of consent do not override a subject’s refusal to provide broad consent*.

Tracking IRB approval periods and preventing a lapse is ultimately the UW research teams’ responsibility. However, the UW IRB system, “Zipline”, sends courtesy reminders prior to expiration of approval. If a renewal application or study closure is not received within 90 days of expiration, the Human Subjects Division (HSD) may administratively close the study. In some circumstances, HSD may refuse to review additional submissions from the researcher until a status report is received, the lapse may be considered continuing non-compliance, and the study may be “terminated” by the IRB.

### 9.4 NIH Reporting

#### 9.4.1. Quarterly Progress Reports

Year 1, Quarter 1 (Y1Q1) interim progress report – 3 months after the start of the Award

Year 1, Quarter 2 (Y1Q2) interim progress report – 6 months after the start of the Award

Year 1, Quarter 3 (Y1Q3) interim progress report – 9 months after the start of the Award

Year 1, Quarter 4 (Y1Q4) interim progress report – Annual Report is due instead of Quarter 4 interim progress report

##### Annual Progress

Annual progress report – due at the end of the current project year as indicated in the bilateral agreement. The Annual Report is cumulative, i.e. covers the activity in Q1-4. The annual report also counts as the progress report for the Quarter 4 of the project year.

##### Financial Reporting

SF425: The SF-425 can be accessed at https://www.grants.gov/forms/post-award-reportingforms.html. The FFR must be uploaded via eRA Commons. Instructions for submitting the FFR can be accessed at: https://era.nih.gov/erahelp/commons/FFR/Archive_FFR/ffr_submittingForm.htm

In addition to formal reporting listed above, we have monthly Steering Committee meetings and bi-monthy small group discussions with NIH program officers.

#### 9.4.2 Milestone Tracking and Reporting

AI-READI milestones are available in a project Gantt chart and accessible to all project members in the shared AI-READI Google Drive. Milestones will be monitored by each milestone team and project managers will update the Gantt chart every quarter. Milestones associated with data collection will be monitored and reported by UAB. Milestone completion and challenges will be reported to the NIH in quarterly progress reports and the Annual Report.

## 10. Data Release and Public Access

Harmonized data will be released for licensed public access once per year, according to a schedule that will be determined after the pilot year. Each data release will be additive, containing both new and previously released data. Access to the AI-READI dataset is described at https://www.aireadi.org.

## 11. Study Completion and Closeout Procedures

### 11.1 Participant Notification and Return of Data

Due to delay in obtaining the laboratory results and lack of standard interpretation methods for other data (e.g., imaging), it is unlikely that most of the data collected during this protocol would be clinically actionable. All blood and urine testing results will eventually be available to the participants, but there may be a substantial lag between the study visit and the return of results.

Subjects will receive some results obtained on site during the visit (e.g., blood pressure and vision exam results) or after the return of monitoring devices (e.g., CGM results). However, laboratory testing results may not be available within the clinically actionable timeline. The laboratory test results will be emailed to the participant after they are available, usually within 3-4 months. The lab results will be provided in a document that explains what each test is measuring and its purpose, along with a graphic showing their individual result for the test and where it falls in the reference interval for “normal” (See Appendix F). The document instructs the subject to discuss the results with their primary care provider or specialist. AI-READI will not provide interpretation of the data due to the high number of subjects and the absence of a physician overseeing the results. A summary of the results returned to participants is provided in Table 11.

**Table 11.** List of results that will be available to subjects.

| Domain | Variable |
| --- | --- |
| Vitals | Systolic / Diastolic blood pressure |
| Vitals | Heart Rate |
| Eye Health | Visual acuity of each eye |
| Systemic Health | Complete Blood Count (white blood cell count, red blood cell count, platelet count, hemoglobin) |
| Systemic Health | Comprehensive Metabolic Panel (Glucose, Calcium, Total Protein, Globulin, Albumin, A/G Ratio, Chloride, Carbon Dioxide, Potassium, Sodium, ALT, AST, Bilirubin, Alkaline Phosphatase, BUN (Blood Urea Nitrogen), Creatinine, BUN/Creatinine Ratio, estimated Glomerular Filtration Rate) |
| Systemic Health | Hemoglobin A1c |
| Systemic Health | Lipid Panel (Total Cholesterol, Triglycerides, HDL Cholesterol, VLDL Cholesterol) |
| Urine Analysis | Urine Albumin |
| Urine Analysis | Urine Creatinine |
| Cardiology | High sensitivity troponin (hs-TNI) |
| Cardiology | BNP |
| Endocrinology | Hs-CRP |
| Endocrinology | C-peptide |
| Endocrinology | Continuous Blood Glucose (from Dexcom G6) |

Since we are recruiting from the EMRs, not every patient is seen by a specialist at the study sites or has access to care from a specialty clinic. Thus, we will return the data that can be interpreted by a primary care doctor. In addition, we are not distributing any test results that do not have standard interpretation methods because of their raw data format (e.g., retinal images, ECGs, fitness tracking and environmental sensor data). Genetic testing data, cognitive testing results, results derived from samples left in a biorepository or data obtained in future research projects will not be returned to participants.

The study team will not curate the results in any way and therefore cannot exclude results that are normal and do not require treatment or intervention. Knowing that their values are normal may be reassuring to the subjects. Furthermore, these normal findings may serve as a baseline in case of any future abnormal findings.

### 11.2 Participant Exit Survey

When team members complete the “Device Return” form in REDCap indicating that a participant has returned the home monitoring devices, they must also select the option to trigger REDCap to email the participant a voluntary exit survey (Appendix G). The survey is intended to better understand the participant experience and provide data to the Data team for assessing participant satisfaction. Data from the Participant Exit Survey may be used to improve the study.

## 12. Policies

### 12.1 Confidentiality Procedures

The AI-READI database will only publicly release de-identified data, randomized participant IDs, and biospecimen availability and storage location (by participant ID). Any protected health information accessed and used during the recruitment process will be held in a secure REDCap database. Controlled access to the Azure limited dataset that includes genetic information will only be shared with appropriate security and approval measures in place. For more information on how AI-READI data is being stored and protected, see the following publication on behalf of the AI-READI consortium: https://doi.org/10.1016/j.eclinm.2025.103729

Data will be shared with the investigators in this project who are part of developing software for de-identifying, data uploading and data sharing. Principal investigators and lead co-investigators at each participating institution will oversee and/or manage the sharing. All of the research staff under the PI and lead site PIs will be able to see data under the supervision of the PIs and site PIs. De-identified data will be released for download following an application process as described at https://www.aireadi.org. The risk of a confidentiality breach is low because all identifiable information will be kept on encrypted computers/servers or in a locked cabinet/room with restricted access at all sites. Only authorized research personnel will have access to identifiable information.

### 12.2 Scientific Publications and Presentations

Any abstract or manuscript that arises from the AI-READI project will be approved by the Steering Committee before it is submitted. The order of named authors on the author line are at the discretion of main authors of the publication, and the author line must end with “on behalf of the AI-READI Consortium”. The list of AI-READI consortium members may be provided as a supplement to the manuscript. Any individual within the project may be included in the AI-READI consortium list, provided they contributed to the data contained within the manuscript or contributed to the writing and/or review of the manuscript. For every manuscript submitted, team members who wish to be included in the AI-READI consortium list must approve their inclusion and indicate that they have read and approve of the manuscript.

Future manuscript and presentation preparations, grant preparations, future research studies, future AI/ML algorithms, repository/data sharing platform creations, and future research using the stored specimen and data are anticipated after data release for restricted public access, and proper acknowledgement of NIH Bridge2AI funding is required.

### 12.3 Publicity

There is a public-facing website for the AI-READI project that will provide information about the Bridge2AI program and AI-READI. The website may be found at: https://aireadi.org/

The AI-READI Advisory Council (AAC) consists of members of the community at each Data site location. The AAC will be used to gather feedback about elements of the project, as well as serving as an interface between the science and the larger community.

## 13. Human Subjects Protection

All research activities will be compliant with current federal and institutional policies of human subjects research. All research team members will have current training in human subjects research.

There are minimal risks associated with completing surveys and questionnaires regarding their general health and experience with T2DM. Participants may become fatigued or bored during survey completion. They may feel uncomfortable answering some sensitive questions. Questionnaires will be self-reporting and completed prior to the in-person visit, allowing for participants to answer those questions in a private and safe environment of their choosing. For self-reporting questionnaires that were not completed by participants prior to their CRU visit, we will provide a private space for their completion. Participants will be given options to opt out of questions they do not feel comfortable answering. Participants may experience eye fatigue during visual acuity tests, letter contrast sensitivity tests, or retinal imaging. Breaks will be provided to ensure that participants are comfortable. Pupil dilation will cause light sensitivity and some participants may find this uncomfortable. Disposable sunglasses will be provided to participants to ameliorate this sensation. In some instances, pupil dilation may cause a stinging sensation due to the effect of the drug on the dilating muscles within the eye. This sensation is temporary. Blood collection may be uncomfortable for some. To minimize this possibility, highly trained and experienced phlebotomists will be used for collection. Participants will be reminded to arrive at their appointment well-hydrated and rested and will be offered a snack after their blood draw. Urine collection is a routine practice and should not cause discomfort. Participants will be informed that they may discontinue participation at any time. Snacks will be provided to all participants.

There is a risk of loss of confidentiality as well as the possibility that personal information may inadvertently be revealed. Participants will be assigned a unique study number in order to de-identify information collected for analysis. All written surveys and data collection report forms will be coded with a unique anonymous identifier for tracking purposes and data will be entered into REDCap using this unique identifier. Data will be stored on password-protected servers and all written materials will be stored in locked spaces only accessible by approved research staff. All computer files will be secured in password-protected files only accessible by approved research staff. The final protocol will be accompanied by a manual of procedures. Any deviations from the protocol or procedures will be documented and shared with the IRB.

The consent to participate in AI-READI will include a request for permission for future studies of residual biospecimen material, additional analysis of existing biospecimen material, and sharing of research data and materials with ancillary studies and other investigators.

Because this is a data collection study, participants will not likely recognize personal health benefits from participating. Participants will have access to results as listed in Table 11, but it is not likely that they are given this information in a timely matter. They may benefit from knowing that they are contributing to a groundbreaking dataset that will provide many opportunities for scientists to better understand the progression of T2DM, as well as the return to health (salutogenesis).

## 14. MOP Maintenance

The AI-READI MOP will be maintained by UAB Data members. Any changes to the procedures described within should be discussed with all Data MPIs to maintain standardization across sites. Exceptions are the order of performing some procedures during the in-person visit, which can be site-specific. All amendments to the MOP will be indicated with a date and short description on a page preceding the Table of Contents. This document is the final version of the MOP with all amendments incorporated.

## 15. List of Supplies and Equipment

Table 12 lists the supplies and equipment that all Data sites should have on hand in order to implement the AI-READI protocol.

**Table 12.** Supplies and Equipment.

| Element of Protocol | Purchasing Information |
| --- | --- |
| **General** |  |
| Sani-Cloths or equivalent | UAB: Super Sani-Cloth Germicidal Disposable wipe |
| Face Masks |  |
| Gloves |  |
| Disinfectant Spray |  |
| Hand Sanitizer |  |
| Pens |  |
| iPads (at least 2 per site) with the following installed:<br>• REDCap mobile<br>• MoCA Duo App | The MOP provides instructions on how to download these applications |
| Stylus or iPad pencil (generic) | Not standardized; any will do |
| Bluetooth keyboard for iPad (optional) |  |
| Hardcopy forms - all (as backup) | Will need to print REDCap forms for all questionnaires and data collection to keep on hand as backups |
| Gift cards for honorarium | Will be site-specific |
| Wide-mouth sharps container | For disposal of G6 applicator |
| Biohazard disposal bin |  |
| **Physical Assessment** |  |
| Scale (mechanical beam or digital) | Recently calibrated |
| Height Rod | Must provide metric values |
| Tape measure | Must provide metric values |
| Digital sphygmomanometer (must also give HR) | Any manufacturer is fine as long as it is clinical quality |
| **Vision Testing** |  |
| Occluder |  |
| ND-occluder Neutral Density filter) for Mesopic tests | https://www.edmundoptics.com/p/20-od-25mm-dia-absorptive-nd-filter/8293/ super glue the lens to a occluder |
| M&S Technologies EVA device | CTS-EVA-e-ETDRS |
| Mars Letter Contrast Sensitivity | https://marsperceptrix.com/ |
| autorefractor |  |
| 20 inch string or tape measure |  |
| stand for Mars charts | UAB uses a KopyAid metal stand |
| Light meter (lux meter) | For setting appropriate ambient light levels (ex: Sper Scientific Lightmeter) |
| **Retinal Imaging** |  |
| Optomed Aurora | Supplied by Optomed |
| Dilating drops |  |
| Eidon Fundus (iCare) |  |
| Spectralis Hra OCTA (Heidelberg Engineering) |  |
| Triton DRI OCT (Topcon Healthcare) |  |
| Maestro2 3D OCT-1 (Topcon Healthcare) |  |
| Angioplex - Cirrus 5000 (Zeiss) |  |
| FLIO (Heidelberg Engineering) |  |
| Tissues |  |
| Disposable sunglasses |  |
| **Electrocardiogram** |  |
| Philips Pagewriter TC30 Cardiograph |  |
| Reclining chair |  |
| Hospital gowns or disposable exam capes |  |
| Disposable electrodes | CardinalHealth Kendall 5400 Diagnostic Tab Electrodes; Cardinal Health part # 31433538. https://www.cardinalhealth.com/en/product-solutions/medical/patient-monitoring/electrocardiography/diagnostic-ecg-electrodes/kendall-diagnostic-tab-electrodes.html |
| Alcohol swabs or wipes |  |
| Gauze pads |  |
| Adhesive removal wipes |  |
| Disposable drape sheet |  |
| Razor |  |
| **Monofilament Test** |  |
| Monofilament testing | Disposable 10g monofilament; AliMed disposable monofilaments 10g. (5.07) ADA. 20/pk, ALI32745, Medline.com. https://www.medline.com/search/products/ALI32745<br>Reusable monofilament https://www.alimed.com/retractable-monofilaments.html?srltid=AfmBOopCZGHmc0tlqlaatLkzSzpKgw6Ak03Nn60dYE1dXvB-K37ob8dw |
| **Clinical Lab Tests and Biorepository Samples** |  |
| centrifuge(s) - varies by site | Swinging bucket rotors compatible with the vacutainers being collected |
| Lavender-top EDTA Vacutainers | 6ml draw (13x100mm) BD 367863 - Need 2 per participant<br>3ml draw (13x75mm) BD 367856 - Need 1 per participant<br>**\*tubes have expiration dates; do not order too many at a time** |
| Gold top Gel-barrier Vacutainers (SST/serum tubes) | 5ml draw (13x100mm) BD 367986 - Need 2 per participant<br>**\*tubes have expiration dates; do not order too many at a time** |
| PAXgene RNA Vacutainers | 2.5ml draw (16x100mm) BD 762165 - Need 1 per participant<br>**\*tubes have expiration dates; do not order too many at a time** |
| CPT Vacutainers | 8ml draw (16x125mm) BD 362761 - Need 2 per participant<br>**\*tubes have expiration dates; do not order too many at a time** |
| Urine vacutainer kits | BD 364975 **AND**<br>BD 364979 - Need 1 of each per participant. |
| Vacutainer/urine collection bottle labels | UAB will provide 2"x1" labels preprinted with participant ID and collection tube type |
| Mironic 0.75ml 2D Data-Matrix coded Screw Cap tubes for plasma and serum | MP52757<br>UAB will supply for the entirety of the project |
| Micronic Screw caps for 0.75ml tubes | MP53877 (purple), MP53875 (red), MP53878 (blue), MP53874 (orange)<br>UAB will supply for the entirety of the project |
| Micronic 2ml 2D Data-Matrix | MP52858 |
| coded Screw Cap tubes<br>For buffy coat samples | UAB will supply for the entirety of the project |
| Micronic Screw caps for 2ml tubes | MP53420 (grey)<br>UAB will supply for the entirety of the project |
| Micronic tube labels | UAB will provide 0.87"x0.4" freezer compatible labels preprinted with participant ID |
| Micronic box labels | UAB will provide 2.125"x0.27" preprinted freezer compatible box labels with boc identifiers defined in the preprinted box layout sheets |
| Storage tube - 2ml, skirted | ThermoFisher 02-681-343 - 4 per participant |
| Storage tube caps with O-ring | ThermoFisher 02-681-358 - 4 per participant |
| Storage tube freezer box<br>(Used for NORC lab samples) | 2" storage box with 9x9 tube grid - Can vary by site (UAB uses ThermoFisher 03-391-515). Will need separate boxes for each sample type (plasma, serum, whole blood, urine) |
| Rubber bands | For securing NORC boxes for shipping - we suggest Staples Economy Rubber Bands, #16 (28616-CC) |
| Transfer/Mixing Tube | 5mL Eppendorf or 15mL conical tube |
| Storage tube labels | UAB will provide |
| Storage tube box label | UAB will provide 2"x1" preprinted labels with incrementing box numbers |
| Blank labels | UAB will provide blank labels as needed |
| Marker, permanent/waterproof | Vary by site |
| Transfer disposable pipettes | Can vary by site (UAB uses Sarstedt 86.11741.300) |
| Micropipette+tips | Can vary by site 400ul-1000ul range required |
| Laptop or computer for entering barcode data into spreadsheet | Vary by site |
| 2D Bar code reader for micronics tubes |  |
| Access to -80°C freezer | Varies by site |
| **Biospecimen Shipping** |  |
| Same day shipper<br>(CPT/Paxgene RNA tubes) | ThermoFisher 03-530-35 (Sonoco 409) Therapak™ 409 |
| Same day shipper IATA required accessories: | Fisher 22-130-039 for Therapak™ 10300 Absorbant material<br>Fisher 22-130-040 for Therapak™ 10306 Absorbant material<br>Fisher 22-311-100 Minigrip LLC SBL2X810B5 specimen transport bags<br>Any sealing packing tape (for example, Staples 886404) |
| Batch/dry ice shipper | Small shipper - ThermoFisher 22-130-407 |
| Batch/dry ice shipper | Medium shipper - ThermoFisher 22-130-321 |
| Batch/dry ice shipper | Large shipper - ThermoFisher 22-130-320 |
| Dry Ice | Varies by site |
| Batch/Dry ice shipper IATA required accessories | Varies by site - IATA requires appropriate labeling |
| Self-seal Bubble bags | ULINE S-682 (6"x8.5") or equivalent |
| FedEx Clinical Paks-large | FedEx provides for free |
| **Monitors and Sensors** |  |
| Dexcom G6 | Purchased by UW and shipped to each site from Dexcom.<br>**Note:** Do not order all units at once. Order 3-4 months in advance. Dexcom units expire. |
| Physical Activity Monitor (Garmin watches) | Vivosmart 5, Part # 010-02645-00 |
| Environmental Sensor AND location reporting card | Supplied by UW |
| FedEx boxes for device return | Free from FedEx: FedEx Express box (FedEx Box size: 8-3/4" x 2-5/8" x 11-1/4", FedEx Express box S2, part number 167027) |
| Bubble wrap sleeve or other packaging for device return | Bubble wrap sleeves -> something like this |
| Printed instructions for return of devices | Print from file |

## Data Availability

The data collected in this study is available to researchers through a public access or a controlled access database, depending on the variables requested. A licensing process is required for access and is described at https://aireadi.org/

## 16. Abbreviation Glossary

AI: Artificial Intelligence
API: Application Programming Interface
BCVA: Best-Corrected Visual Acuity
BED: Browser Extensible Data format:
BIDS (Brain Imaging Data Structure): standards used for organizing neuroimaging (mRIs) and behavioral data.
CES-D-10: Center of Epidemiologic Studies Depression Scale, 10-item version
CRC: Clinical Research Coordinator
CRU: Clinical Research Unit
CGM: Continuous Glucose Monitoring
DICOM: Digital Imaging and Communications in Medicine: standards used for communication and management of medical imaging information and data. Will be used for retinal imaging.
EHR: Electronic Health Record:
EKG/ECG: Electrocardiogram: Device that records electrical signals from the heart, shown as waves
ETDRS: Early Treatment Diabetic Retinopathy Study: test used for visual acuity; based on Sloan letters
EVA: Electronic Visual Acuity
FAIR: Findable, Accessible, Interoperable, Reusable Data: High level principles for making digital research objects reusable for humans and machines. Progressively adopted by all stakeholders. Fairhub.io: open-source, cloud-based free platform for preparing, sharing, and accessing AI-READI datasets.
FLIO: Fluorescence Lifetime Imaging Ophthalmoscopy
IRB: Institutional Review Board:
LOINC: Logical Observation Identifiers, Names and Codes: International standard for identifying health measurements, observations and documents. It is one of several designated standards for use in the U.S. Federal Government system for the electronic exchange of clinical health information.
ML: Machine Learning
MoCA: Montreal Cognitive Assessment: A brief, 30-question test that helps detect mild cognitive impairment
NIH: National Institute of Health
OCT: Optical Coherence Tomography: cross-sectional pictures of the retina, allowing for the analysis of retina layers and retinal thickness. Used to view anatomical structure.
OCTA (Optical Coherence Tomography Angiography): sequential scans that detect and illustrate movement of blood/particles in the posterior eye. Used to view vascular structure.
OD: Right eye
OMOP: Observational Medical Outcomes Partnership Data Model: standards used for clinical information, research visits, and serological data.
ONH: Optic Nerve Head
OS: Left eye
OU: Both eyes
PAID-5: Problem Areas In Diabetesicense-free collection of standardized measuremvalidated questionnaire that queries for diabetes-related impact on lifestyle
PhenX Toolkit: a license-free collection of standardized measurement protocols for enhancing biomedical and clinical research.
PHI: HIPAA protected Personal Health Information
REDCap: Research Electronic Data Capture: a secure, web-based platform for managing surveys and research data
SDOH: Social Determinants of Health
T2DM: Type 2 Diabetes Mellitus
UAB: University of Alabama at Birmingham
UCSD: University of California San Diego
UW: University of Washington

## 17. Acknowledgements

This work was supported by the National Institutes of Health (NIH) grants OT2OD032644, P30DK035816 and UL1TR003096 and Research to Prevent Blindness. We thank the Microsoft AI for Good Lab for supporting the cloud services needed for the project. We thank Topcon Corporation (Tokyo, Japan), Optomed (Oulu, Finland), iCare World (Raleigh, NC), and Carl Zeiss (Oberkochen, Germany) for loaning their devices for research purposes at no cost. We thank Heidelberg Engineering (Heidelberg, Germany), Dexcom (San Diego, CA), and Garmin (Olathe, KS) for research discounts on study devices. We also thank the study participants and the AI-READI Advisory Council.

## 20. Appendix

The following pages contain AI-READI-specific documents that support recruitment, protocol details, return of data and data interpretation.

### APPENDIX A: Recruitment Letter

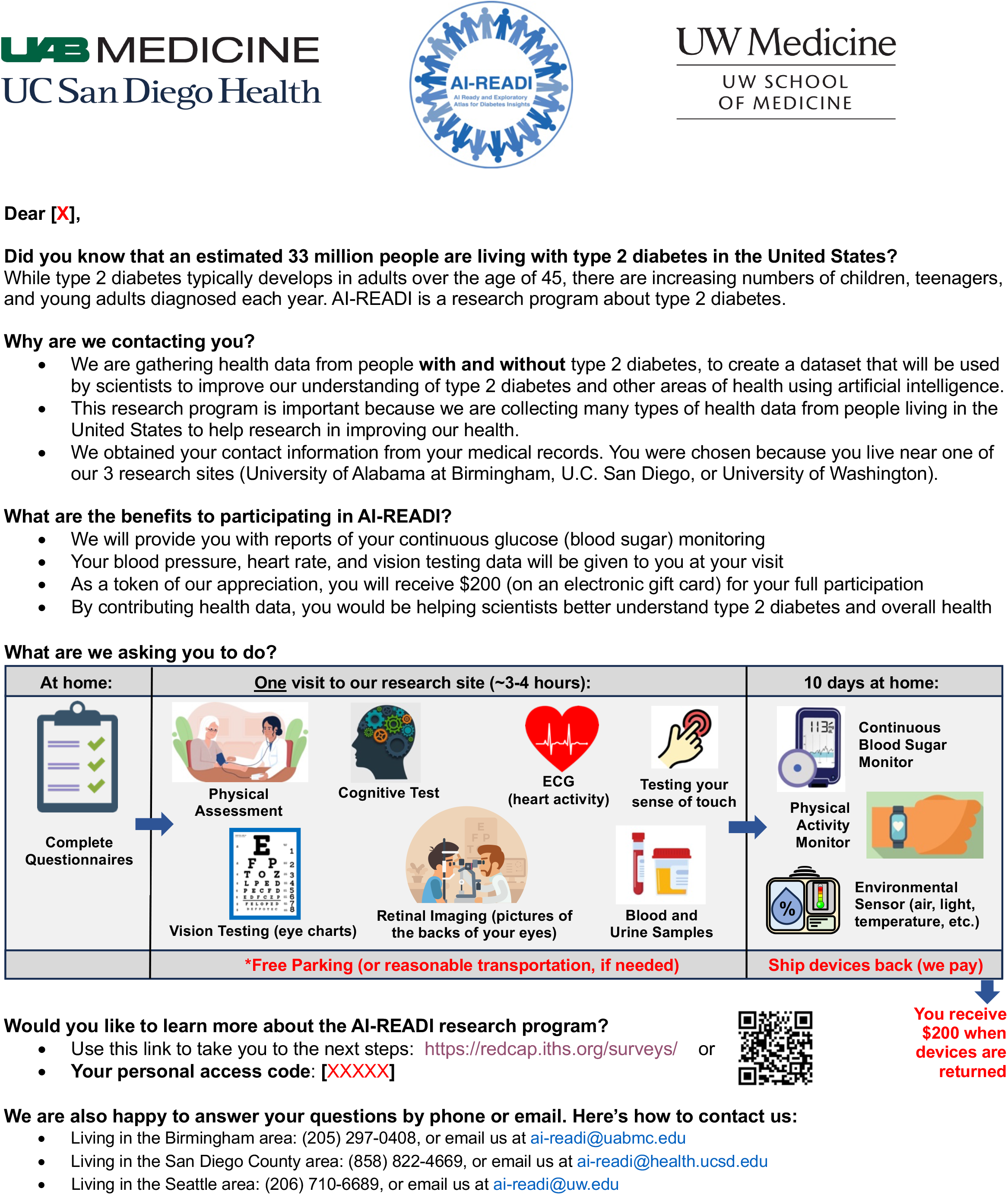

### APPENDIX B: Participant Return of Data Card

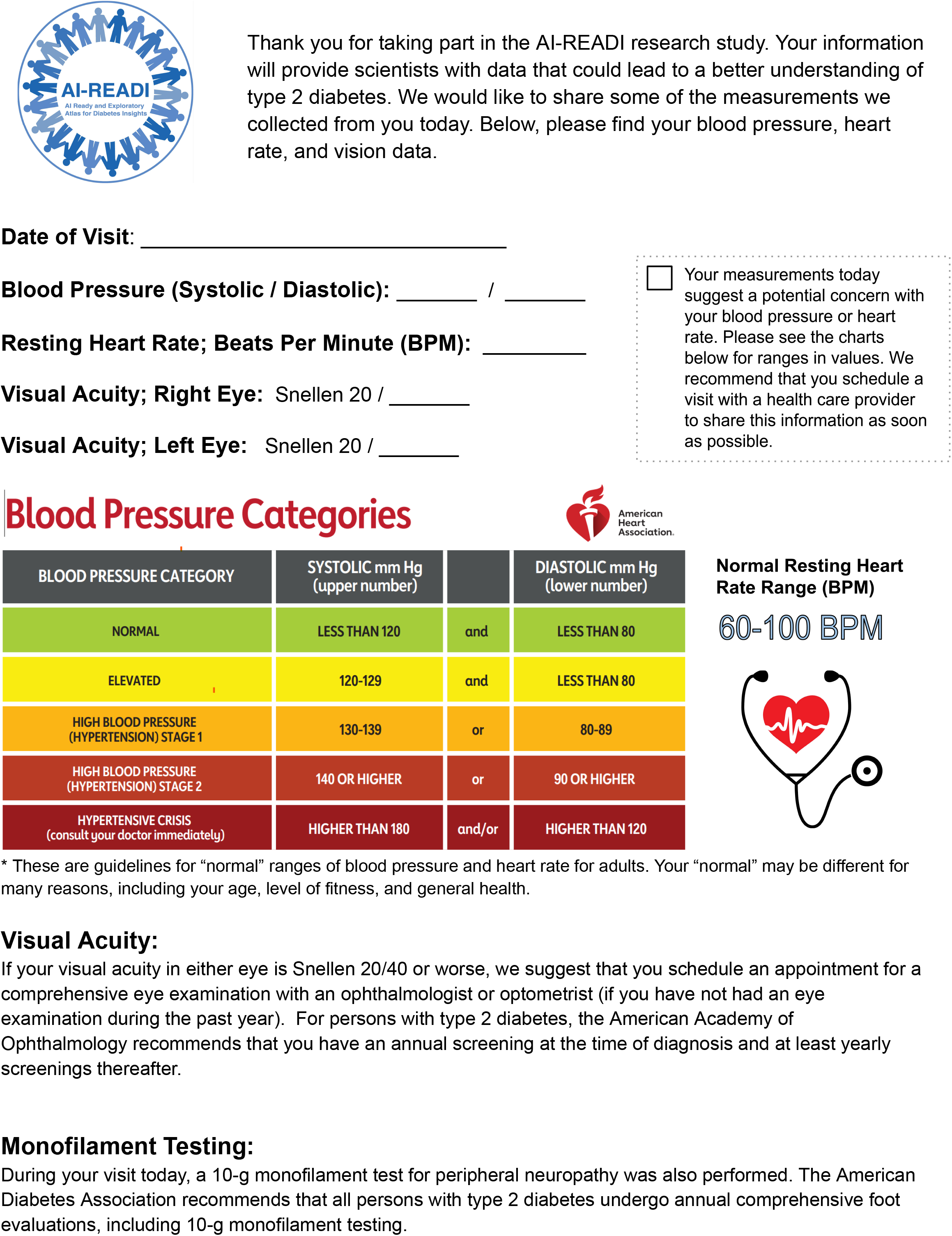

### APPENDIX C

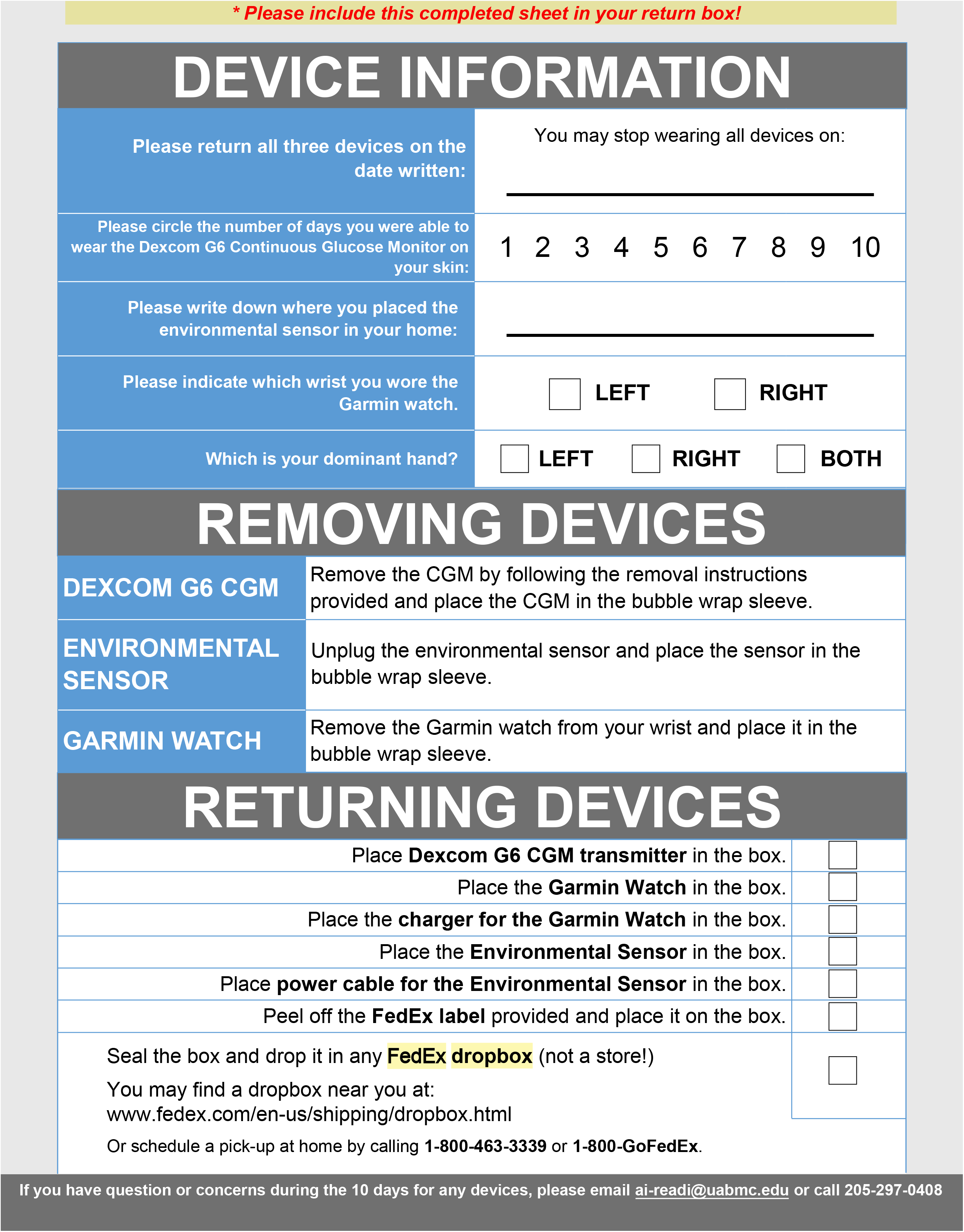

### APPENDIX D

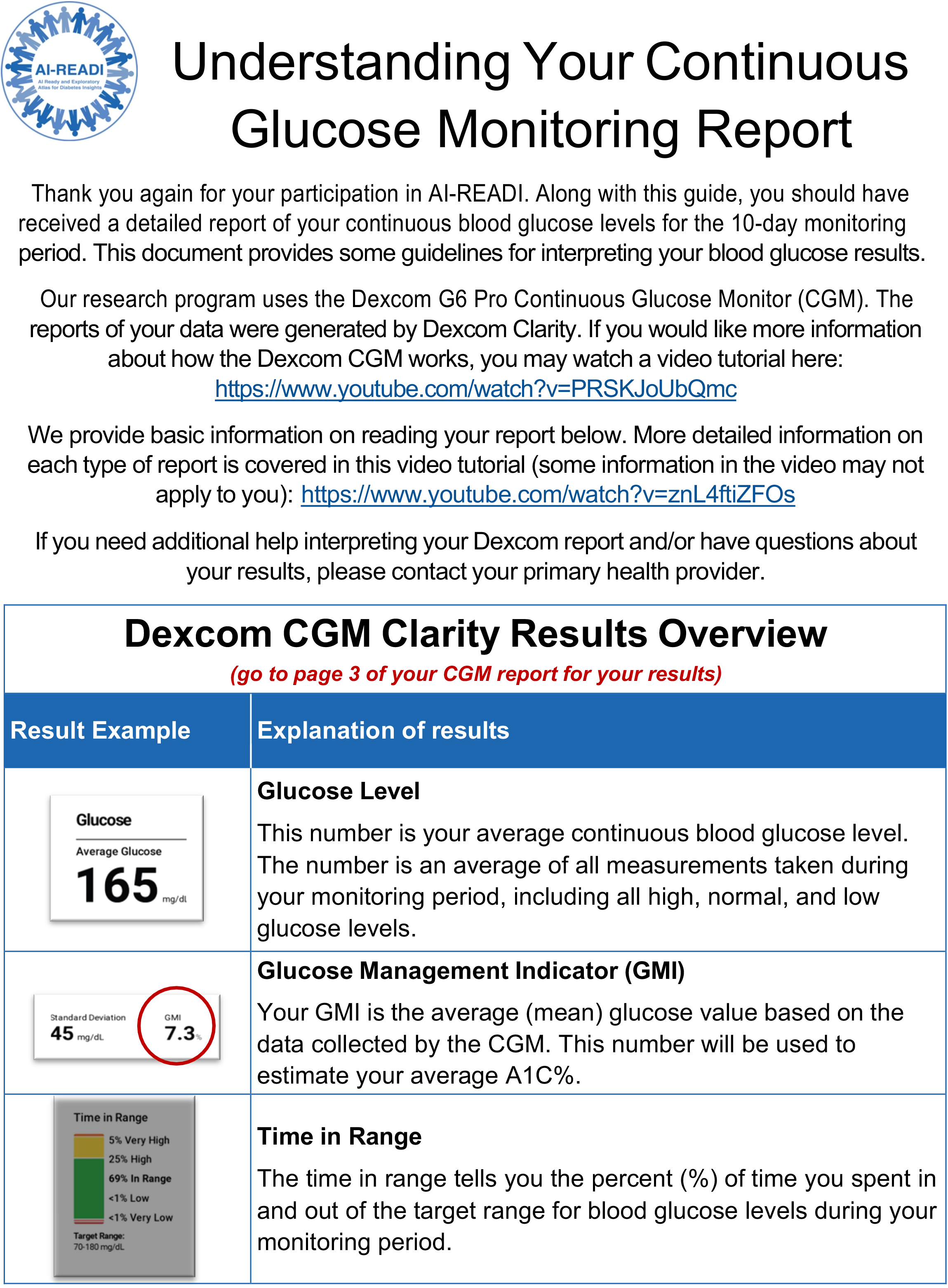
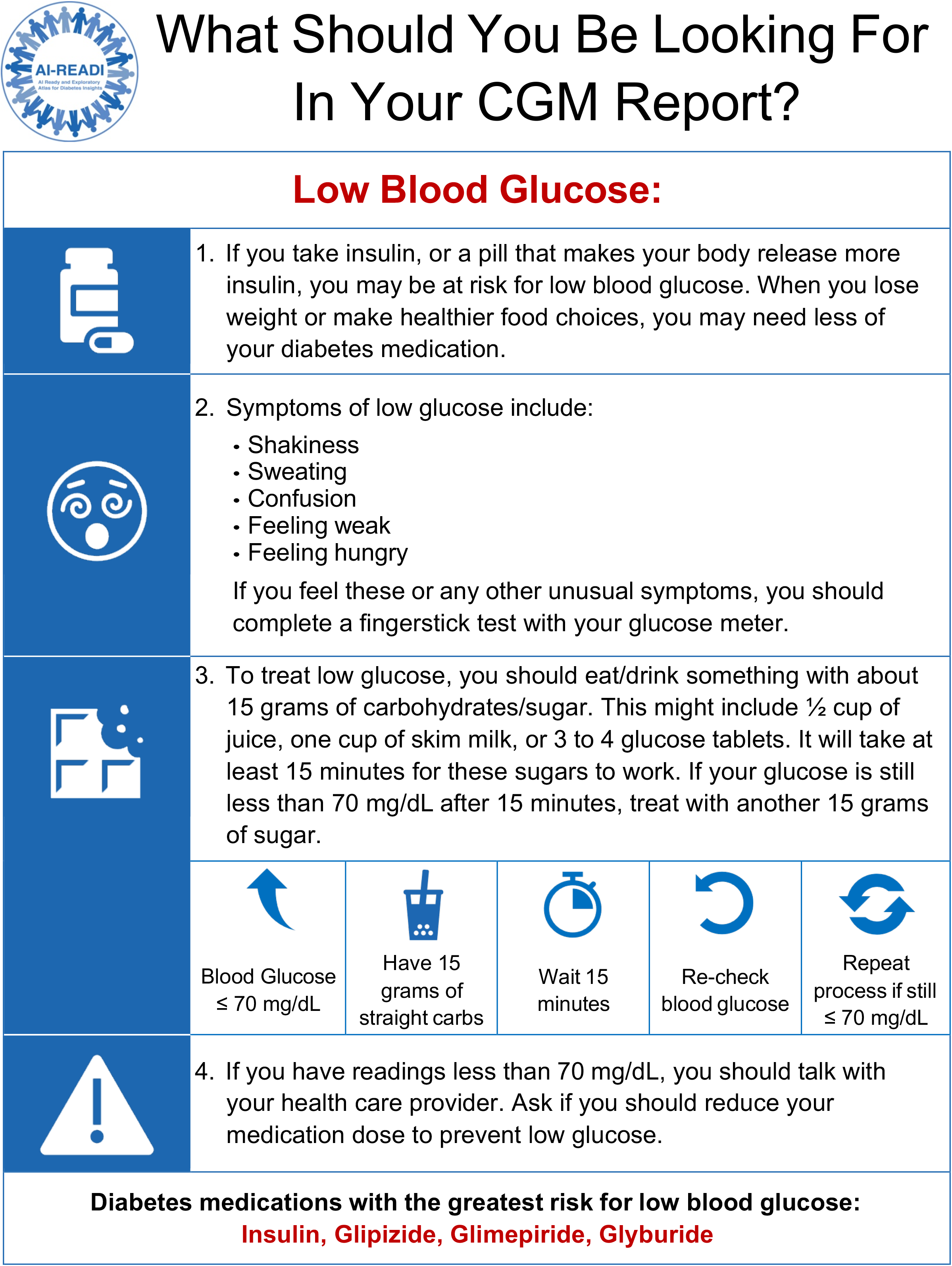
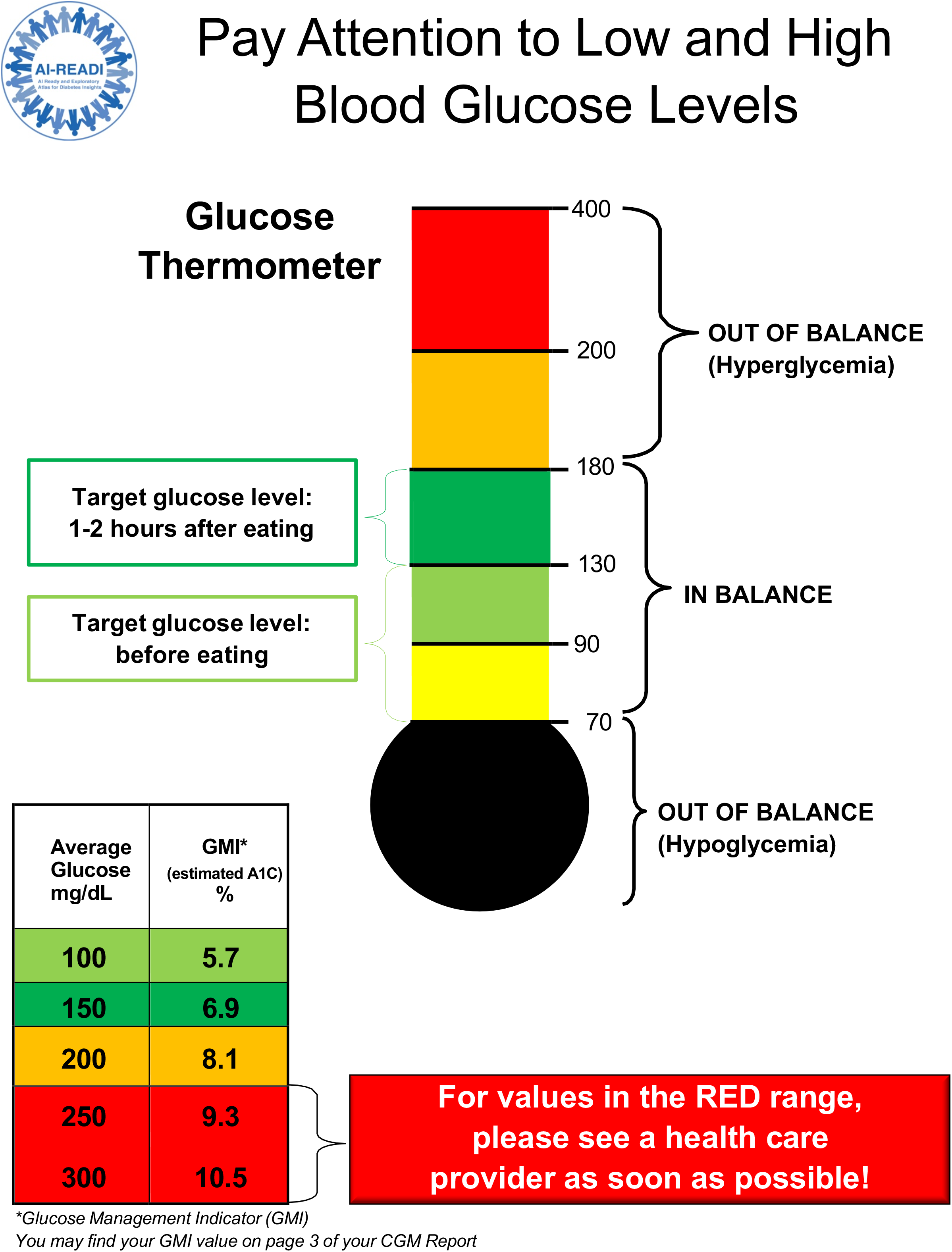
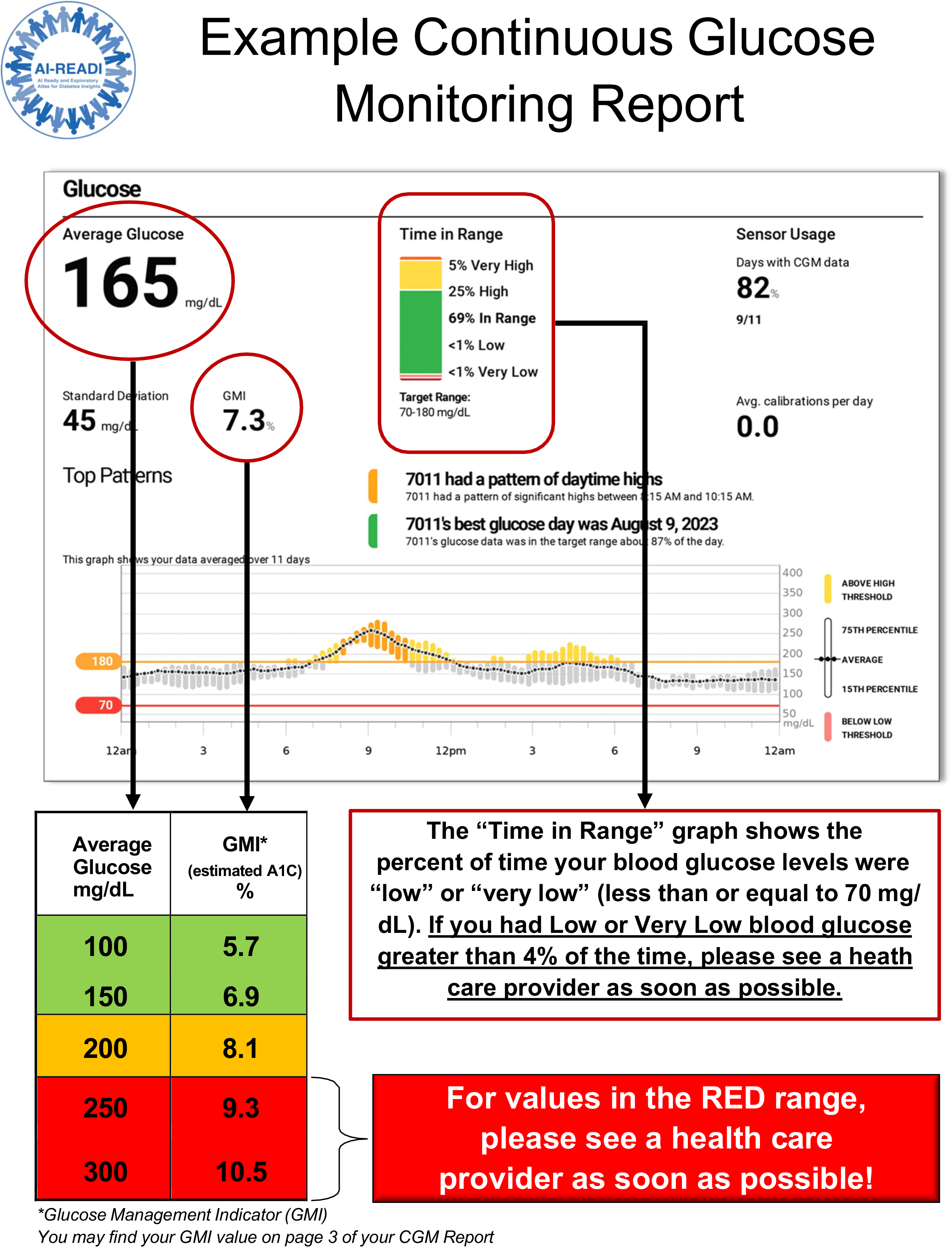

### APPENDIX E: Retinal Imaging Checklist

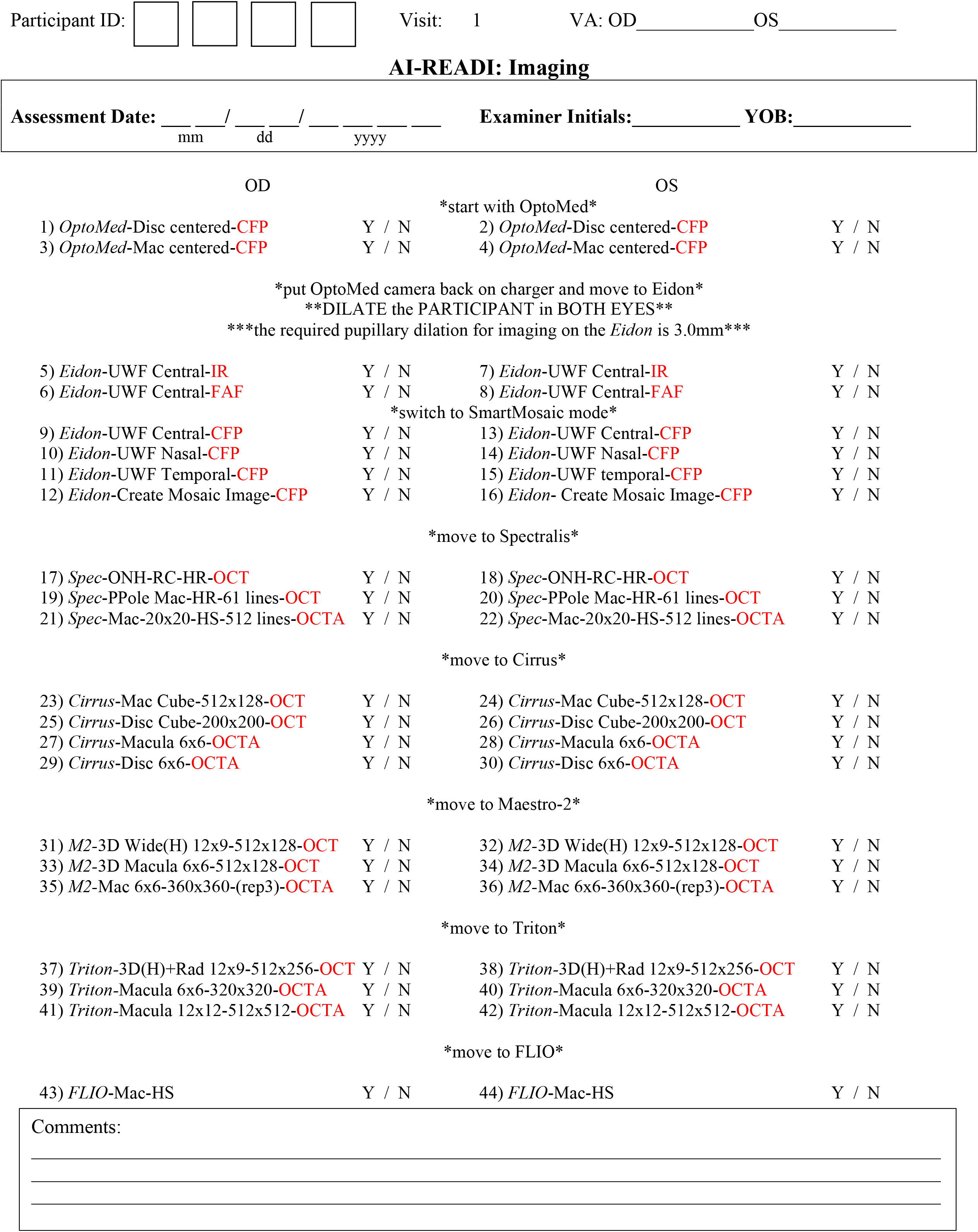

### APPENDIX F: Participant Return of Lab Test Results

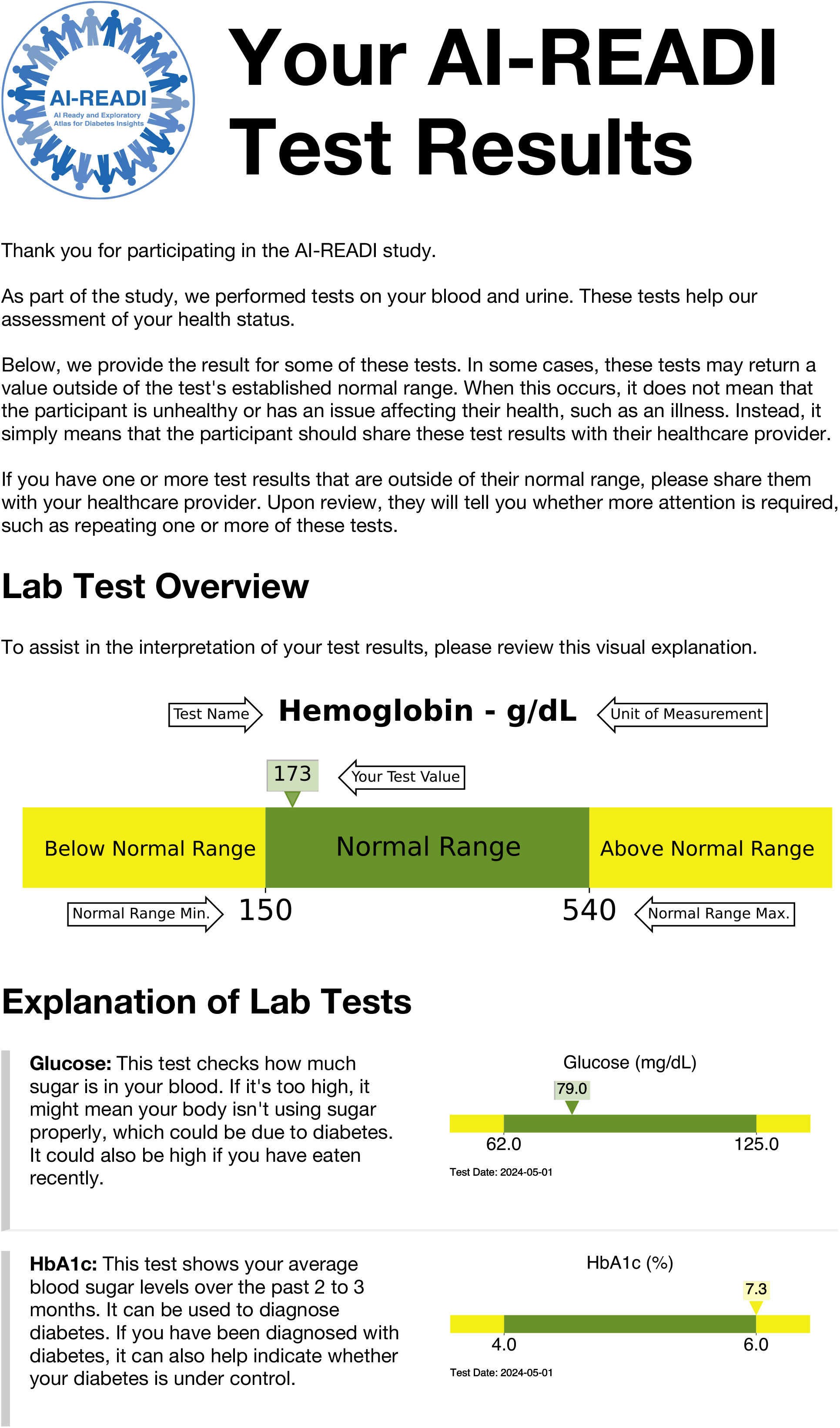
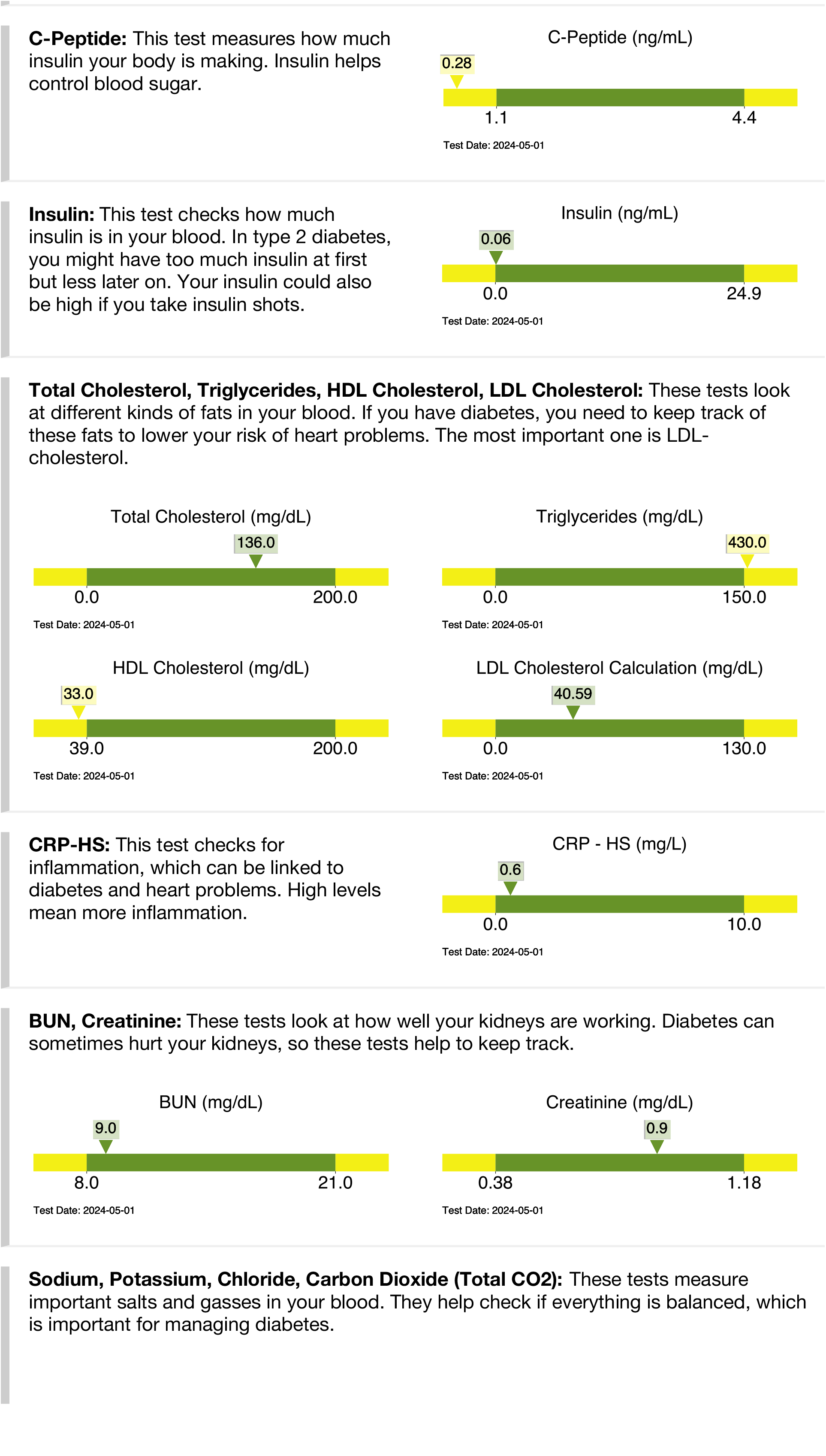
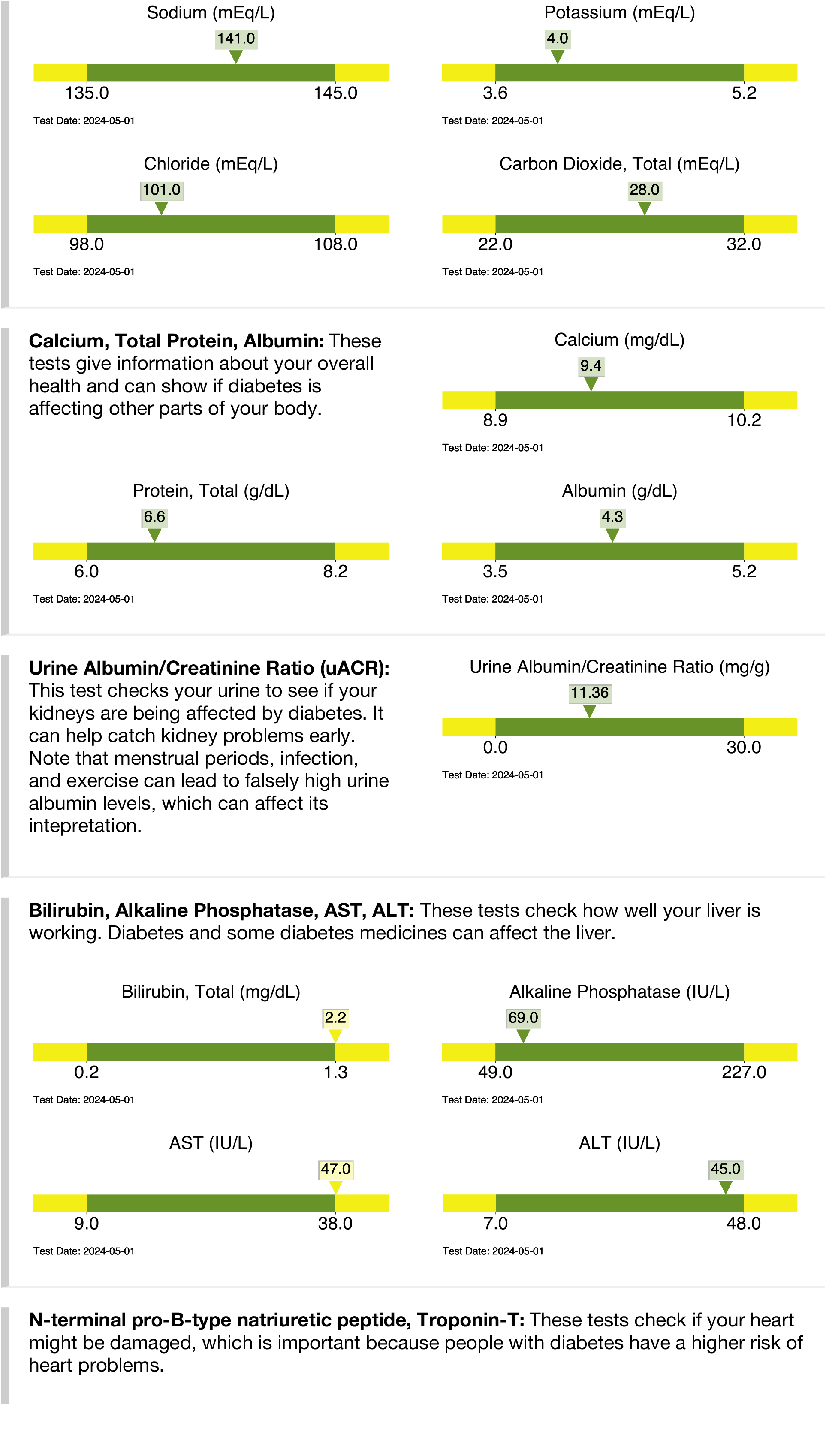
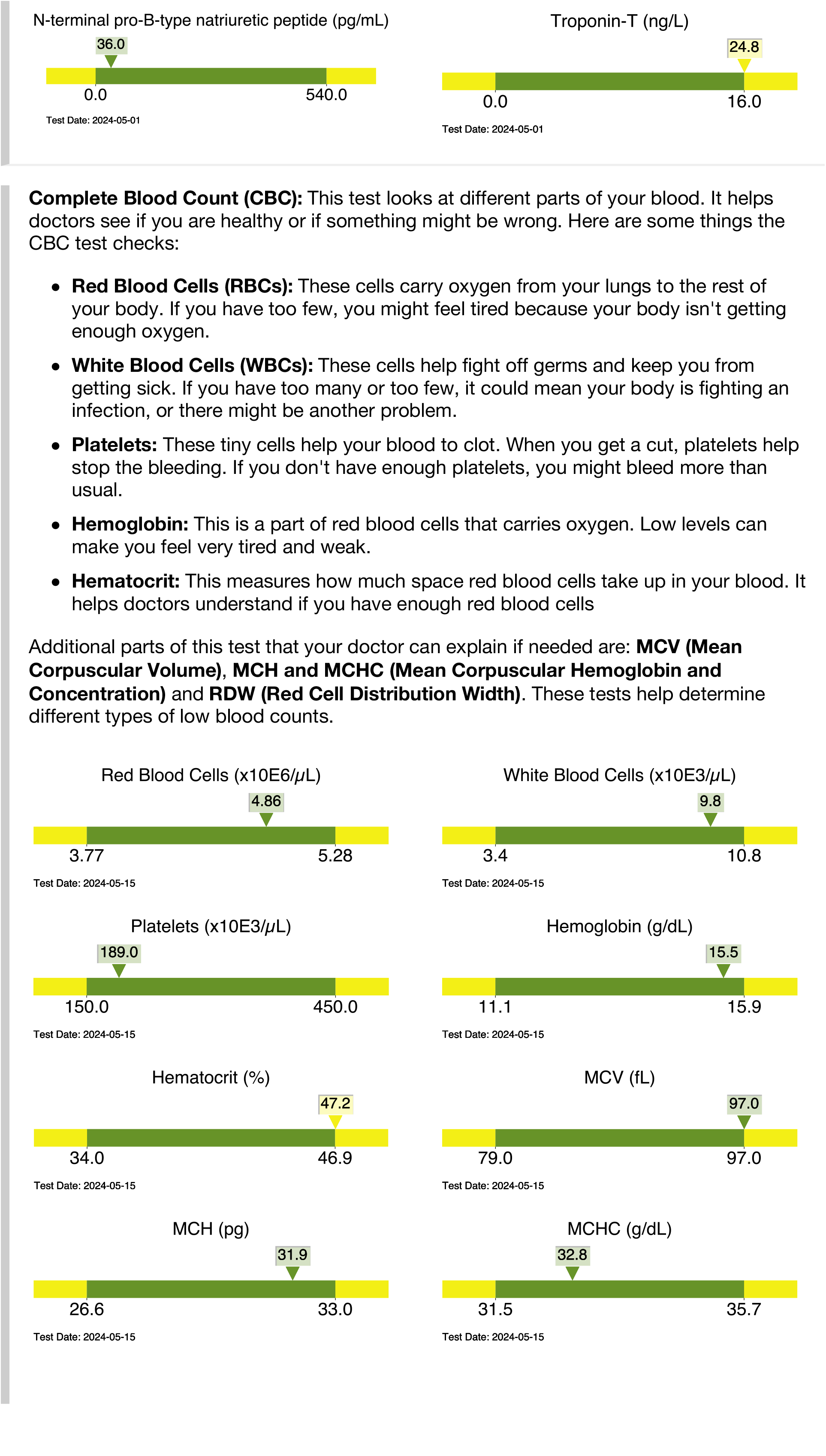
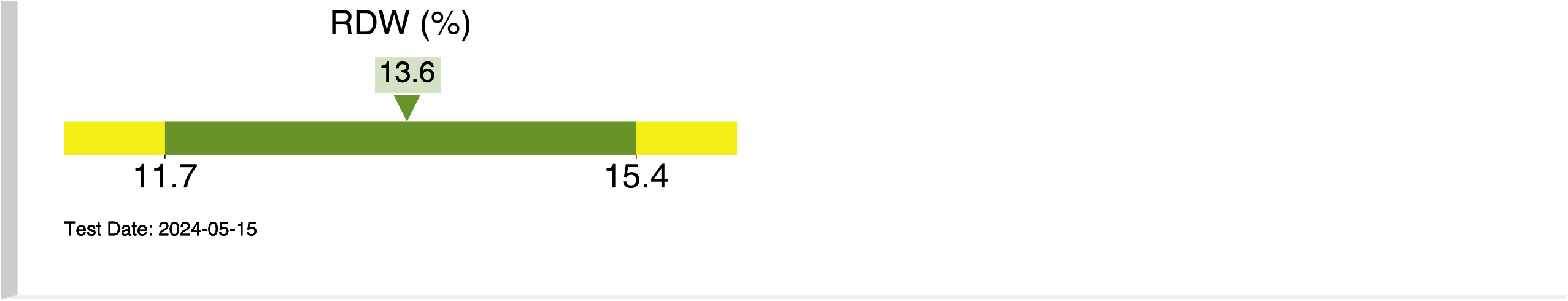

### APPENDIX G

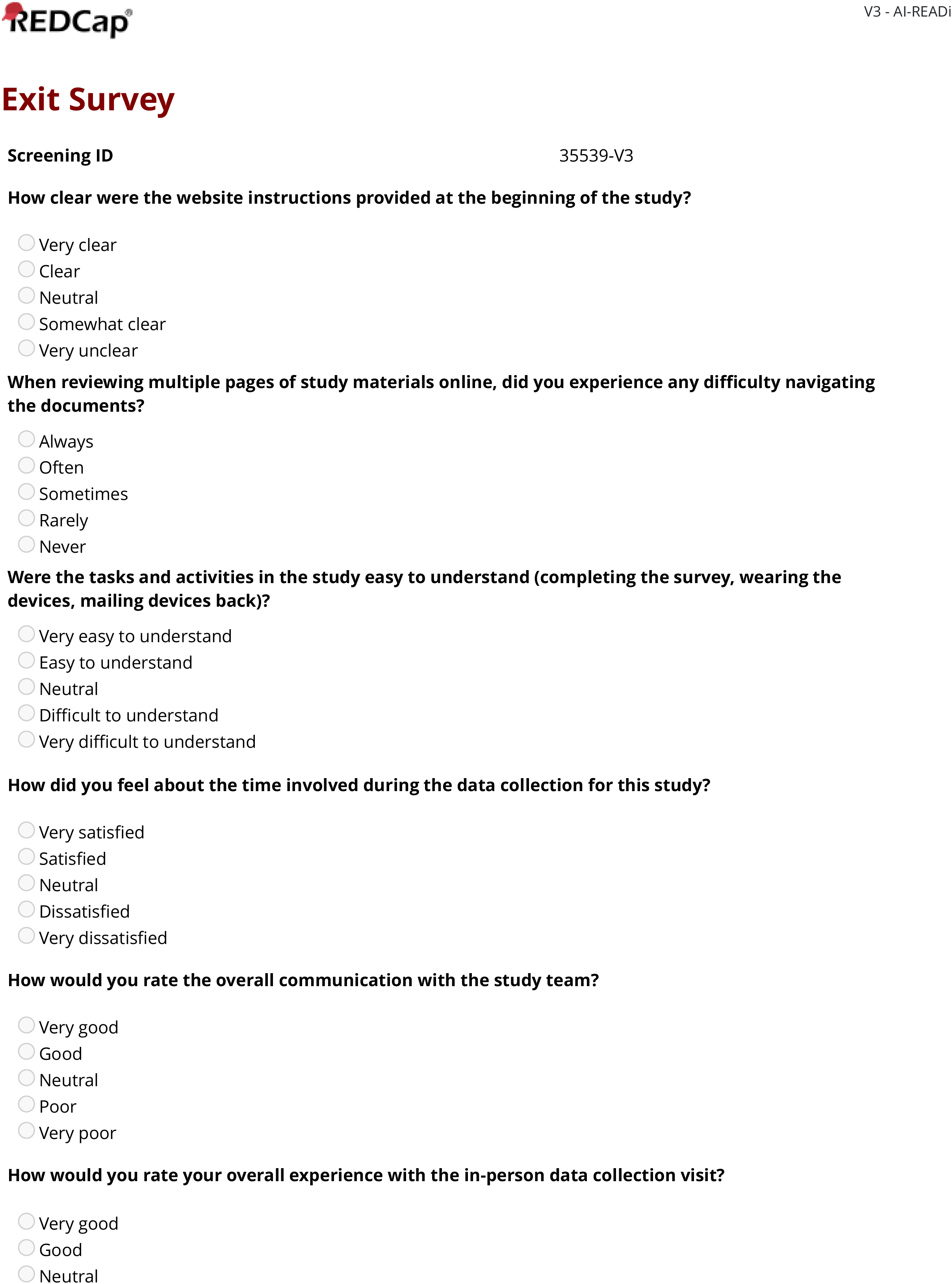
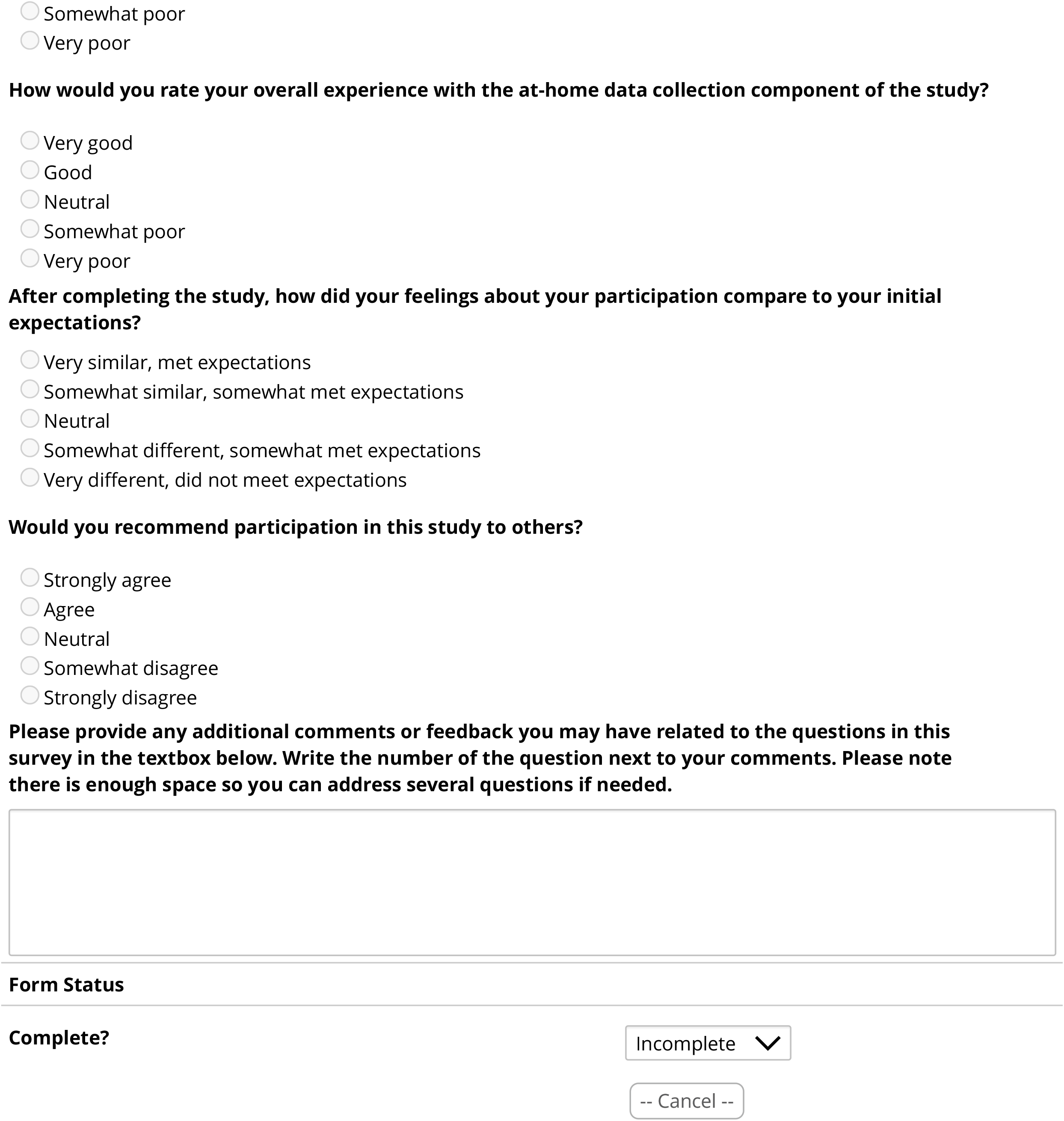

